# Uncovering causal gene-tissue pairs and variants: A multivariable TWAS method controlling for infinitesimal effects

**DOI:** 10.1101/2024.11.13.24317250

**Authors:** Yihe Yang, Noah Lorincz-Comi, Xiaofeng Zhu

## Abstract

Transcriptome-wide association studies (TWAS) are commonly used to prioritize causal genes underlying associations found in genome-wide association studies (GWAS) and have been extended to identify causal genes through multivariable TWAS methods. However, recent studies have shown that widespread infinitesimal effects due to polygenicity can impair the performance of these methods. In this report, we introduce a multivariable TWAS method named Tissue-Gene pairs, direct causal Variants, and Infinitesimal effects selector (TGVIS) to identify tissue-specific causal genes and direct causal variants while accounting for infinitesimal effects. In simulations, TGVIS maintains an accurate prioritization of causal gene-tissue pairs and variants and demonstrates comparable or superior power to existing approaches, regardless of the presence of infinitesimal effects. In the real data analysis of GWAS summary data of 45 cardiometabolic traits and expression/splicing quantitative trait loci (eQTL/sQTL) from 31 tissues, TGVIS is able to improve causal gene prioritization and identifies novel genes that were missed by conventional TWAS.

## Introduction

Over the past two decades, genome-wide association studies (GWAS) have identified thousands of genetic variants associated with complex traits^1–3^. However, most GWAS signals are detected in non-coding regions and have been shown to have complex regulatory landscapes across different tissues and cell types^4^, making it challenging to pinpoint causal variants and genes driving these GWAS signals. Joint GWAS and expression quantitative trait loci (eQTL) data analysis methods, such as colocalization^5^, transcriptome-wide association studies (TWAS)^6^, and *cis*-Mendelian randomization (*cis*-MR)^7^, have been developed to prioritize causal genes at GWAS loci^8^. Colocalization simultaneously examines the expression of a gene and a trait to determine whether they share common causal genetic variants at a locus^5^. Both TWAS and *cis*-MR assume a causal diagram where eQTLs regulate tissue-specific gene expression that subsequently affects a trait, and they identify these tissue-specific causal genes by testing the significance of the causal effect estimates. Furthermore, these methods have been extended to a broader range of molecular phenotypes, such as splicing events^9^ and protein abundance^10^, with regulatory QTLs being splicing QTLs (sQTLs) and protein QTLs (pQTLs), which we call xQTLs in general.

Nevertheless, colocalization, TWAS, and *cis*-MR are all univariable methods that statistically measure the marginal correlations of genetic effect sizes between a trait and a tissue-specific expression of a gene. Non-causal gene-tissue pairs may be falsely detected by these univariable methods due to the *cis*-gene-tissue co-regulations with causal gene-tissue pairs^8,11,12^. The underlying mechanism may come in the following respects: the tissue-specific eQTLs of a causal gene are in linkage disequilibrium (LD) with (1) the eQTLs of nearby non-causal genes^13^ and (2) the eQTLs of causal genes expressed in non-causal tissues^14^. In addition, some variants can influence a trait independently of causal gene-tissue pairs, which are frequently denoted as direct causal variants^13^ and horizontal pleiotropy^15^. The non-causal gene-tissue pairs may be incorrectly detected when their eQTLs are in LD with direct causal variants.

Multivariable TWAS methods, such as causal TWAS (cTWAS)^13^ and Tissue-Gene Fine-Mapping (TGFM)^14^, have been proposed to address these issues. Specifically, cTWAS identifies causal genes and direct causal variants among multiple candidates using the sum of single effects (SuSiE)^16,14^ by examining tissues separately. TGFM extends cTWAS to allow multiple tissues to be analyzed simultaneously and can identify the trait-relevant tissues beyond the causal variants and genes. However, Cui et al.^17^ recently reported that current Bayesian fine-mapping methods, including SuSiE^16,18^ and FINEMAP^19^, have a high replication failure rate (RFR) in practice. Cui et al.^17^ discovered that the widespread infinitesimal effects, which may stem from the polygenicity of complex traits, are the sources of the high RFR, and accounting for the infinitesimal effects can reduce the RFR and improve statistical power. Similarly, the polygenicity can also lead to inflating the test statistics in standard TWAS^20^ and traditional linkage studies^21^. Thus, due to the lack of modeling infinitesimal effects, it is expected that cTWAS and TGFM can be vulnerable to spurious prioritization and reduced statistical power.

We present the <u>T</u>issue-<u>G</u>ene pair, direct causal <u>V</u>ariants, and Infinitesimal effect <u>S</u>elector (TGVIS), a multivariable TWAS method to identify causal gene-tissue pairs and direct causal variants while incorporating infinitesimal effects. TGVIS employs SuSiE^16,14^ for fine-mapping causal gene-tissue pairs and direct causal variants, and uses restricted maximum likelihood (REML)^22^ to estimate the infinitesimal effects. In addition, we introduce the Pratt index^23^ to rank the importance for improving the prioritization of causal genes and variants. We applied TGVIS to identify causal *cis*-gene-tissue pairs and direct causal variants for 45 cardiometabolic traits using GWAS datasets with the largest sample sizes to date^3,24–32^, by incorporating the eQTL and sQTL summary statistics from 28 tissues from genotype-tissue expression (GTEx)^12^, and the eQTL summary statistics of kidney tubulointerstitial^33^, kidney glomerular^33^, and pancreatic islets^34^ tissues. We summarized the causal gene-tissue pairs and direct causal variants, highlighted the pleiotropic effects at the gene-tissue level, and demonstrated the different functional activity^35^ of eQTLs/sQTLs mediated through gene-tissues and the direct causal variants. Moreover, we mapped the trait-relevant major tissues and demonstrated the enrichments of genes identified by TGVIS in terms of colocalization^5^, on the silver standard of lipid genes^13^, FDA-approved drug-target genes^36^, and genes detected through pQTL summary data^10^. Our study reveals a broader picture of gene and tissue co-regulations, which can provide novel biological insights into complex traits.

## Results

### Overview of method

**Figure 1A** illustrates the causal diagram assumed in this report. Specifically, we hypothesize that a set of xQTLs influence the products of genes (e.g., expressions and splicing events) at a locus. Gene co-regulation^8,12^, i.e., the correlation of xQTL effects among multiple gene products, can emerge due to shared xQTLs or being in LD among them. Meanwhile, tissue co-regulation^11,37,38^, defined as the correlation of gene expression across multiple tissues, can arise because of the same mechanism. In the gene and tissue co-regulation network, certain gene-tissue pairs directly influence a trait without mediation by other gene-tissue pairs, which are referred to as causal gene-tissue pairs. In addition, some genetic variants may directly influence the trait, which we consider as direct causal variants. Besides these direct causal variants which have relatively large effects, we assume there are polygenic or infinitesimal effects that can be modeled through a normal distribution with mean zero and small variance^17^ (**Method**).

**Figure 1:**
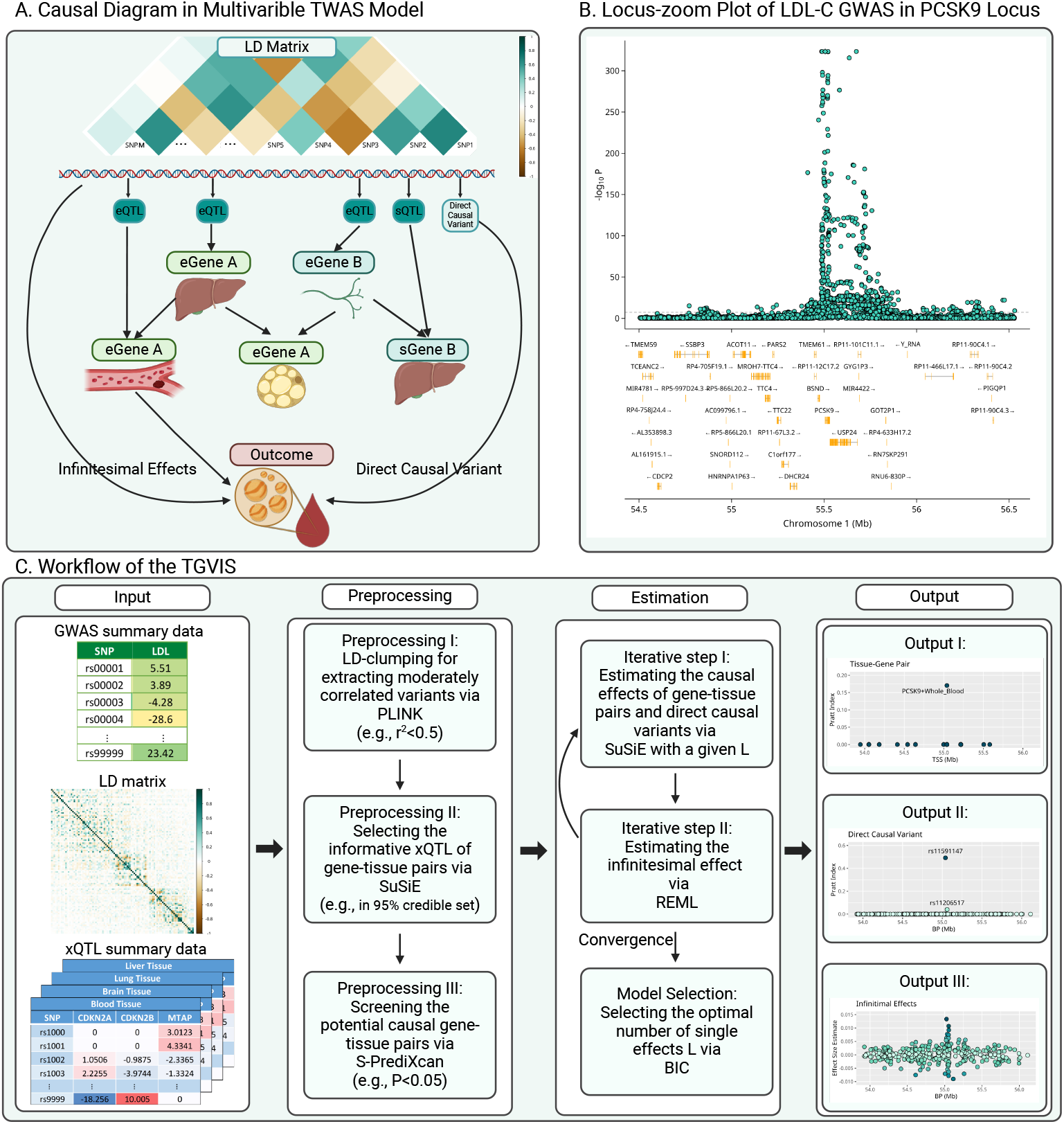
Overview of TGVIS. **A**: A hypothetical causal diagram illustrating the relationships between variants (including xQTLs, direct causal variants, and non-causal variants), tissue-specific gene expressions, and an outcome in a *cis*-region, where the arrows indicate the flow of causal effects in the causal diagram. Variants may be in LD, with only a subset having cis-regulatory effects. Gene expressions or splicing events are tissue-specific and form a complex co-regulation network. Only molecular phenotypes directly connected to the outcome are considered causal. **B**: Locus-zoom plot of the LDL-C GWAS in the *PCSK9* locus. The bottom panel displays the coding regions of genes located within this locus, including *PCSK9, UPS24, BSND*, etc. **C**: Workflow of TGVIS, consisting of three main steps. (I) Input, including GWAS summary data, eQTL summary data from multiple tissues, and LD matrix. (II) Preprocessing, including eQTL selection and prescreening. We applied S-Predixcan to pre-screen some noise pairs, aiming to reduce the dimension of the multivariable TWAS model to a reasonable scale. (III) Estimation, where TGVIS first selects the causal genetissue pairs and direct causal variants via SuSiE and then estimates the infinitesimal effect via REML. (IV) Output, including the causal effect estimate, direct causal effect estimate, and infinitesimal effect estimates. We output plots demonstrating the causal gene-tissue pairs, direct causal variants and predicted infinitesimal effects: (1) the Pratt indices and other statistics such as PIPs, estimates, SEs of causal gene-tissue pairs in the 95% credible sets, (2) the Pratt indices of the direct causal variants in the 95% credible sets, and (3) the best linear unbiased predictors of infinitesimal effects. The non-zero variance in output III in this figure suggests the non-zero contribution of infinitesimal effects.

The curse of dimensionality poses a substantial challenge in the multivariable TWAS model. **Figure 1B** illustrates this challenge by an example of the association evidence with low-density lipoprotein cholesterol (LDL-C)^1^ at the *PCSK9* locus, where dozens of coding genes and long non-coding ribonucleic acids (RNAs) are located, along with multiple potential direct causal variants. Conventional statistical methods cannot precisely identify causal gene-tissue pairs and variants because there are many correlated candidates which frequently range from hundreds to thousands^16^. The proposed TGVIS enables to overcome the curse of dimensionality. **Figure 1C** describes the workflow of TGVIS, where the inputs are the GWAS summary statistics of a trait, xQTL summary statistics of gene-tissue pairs, and a reference LD matrix of the variants at the locus. TGVIS utilizes a profile-likelihood approach to estimate the causal effects of gene-tissue pairs and directly causal variant effects with SuSiE^16,18^ and model the infinitesimal effects via REML^22^. This profile-likelihood iterates until all estimates are converge. The details are described in the **Methods** and the **Supplementary Materials**.

In practice, another challenge arises when selecting a causal gene-tissue pair based solely on its posterior inclusion probability (PIP) because many gene-tissue pairs share the same sets of xQTLs at a locus, making them statistically indistinguishable. SuSiE groups these pairs into a credible set during fine-mapping and introduces a single effect to describe the contributions of the variables in the same credible set. Therefore, all inferences should be made based on the single effects defined by SuSiE’s credible sets. To quantify the contribution of each gene-tissue pair and direct causal variant, we introduce the Pratt index^23^ as a metric parallel to PIP. While PIP measures the significance of variables from a Bayesian viewpoint, the Pratt index quantifies their predictive importance. In the application, we calculated the cumulative Pratt index of variables in a 95% credible set (CS-Pratt) and filtered out the credible sets with low CS-Pratt values (**Methods** and **Supplementary Materials)**. We observed that this procedure improved the precision of causal gene and variant identification.

### Simulation

We compared the TGVIS with 4 multivariable MR and TWAS methods: *cis*IVW^39^, Grant2022^40^, cTWAS^13^, and TGFM^14^. We applied the following criteria for determining the causality: the 95% credible set for TGVIS, TGFM, and cTWAS; P < 0.05 for *cis*IVW; and selection by lasso in the Grant2022. We did not consider univariable methods because of their high type-I error rates when the goal is to identify causal genes, given that xQTL effect sizes for multiple genes are often correlated^13^. Detailed information on the settings and results can be found in the **Methods** and **Supplementary Materials**.

We first assessed the accuracy of causal effect estimation for gene-tissue pairs. When infinitesimal effects were absent, TGVIS showed a mean square error (MSE) for causal effect estimates similar to that of cTWAS, and TGFM, while both *cis*IVW and Grant2022 exhibited substantially larger MSE (as shown in the left two panels in **Figure 2A)**. However, when infinitesimal effects were present, TGVIS demonstrated a visibly lower MSE compared to the other methods, with cTWAS and TGFM showing approximately 32% higher MSE than TGVIS (as shown in the right two panels in **Figure 2A)**. These results indicate that TGVIS generally outperforms its competitors by accounting for infinitesimal effects.

**Figure 2:**
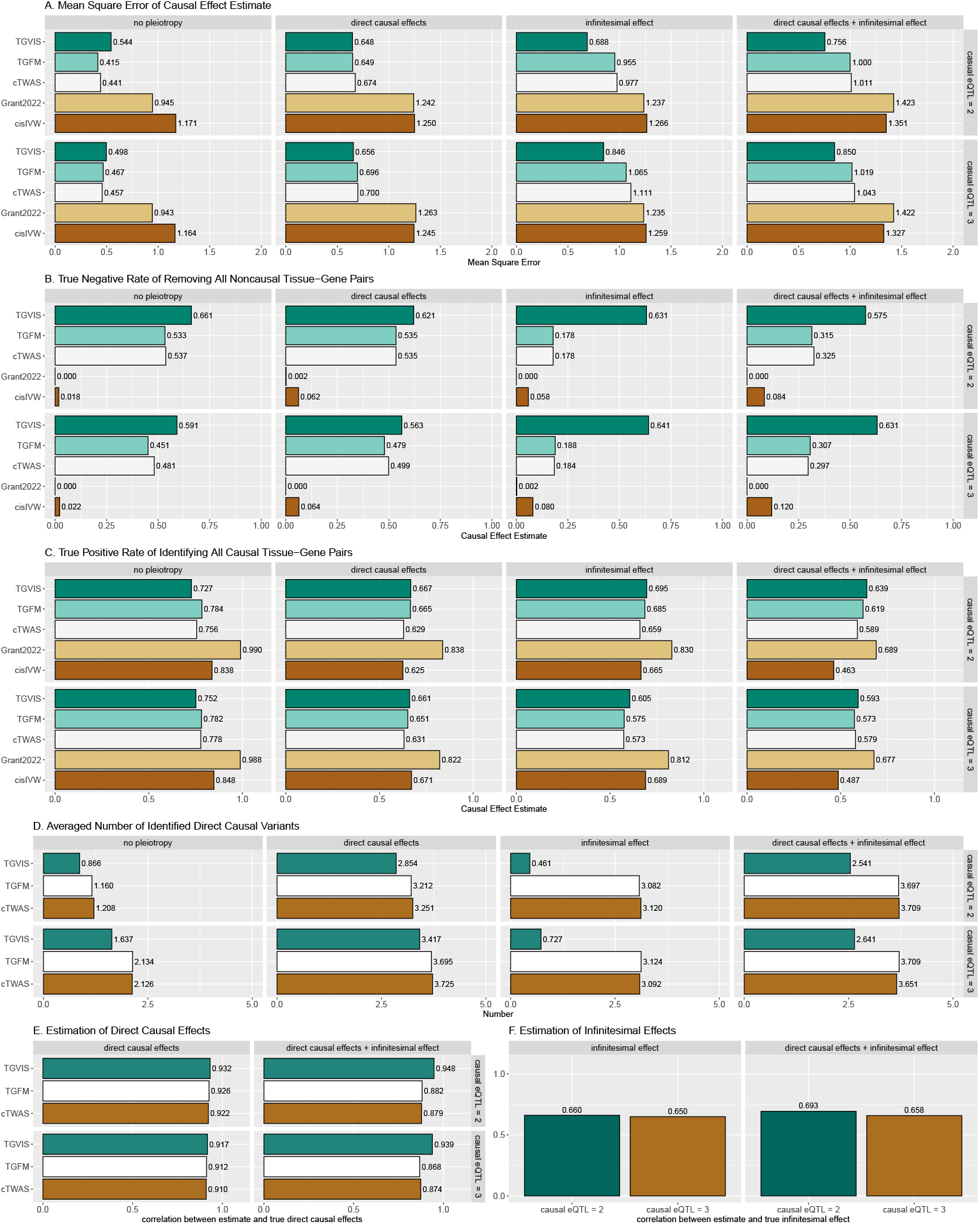
Simulation results comparing the performances of TGVIS, TGFM, cTWAS, Grant2022, and *cis*IVW with xQTL sample size = 200. **A**: The MSE of causal effect estimates under no pleiotropy, in the presence of direct causal variants, infinitesimal effects, and both. **B**: The true negative rate of identifying all 98 non-causal gene-tissue pairs under different scenarios i.e., no pleiotropy, in the presence of direct causal variants, infinitesimal effects, and both. This is equivalent to that if a method incorrectly identifies any non-causal pairs as causal, it will not be counted as a true negative event. **C**: Bar plots display the true positive rates of identifying all 2 causal gene-tissue pairs under different scenarios. **D**: The averaged number of identified direct causal variants by the different methods. The number of true causal variants were set to 0, 2, 0, and 2 for no-pleiotropy, direct-causal-variant, infinitesimal-effects, and direct-causal-variant and infinitesimal-effects, respectively. **E**: The averaged correlation of the true and estimated direct causal effects across simulations. **F**: The averaged correlation of the true and predicted infinitesimal effects across simulations.

We then compared the true negative rate (TNR) and true positive rate (TPR) of these five methods. A true negative is defined as a method that correctly identifies all 98 non-causal gene-tissue pairs. Similarly, a true positive is defined as a method that correctly identifies the 2 causal gene-tissue pairs. Across all the scenarios (**Figure 2B**), TGVIS achieved the highest TNR, with an average of 0.614, followed by TGFM and cTWAS, with average TNRs of 0.513 and 0.499, respectively. *Cis*IVW and Grant2022 performed worst, with averages TNR of 0.064 and 0.013, respectively, indicating that these two methods are prone to identifying a substantial number of false positive gene-tissue pairs. On the other hand, TGVIS exhibited a similar TPR (average TPR = 0.667) as TGFM, cTWAS, and *cis*IVW (average TPRs of 0.649, 0.667, and 0.661, respectively), while Grant2022 had the highest TPR (averages TPR= 0.831) (**Figure 2C**), which is not surprising given that Grant2022 also has lowest TNR.

We further assessed the performance in detecting direct causal variants. In scenarios where no direct causal variants were present, the TGVIS identified fewer direct causal variants, with an average number of 0.92, compared to 2.39 for TGFM and 2.38 for cTWAS (**Figure 2D**). When there were two direct causal variants present, TGVIS identified an average of 2.86 direct causal variants, compared to 3.58 for both cTWAS and TGFM. The averaged correlations between the estimated and true direct causal effects across simulations were high for all three methods (**Figure 2E**). However, predicting infinitesimal effects remains challenging, as evidenced by an average correlation of 0.663 between the predicted and true infinitesimal effects in TGVIS (**Figure 2F**).

### Searching potentially causal gene-tissue pairs and variants for 45 cardiometabolic traits

We systematically analyzed 45 cardiometabolic traits and eQTL/sQTL summary statistics (**Table S1-S2**) to identify potential causal gene-tissue pairs and direct causal variants. For the TVGIS, we considered whether a gene-tissue pair or direct causal variant was causal if (1) it was within a 95% credible set and (2) had a CS-Pratt > 0.15. The criteria of CS-Pratt >0.15 was established based on the empirical evidence (**Methods**). For TGFM, we followed the authors’ recommendation of considering individual PIP > 0.5 as indicative of causality. Additionally, we did not compare with cTWAS because it analyzes tissues separately^13^.

TGVIS and TGFM identified a median of 119.5 and 227.5 causal gene-tissue pairs, and 42 and 183 causal variants per trait (**Figure 3A, Table S3-S6**), respectively. Additionally, TGVIS detected a median of 0.313 causal gene-tissue pairs and 0.115 direct causal variants per locus, while TGFM identified a median of 0.469 causal gene-tissue pairs and 0.466 direct causal variants per locus (**Figure 3C**). Overall, TGVIS reduced the number of causal gene-tissue pairs by a median of 55.7% and the number of direct causal variants by 24.5% per trait compared to TGFM, representing the improved resolution of TGVIS over TGFM.

**Figure 3:**
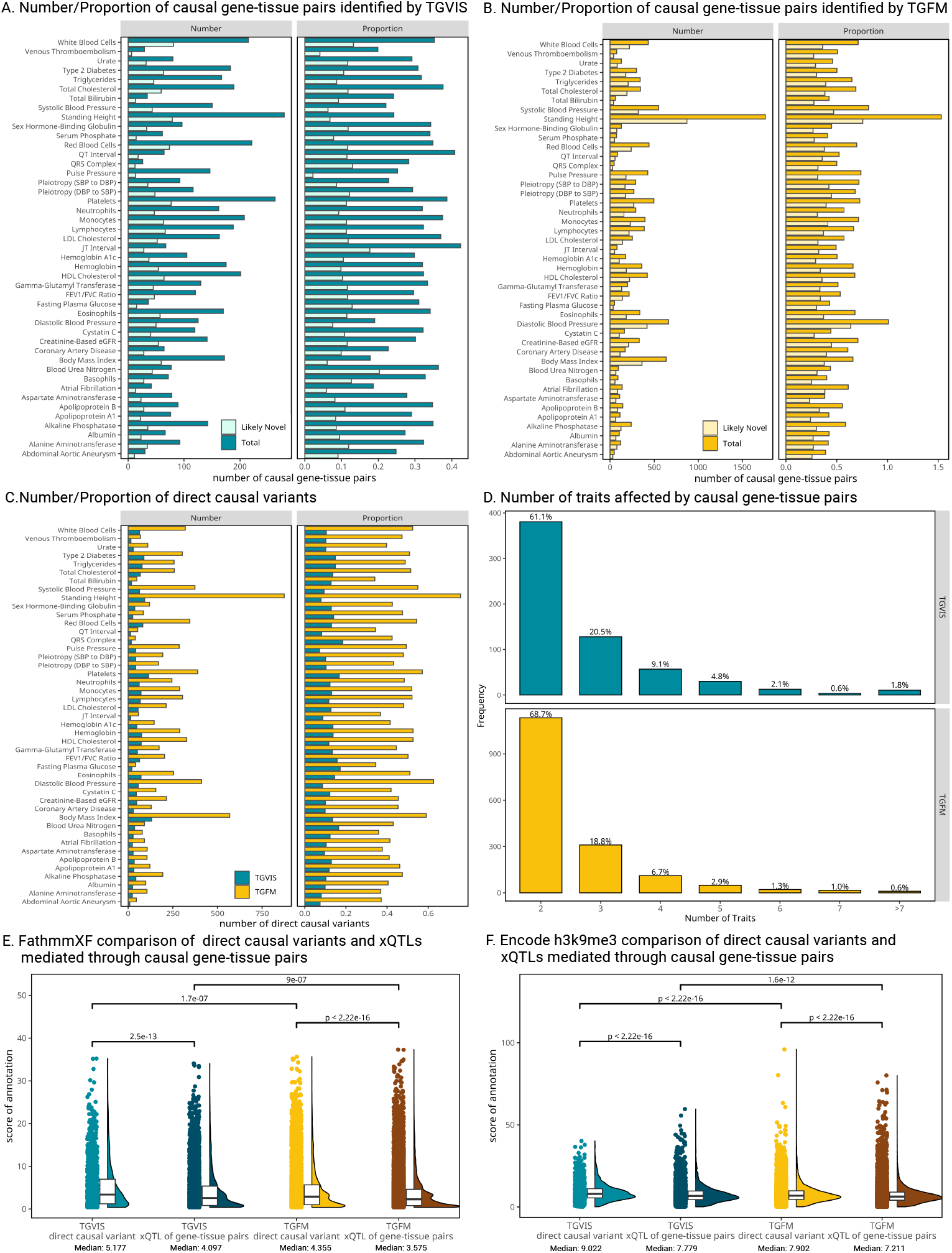
Summary of the identification of causal gene-tissue pairs and direct causal variants. **A-B**: The number and proportion of causal and likely novel causal gene-tissue pairs identified by TGVIS and TGFM, respectively. Likely novel gene-tissue pairs are defined as those not present in the list of significant gene-tissue pairs identified by univariable S-PrediXcan (P < 0.05/20000). The proportion refers to the average number of causal and likely novel causal gene-tissue pairs per locus. **C**: The number and proportion of direct causal variants identified by TGVIS and TGFM. **D**: The distribution of the number of traits affected by causal gene-tissue pairs. **E-F**: The distributions of scores for FathmmXF and Encode H3K9me3Sum annotations. Raincloud plots illustrate four classes: direct causal variants and xQTLs of causal gene-tissue pairs identified by TGVIS and TGFM. Pairwise Wilcoxon signed-rank test P-values are displayed at the top, while medians of annotation scores are shown at the bottom.

We expected that general causal gene-tissue pairs detected by TGVIS and TGFM would likely be included among those identified by univariable TWAS methods such as S-PrediXcan^41^. Surprisingly, among the causal pairs identified by TGVIS, a median of 34.3% were undetected by S-PrediXcan, and this proportion was 60.1% for TGFM (**Figure 3A, Table S22**). For example, TGVIS identified *SCN2A*-Nerve_Tibial as a novel causal gene-tissue pair for 17 traits (**Figure S37**) but missed by TWAS. Both TGVIS and TWAS identified *SCN2A*-Nerve_Tibial for type 2 diabetes. Our findings suggest *SCN2A* may regulate a wide range of metabolic traits. These results indicate that TGVIS not only fine-maps causal genes detected but also uncovers novel genes by modeling multiple tissue-gene pairs simultaneously.

We investigated how many traits can be influenced by a causal gene-tissue pair, reflecting the pleiotropic effect at the gene-tissue level. Among the causal gene-tissue pairs falling in credible sets of size less than 2, 22.4% identified by TGVIS and 16.7% by TGFM exhibit pleiotropic effect (**Figure 3D, Table S7-S8**), indicating that many of these causal genes contribute to shared biological mechanisms across multiple traits.

We further examined whether the direct causal variants and xQTLs mediated by causal gene-tissue pairs differ in functionality using functional annotations^35^ (**Methods)**. Significant differences were observed between these two types of variants identified by either TGVIS or TGFM across multiple annotations (**Table S11**). As shown in **Figure 3E** and **3F**, the direct causal variants generally have higher FathmmXF and h3k9me3 scores than the xQTLs mediated by causal gene-tissue pairs (Wilcoxon signed-rank test, P<2.2E-16), suggesting distinct biological mechanisms for many of these variants.

We observed that multiple eGenes and sGenes often shared the same set of variants as their xQTL, highlighting the importance of making inferences based on credible sets rather than individual variables. Most credible sets consisted of 2 to 4 gene-tissue pairs (60.5%), although some credible sets included more than 10 (11.5%) for TGVIS (**Figure 4A, Table S12**). In comparison, TGFM resulted predominantly featured single gene-tissue pairs (56.0%) and 2 to 4 pairs (41.7%) per credible set (**Figure S18, Table S12**). On the other hand, most of the credible sets only had one xQTL (66.6%), followed by two xQTLs (12.6%) for TGVIS (**Figure 4B, Table S13)**. As for TGFM, these percentages were 26.9% for one xQTL and 24.4% for two xQTLs (**Figure S20, Table S13**). These differences arise because TGFM resampled all xQTLs in the 95% credible sets, typically incorporating more variants, whereas TGVIS applied a stricter criterion for selecting xQTLs (**Methods, Figure S18-S19**).

**Figure 4:**
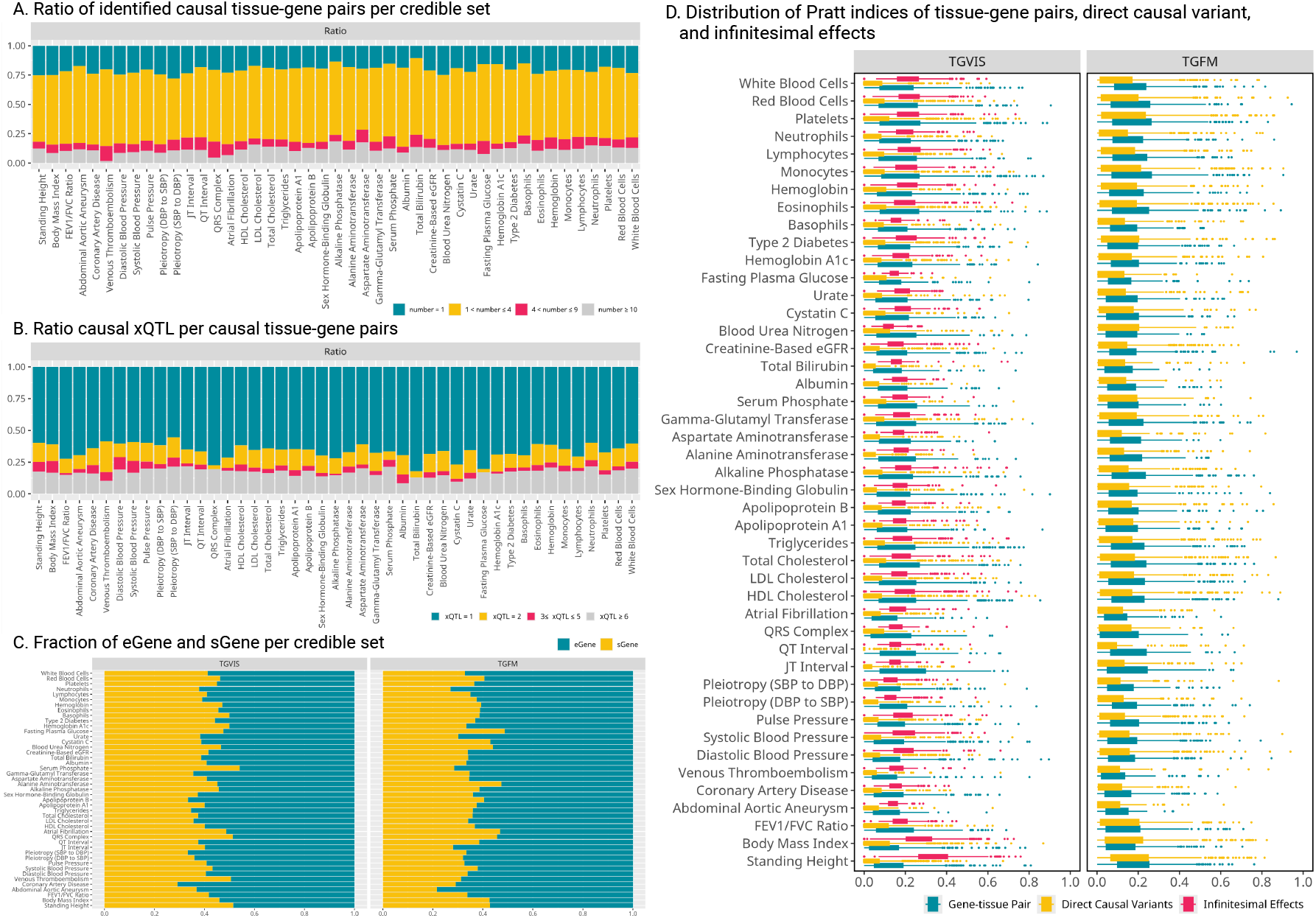
Genetic architecture inferred from the identification of causal gene-tissue pairs and direct causal variants. **A**: The ratio of identified causal gene-tissue pairs per credible set by TVGIS. Different gene-tissue pairs may share the same set of xQTLs, and end in the same credible set. **B**: The ratio of the number of causal eQTLs over the number of sQTLs per causal gene-tissue pair, indicating the distribution of eQTLs and sQTLs contributing to the gene-tissue pairs. **C**: The distribution of eGene and sGene in credible sets identified by TGVIS and TGFM. When a credible set contains multiple gene-tissue pairs, we calculate the proportion of eGenes and sGenes. **D**: The distribution of Pratt Index estimates for different traits, with a comparison between TGVIS and TGFM. In the boxplot, each point represents the Pratt Index of various molecular phenotypes within a single locus.

We investigated the proportions of identified causal eGenes and sGenes for the 45 cardiometabolic traits (**Methods**). TGVIS showed eGenes and sGenes proportions of 58.1% and 41.9%, respectively, while TGFM resulted in 63.5% for eGenes and 36.5% for sGenes (**Figure 4C, Figure S21**). These results align with the proportions observed in the GTEx Consortium (63% *cis*-eQTL vs. 37% *cis*-sQTL)^12^, with TGFM’s proportions being slightly closer. A potential explanation is that TGVIS’ eGenes and sGenes were more likely enriched for causal genes specific to cardiometabolic traits, leading to a slight difference, though this difference is not substantial.

We calculated the Pratt index of gene-tissue pairs, direct causal variants, and infinitesimal effects based on its additive property (**Figure 4D, Table S15**), which helps measure the contributions of these three potentially correlated components (**Methods**). For TGVIS, the median of the Pratt index was 0.161, 0.059, and 0.182 for gene-tissue pairs, direct causal variants, and infinitesimal effects, respectively, with a median sum of the Pratt index of 0.403. In comparison, for TGFM, the median of the Pratt index was 0.145 for gene-tissue pairs and 0.114 for direct causal variants, with a median sum of the Pratt index of 0.262. These results support the existence of widespread infinitesimal effects.

### Major relevant tissue map of cardiometabolic traits

We searched the major relevant tissues by counting their numbers to the causal gene-tissue pairs in credible sets identified by TGVIS and TGFM (**Methods**). We ranked the top relevant tissues according to their contributions and clustered similar traits and tissues based on the similarity of the identified causal gene-tissue pairs (**Figure 5A-5B, Figure S22-S23**). Overall, we observed similar major relevant tissues and clustering patterns using both methods, although there were some notable differences. TGVIS tended to cluster similar traits more closely together than TGFM. For instance, TGVIS grouped all blood pressure traits into close clusters, placing them near coronary artery disease (CAD), whereas TGFM positioned systolic and diastolic blood pressures (SBP and DBP) farther from pulse pressure (PP) and CAD. Similarly, serum lipid traits were clustered together by TGVIS, but not by TGFM. On the other hand, arterial tissues consistently emerged as the major tissue for blood pressure traits and CAD, while heart tissues were the major tissue for the QRS complex, atrial fibrillation, QT interval, and JT interval. Fibroblasts were highlighted as an important tissue for many traits, aligning with recent findings about their role in tissue integrity and chronic inflammation, alongside other tissues such as adipose tissue and liver^42^.

**Figure 5:**
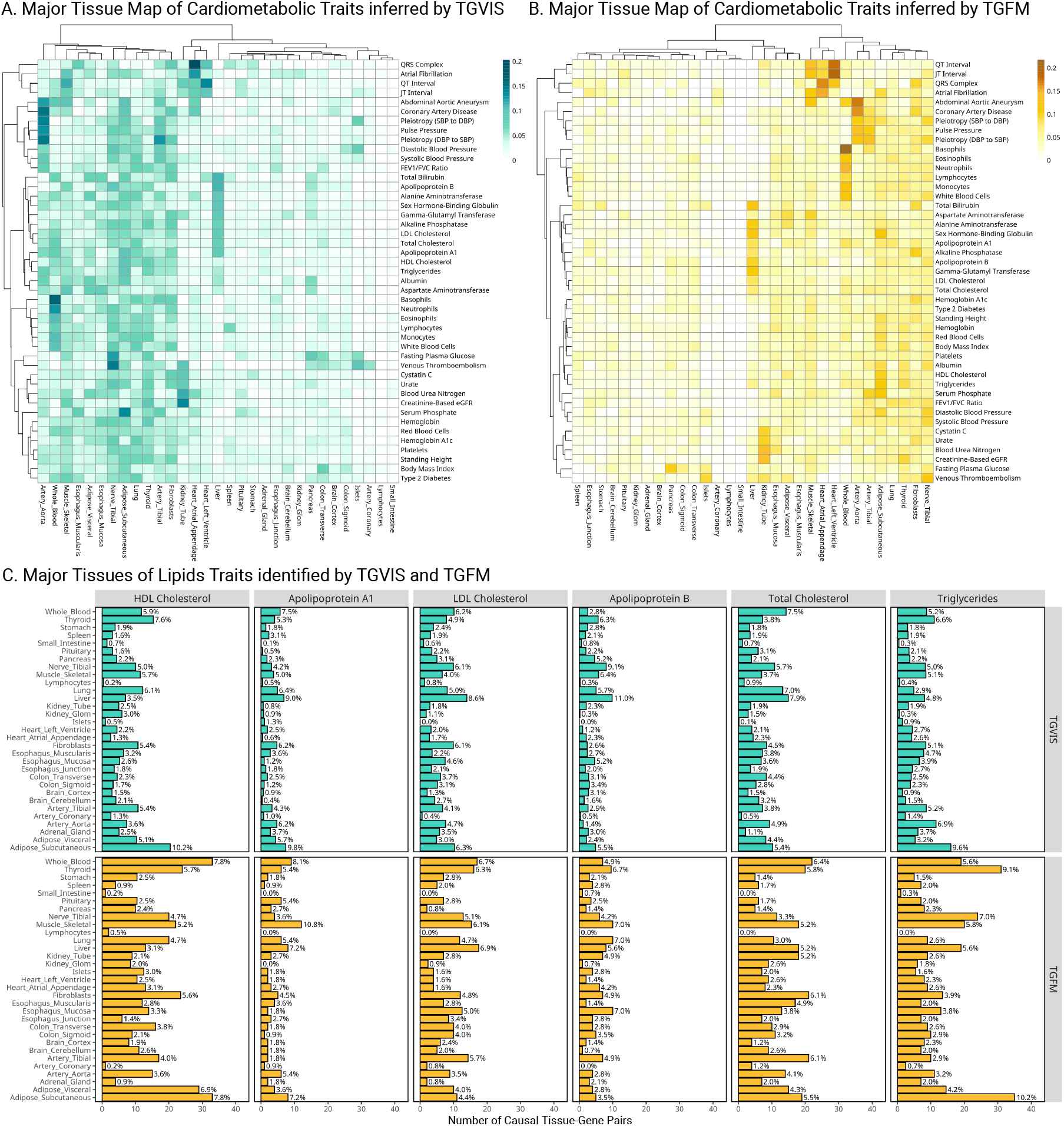
Distribution of major tissues for cardiometabolic traits. **A**: Heatmaps display the major tissues associated with each trait, identified by TGVIS. **B**: Heatmaps display the major tissues associated with each trait, identified by TGFM. The major gene-tissue pairs are cataloged based on stringent criteria (CS-Pratt > 0.15 for TGVIS and PIP > 0.5 for TGFM) and the proportions of major tissues derived from significant gene-tissue pairs for each trait are quantified. Hierarchical clustering is applied to arrange the heatmaps, utilizing the Ward2 method and Euclidean distance. **C**: Major tissues of lipid traits identified by TGVIS and TGFM. This panel shows bar plots detailing the number of causal gene-tissue pairs for various lipid traits, including HDL-C, LDL-C, TC, triglycerides, APOA1, and APOB, as identified by both TGVIS (top) and TGFM (bottom).

We considered several lipids traits, including LDL-C, HDL-C, TC, triglyceride, apolipoprotein A1 (APOA1), and apolipoprotein B (APOB), as examples to illustrate the proportional counts of each tissue identified in the credible sets. For HDL-C and triglycerides, the most relevant tissue was subcutaneous adipose (**Figure 5C**). In contrast, liver tissue was consistently the most relevant tissue for LDL-C, APOB, and TC, despite the small sample size for the liver tissue gene expression data^12^. For APOA1, the two most relevant tissues were the liver and subcutaneous adipose tissue. **Figures S24-S32** display the plots of major tissues for the rest of the traits. Overall, the TGVIS and TGFM produced in general consistent results.

### Evaluation of the identified gene-tissue pairs

To evaluate the accuracy of the prioritization of causal gene-tissue pairs, we first compared the colocalization evidence of the causal credible sets identified by TGVIS and TGFM through Coloc-SuSiE^5^. Since a credible set could include multiple tissue-gene pairs, we defined a colocalization of a credible set in two criteria: (1) the credible set contained at least one gene-tissue pair that is colocalized with the trait; (2) more than 50% of the gene-tissue pairs in the credible set were colocalized with the trait (**Method**). TGVIS had much higher proportions of colocalized credible sets (the median proportions across traits were 93.1% and 77.8% for the two criteria, respectively) than TGFM (the median proportions across traits were both 40.9% for two criteria) (**Figure 6A and Table S16-S17**), suggesting a substantial number of causal tissue-gene pairs identified by TGFM do not have colocalization evidence.

**Figure 6:**
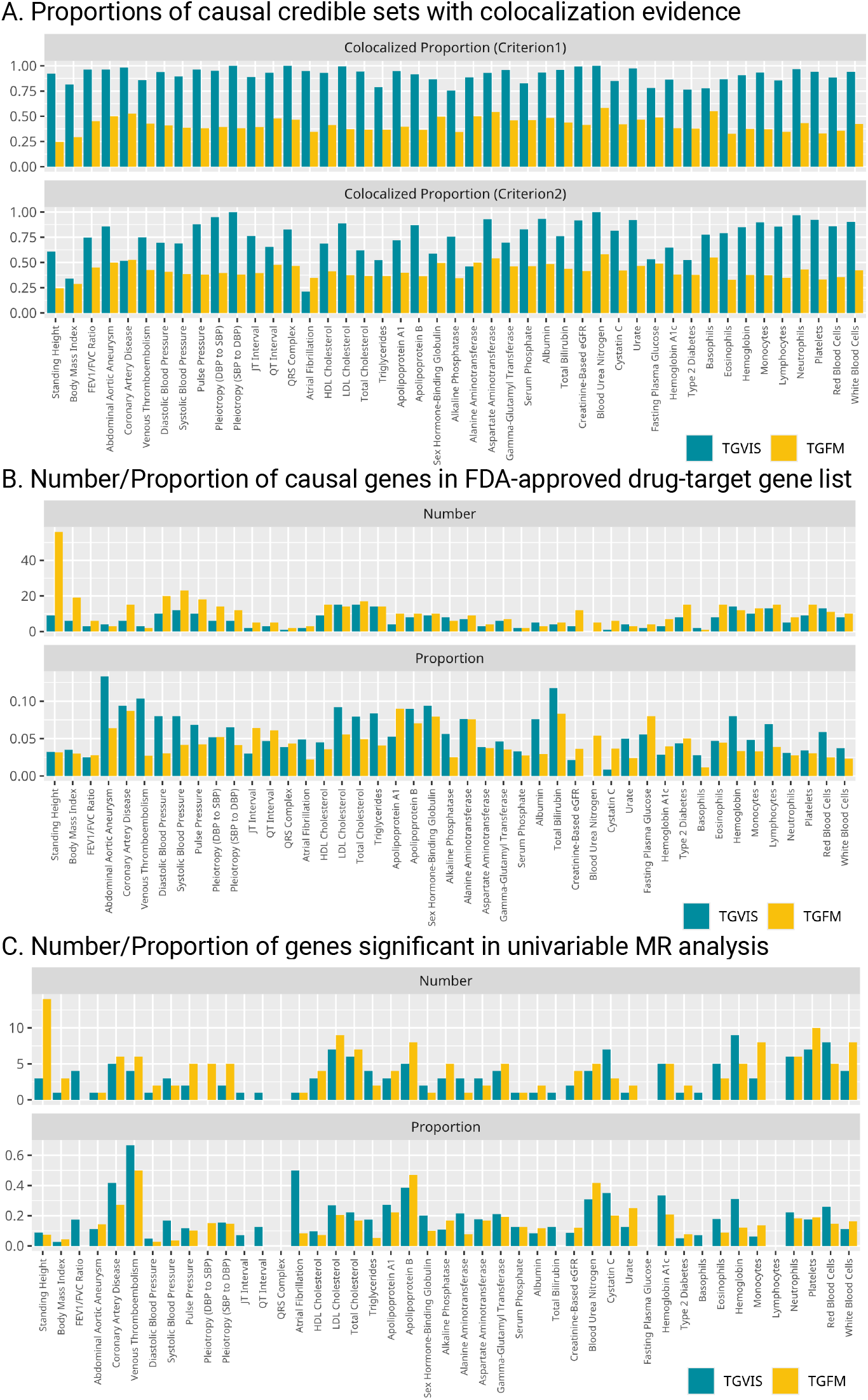
Evaluation of identified gene-tissue pairs. **A**: The colocalized proportions of causal credible sets (under two criteria) yielded by TGVIS and TGFM, respectively. **B**: The numbers and proportions of causal cis-genes in the list of FDA-approved drug-target genes provided by Trajanoska et al., identified by TGVIS (left) and TGFM (right), respectively. **C**: The number of significant pGenes in univariable MR analysis and the ratio of significant pGene in univariable MR analysis divided by significant eGenes/sGenes in eQTL/sQTL analysis.

We next followed the previous analysis strategy^13^ to assess the causal genes for LDL-C identified by TGVIS and TGFM. Precision was evaluated using the 69 known lipid-related genes as the silver standard positive gene set, and nearby genes within a 1MB-radius region as the negative set, as studied by Zhao et al.^13^. We disregarded the tissue part of the identified causal gene-tissue pairs and then calculated how many causal genes were within the lists of sliver and nearby genes. TGVIS demonstrated a precision of 60.0% (9 out of 15), outperforming TGFM, which had a precision of 37.5% (10 out of 28) (**Table S18** and **Figure S33**).

It is reasonable to assume that causal genes are more likely to be druggable targets. We utilized the published list of 6,690 FDA/EMA-approved non-cancer drugs (**Table S1** provided by Trajanoska et al.^36^) to calculate the enrichment of the identified causal genes in the drug list (**Figure 6B** and **Table S19-S20**). Although the number of causal genes identified by TGVIS in the drug-targeted gene list was only 74.3% of that identified by TGFM, the enrichment identified by TGVIS was 1.43 times more than that by TGFM (P = 1.56E-3).

We hypothesized that causal genes detected through eQTLs/sQTLs may be more likely to demonstrate association evidence in protein data. To test this, we conducted univariable MR analysis of protein abundances (pGenes) in blood tissue for genes identified by TGVIS and TGFM, using both *trans-* and *cis*-pQTLs as instrument variables (**Figure 6C** and **Table S21**). On average, 18.1% of pGenes identified by TGVIS showed significant causal evidence, compared to 13.7% of pGenes for TGFM (P = 3.1E-3). However, this proportion is lower than the estimated true positive association rate of 27.8% between predicted *cis*-regulated gene expression and plasma protein abundances^43^. The discrepancy may arise from the fact that pGenes are influenced by widespread *trans*-pQTLs^10^, whereas predicted gene expression is predominantly contributed by *cis*-eQTLs, and its *trans*-regulated effects are much more difficult to detect. This result suggests that eGenes/sGenes and pGenes may represent distinct biological processes related to complex traits^43^.

### Fine-mapping of causal gene-tissue pairs and variants in GWAS loci

We exemplified four loci associated with LDL-C, CAD, and BMI. The first locus contains the *PCSK9* gene for LDL-C (**Figure 7A**). TGVIS identified three 95% credible sets, including *PCSK9-*Whole_Blood and two direct causal variants *rs11591147* and *rs11206517* (**Figure 7B**). After applying the threshold of CS-Pratt > 0.15, *PCSK9-*Whole_Blood (CS-Pratt = 0.17) and *rs11591147* (CS-Pratt = 0.492) remained. In contrast, TGFM identified nine gene-tissue pairs and direct causal variants with PIPs > 0.5 (**Figure 7C**), including the *MROH7*-Esophageal_Mucosa which has no clear connection to the biology of LDL-C. Applying the CS-Pratt threshold, *PCSK9-*Whole_Blood (CS-Pratt = 0.204) and *rs11591147* (CS-Pratt = 0.524) remained, consistent with the results yielded by TGVIS. This example demonstrates how TGVIS reduces false positives by modeling infinitesimal effects and applies the Pratt Index as an additional criterion.

**Figure 7:**
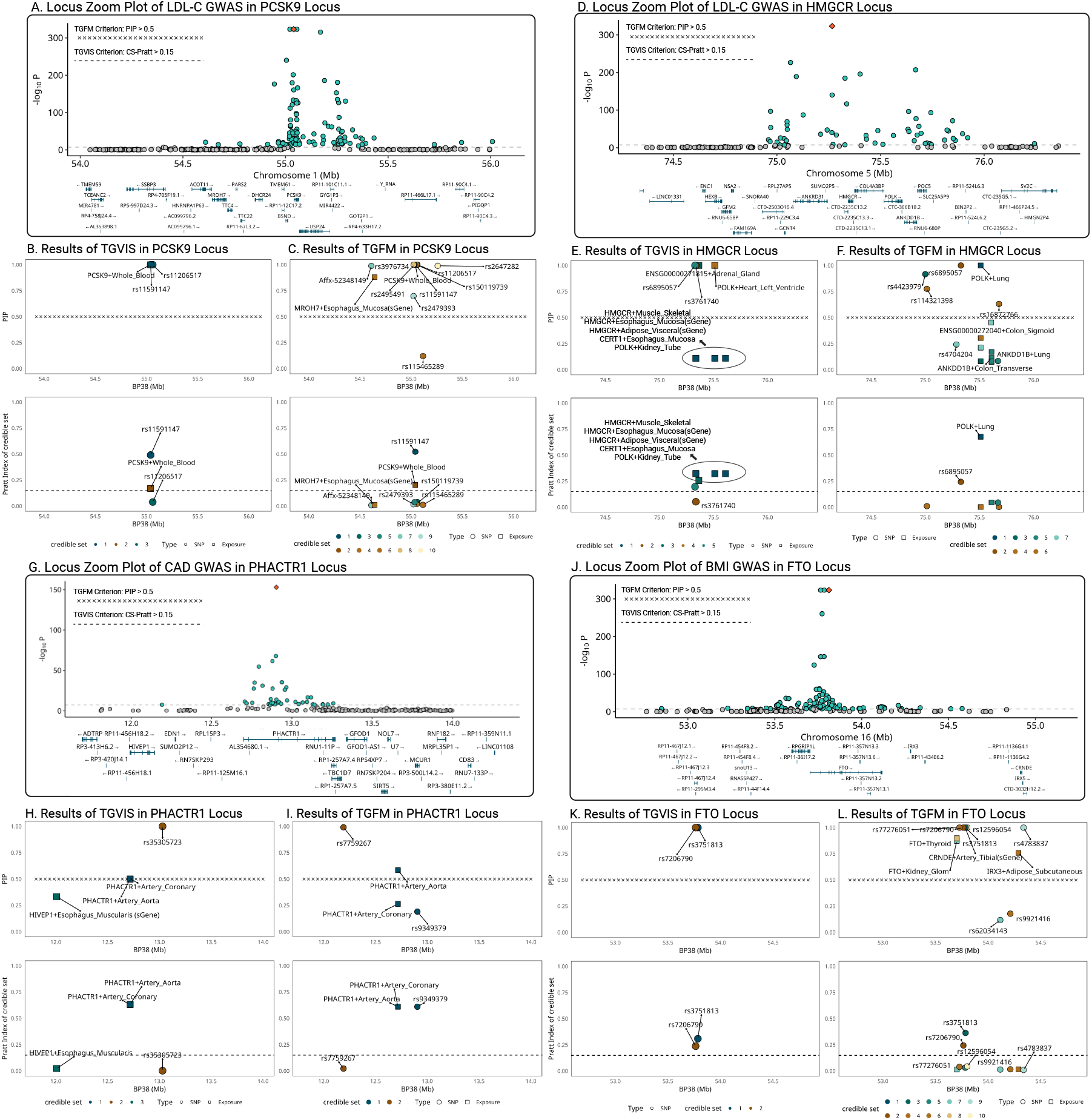
Locuszoom plots comparing the results of TGVIS and TGFM. **A-C**: LDL-C (*PCSK9* locus). **D-F**: LDL-C (*HMGCR* locus). **G-I**: CAD (*PHACTR1* locus), **K-L**: BMI (*FTO* locus). For each locus, we included three plots: (1) the GWAS of the trait, (2) the PIP of gene-tissue pairs and direct causal variants identified by the TGVIS and TGFM, and (3) the Pratt index of corresponding gene-tissue pairs and variants. For TGVIS, causality is determined by (1) the variables are in a 95% credible set and (2) the Pratt index of this credible set is larger 0.15. For TGFM, the causality is determined by (1) the individual PIP is larger than 0.5.

The second locus contains the *HMGCR* gene causal^44^ to LDL-C (**Figure 7D**). TGVIS identified five 95% credible sets (**Figure 7E**). The first credible set (the darkest green) includes 9 gene-tissue pairs, such as *HMGCR-*Muscle_Skeletal and five of its sGenes in esophagus mucosa, nerve tibial, fibroblasts, and adipose visceral, all sharing the same xQTL *rs2112653*. When we applied the threshold of individual PIP > 0.5, none of the pairs in this credible set were selected although they were all in a 95% credible set. However, this set had the highest CS-Pratt of 0.322 among the five 95% credible sets. Conversely, TGFM identified *POLK-*Lung (CS-Pratt = 0.684) but missed the crucial *HMGCR* gene (**Figure 7F**). This is likely a false discovery, as *HMGCR* inhibitor is a key component of statins, which works by inhibiting HMG-CoA reductase and thus reduces LDL-C in the blood^44^.

In the third example, we focused on the *PHACTR1* locus related to CAD (**Figure 7G**). Both TGVIS and TGFM identified a major credible set at this locus, including *PHACTR1-* Artery_Coronary and *PHACTR1-*Artery_Aorta, with CS-Pratt values of 0.632 and 0.612, respectively (**Figure 7H-7I**). In TGVIS, the individual PIPs of them were both 0.5 and the cumulative PIP for this credible set was 1. In contrast, TGFM resampled both the eQTL effect estimates and the PIPs (**Methods**), resulting in a higher individual PIP and Pratt index for *PHACTR1-*Artery_Aorta (PIP = 0.597, Pratt = 0.472) than *PHACTR1-*Artery_Coronary (PIP = 0.222, Pratt = 0.053). However, as noted by Strober et al.^14^, this resampling process tends to favor gene-tissue pairs with larger sample sizes, which may explain the exclusion of *PHACTR1-* Artery_Coronary. TGVIS adheres to the original interpretation of SuSiE that the variables within a credible set cannot be distinguished from the available data.

The final exemplary locus is the *FTO* locus associated with BMI (**Figure 7J**). TGVIS identified only two direct causal variants *rs7206790* and *rs3751813* and did not find any gene-tissue pairs at this locus (**Figure 6K**). In contrast, TGFM identified four gene-tissue pairs: *FTO_*Kidney_Glomerulus, *FTO_*Thyroid, *FTO_*Artery Tibial, and *IRX3*-Adipose_Subcutaneous, and five direct causal variants (**Figure 7L)**. However, the associations between obesity and the expression of the *FTO* gene in the kidney glomerulus, thyroid, and tibial artery are not well-established in the literature. After applying the Pratt index threshold, only two direct causal variants *rs7206790* and *rs3751813*, remained, consistent with the result from the TGVIS. When we reduced the locus radius from 1MB to 500KB and re-ran the analysis, both TGVIS and TGFM identified the sGene of *FTO-*Pancreas as causal, with CS-Pratt values of 0.345 and 0.407, respectively (**Figure S35**). The sQTLs of this *FTO* sGene are *rs7206790* and *rs11642841*, which have been reported by Xu et al.^45^. This example suggests that when applying multivariable TWAS methods, the size of a *cis*-region can be sensitive and need to be calibrated.

## Discussion

In this report, we developed TGVIS to identify causal gene-tissue pairs and direct causal variants in loci identified through GWAS by integrating xQTL summary statistics. Compared to cTWAS^13^ and TGFM^14^, TGVIS not only analyzes multiple tissue-specific xQTL summary data simultaneously to pinpoint causal gene-tissue pairs and direct causal variants, but also models the widespread presence of infinitesimal effects underlying polygenic traits to reduce false discovery rates in detecting causal molecular phenotypes^17^. In addition, TGVIS quantifies the importance of a causal variable by the Pratt index, which has been well established in statistics and has recently been applied to estimate the gene-by-environment contribution^23^. Through simulations, we demonstrated that under the presence of infinitesimal effects, TGVIS has lower MSE and higher TPR and TNR compared to both cTWAS and TGFM (**Figure 2**). In real data analysis, TGVIS outperformed TGFM in the following four aspects: (1) identifying more interpretable major trait-relevant tissues (**Figure 5**); (2) resulting in a higher proportion of colocalized causal credible sets (93.1% vs 40.9%, **Figure 6A**); (3) achieving notably higher precision in the ‘silver standard’ sets of lipids (60.0% vs 37.5%, **Table S15**); and (4) demonstrating significantly greater enrichment evidence based on druggable genes (1.43 times, **Figure 6B**) and causal proteins (1.31 times, **Figure 6C**).

Our analysis of 45 cardiometabolic traits provides several key insights. First, we identified a median of 34.3% causal gene-tissue pairs that were missed in univariable TWAS analysis, suggesting that TGVIS is able to identify novel genes besides fine-mapping the genes detected by conventional TWAS (**Figure 3A**), representing a significant advance in TWAS. Second, we observed that infinitesimal effects can make a substantial contribution to local genetic variation of traits besides the gene-tissue pairs and direct variant (**Figure 4D**), which is consistent with recent studies^17,21,46^. Beyond underlying biological mechanisms such as the polygenicity of human complex traits, the emergence of infinitesimal effects may also be attributed to nonbiological factors, particularly estimation errors in the LD matrix, xQTL effect sizes, and trait GWAS imputation (**Method**). Both empirical observations and theoretical investigation underscore the importance of including infinitesimal effects in future genetic researches and methodological developments. Third, our study indicates that a significant proportion of causal gene-tissue pairs (22.4%) exhibit pleiotropic effects at the gene-tissue level, suggesting shared biological mechanisms across multiple traits (**Figure 3D, Table S7-S8**). Fourth, our findings suggest that for most traits, only a limited number of relevant major tissues are involved (**Figure 4A**), implying that concentrating multi-omics data analyses on these relevant major tissues can be more powerful and efficient, as well as it can make the findings more biologically interpretable. For example, when the analysis is focused on the four major blood-pressure-relevant tissues, i.e., adrenal gland, artery, heart, and kidney, it leads to the identifications of more causal gene-tissue pairs, with an increased Pratt index for blood pressure traits^47^. Fifth, our results indicate that only 18.1% of causal genes from eQTL/sQTL analyses also show causal evidence in univariable MR using pQTL summary data (**Figure 6C**), suggesting that gene expressions and protein abundance represent distinct biological processes in complex traits^43^. Finally, we identified an average of 0.304 causal gene-tissue pairs per locus and failed to identify any causal gene-tissue pairs in many GWAS loci (**Figure 3A**), which is consistent with the recent study showing that the GWAS and eQTL studies are systematically biased toward different types of variants^4^. Interestingly, the eQTLs/sQTLs of causal gene-tissue pairs and direct causal variants have substantially different functional annotations (**Figure 3E-3F, Table S11**), warranting further investigation.

Our study has some limitations. Due to the data and computational constraints, we only analyzed genes using *cis*-eQTL/sQTL summary statistics, limiting our ability to distinguish between genes that share *cis*-eQTLs/sQTLs, which may lead to false discoveries. This issue could potentially be addressed by incorporating *trans*-eQTLs/sQTLs, although this would require much larger sample sizes. In addition, we observed that a credible set often contains 2-4 gene-tissue pairs (**Figure 3C**), likely due to the small sample size in the GTEx data, which results in only 1 or 2 eQTL/sQTL for most gene-tissue pairs (**Figure 4B**). In other words, while TGVIS was able to narrow down to a range of causal gene-tissue pairs, it could not always pinpoint the exact causal pair(s) in some loci. Incorporating external information, such as colocalization evidence with TGVIS, may aid in distinguishing these pairs^48^. Furthermore, our eQTL/sQTL analysis relies on bulk tissue expression data, which may limit our ability to identify cell-type-specific causal genes^49^. For example, recent studies increasingly suggest that *FTO* may not be the causal gene for BMI; instead, experimental evidence indicates that *IRX3* and *IRX5* are the causal genes^50^.

However, the causality of *IRX3* and *IRX5* was observed in experiments using preadipocytes, rather than bulk subcutaneous adipose tissue, which may explain why TGVIS failed to identify these genes (**Figure S36**). Moreover, we used the Pratt index^23^ to rank the importance of variables, but it has inherent statistical limitations^23^. In simulations, the Pratt index slightly underestimates the true contribution, although this underestimation becomes negligible as the sample size increases (**Figure S1-S8**). In real data analysis, we used an empirical cutoff learned by K-means (CS-Pratt = 0.15) to extract important causal variables, which gives us higher precision but may have potentially hindered the discovery of causal gene-tissue pairs with small to moderate causal effects. Last, as suggested in previous studies^14^, the inference of causality based on statistical methods comes with a caveat, assuming no model misspecification and no potential causal elements are missing from the model.

In summary, our developed TGVIS and accompany software pipeline provide a valuable tool in fine-mapping and interpreting GWAS findings.

## Methods

### Multivariable TWAS model

The causal diagram shown in **Figure 1A** can be described by the following multivariable TWAS model:

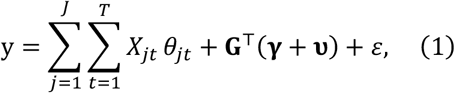

where *y* is a trait, *X*_*jt*_ is the *jt*^th^ gene-tissue pair, **G** = (*G*_1,_ …, *G*_*M*_)^⊤^ is an (*M* × 1) vector of genetic variants in the *cis*-region, **θ** = (θ_11_, …, θ_*JT*_)^⊤^ is an (*JT* × 1) vector of causal effects with θ_*jt*_ being the causal effect of the (*j, t*)th tissue-gene pair, **γ** = (γ_1,_ …, γ_*M*_)^⊤^ is an (*M* × 1) vector of direct causal effects, **υ** = (υ_1,_ …, υ_*M*_)^⊤^ is an (*M* × 1) vector of infinitesimal effects, and ϵ is the random error. Let **β**_*jt*_ = (β_*jt*1_, …, β_*jtM*_) is an (*M* × 1) vector of the *cis*-eQTL effects of *JT* tissue-gene pairs. Then we have

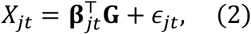

where ϵ_*jt*_ is the noise of the *jt*^th^ gene-tissue pair. The reduced form of (1) is then given by:

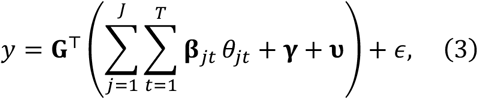

where mathematically 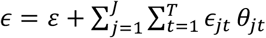.

An alternative version of (1) based on summarized statistics^51^ is:

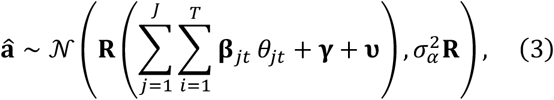

where â = (â_1,_ …, â_*M*_)^⊤^ represents the GWAS effects of the outcome, **R** is an (*M* × *M*) LD matrix of the *M* variants, and 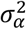 is the variance of this model. The eQTL effect vector **β**_*jt*_ follows the model based on summarized statistics below:

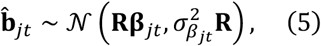

Where 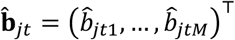 represents the marginal *cis*-eQTL effect estimates for the *jt*^th^ tissue-gene pair, and 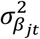 denotes the variance of this model.

To resolve this curse of dimensionality, we utilized the three sparsity conditions that are commonly assumed in current fine-mapping methods^16,19^: (SP1) one or small number of variants causally contribute to tissue or cell-type specific gene expression^12^; (SP2) one or small number of gene-tissue pairs causally contribute to the trait^13,14^; (SP3) one or small number of direct causal variants exist with relatively large effect sizes^13,14^. In terms of statistical model: SP1 corresponds to **β**_*jt*_ being sparse for all *j* and *t*; SP2 corresponds to **θ** being sparse; SP3 corresponds to **γ** being sparse. In addition, we incorporated that variants can have infinitesimal effects: **υ** is normally distributed with a mean 0 and a small, unknown variance ^17^. To our best knowledge, infinitesimal effects have not been modeled in current multivariable TWAS methods.

### Estimation of *cis*-regulatory effect

TGVIS first applies SuSiE^18^ to estimate the non-zero eQTL effect for each gene-tissue pair, based on the fine-mapping model (Equation 5). Specifically, we set *L* = 3 for each pair and determined the non-zero *cis*-regulatory effects based on two criteria: (1) if they are within any 95% credible set and their PIPs exceeds 0.25, and (2) if their individual PIPs are greater than 0.5. The rationale behind this approach is that SuSiE’s 95% credible set can sometimes include too many weakly correlated variants (even after removing highly correlated ones using LD clumping), leading to low PIPs for each variant. Therefore, we used a moderate threshold to filter out credible sets with too many variants. Additionally, due to the low power of detection, the maximum PIP of credible sets might fall below 0.95, so we retained variants with individual PIPs greater than 0.5. Since a locus often contains over 10,000 gene-tissue pairs (mostly sGenes), dynamically selecting using BIC would be computationally burdensome. Additionally, with GTEx sample sizes under 200, only 1-2 gene-tissue pairs can be identified for most gene-tissue pairs. Therefore, we choose to fix *L* = 3.

### Joint modelling of causal tissue-gene pairs, direct causal variants, and infinitesimal effects using profile likelihood

TGVIS estimates **θ, γ**, and **υ** using a profile likelihood approach. Given the estimate **υ**^(*s*)^ from the *s*th iteration, we considered the following fine-mapping model:

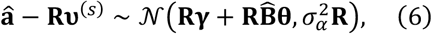

where 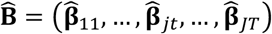 is an *M* × *JT* matrix consisting of estimated *cis*-regulatory effects. To update **γ** and **θ** simultaneously, we applied the same scheme as cTWAS and TGFM, using the function susie_rss(·). The input z-score vector is computed as:

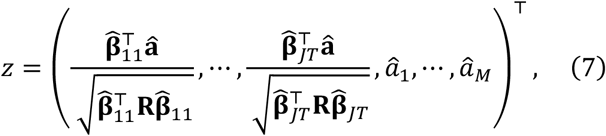

and the other elements of input correlation matrix are computed as:

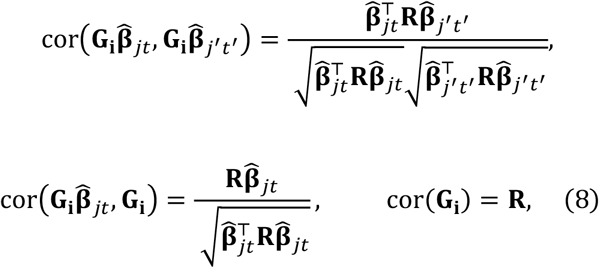

The outputs are denoted as **γ**^(s+1)^ and **θ**^(***s***+1)^.

Next, we consider the following model:

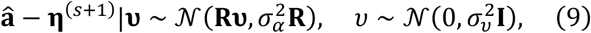

Where 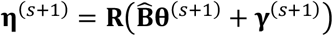. The penalized quasi-likelihood (PQL) of **υ** is

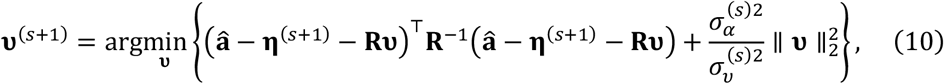

which results in

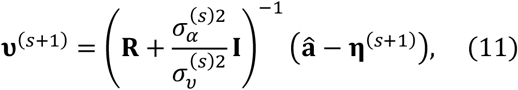

where 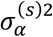 is the current variance estimate. The variance 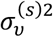 is updated by REML:

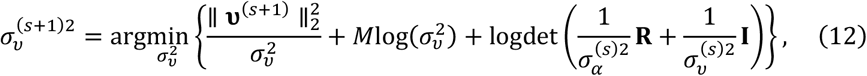

which simplifies to

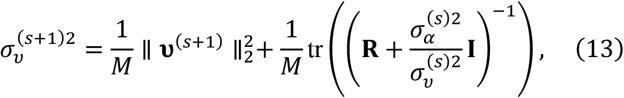

where *M* is the number of variants. We replace 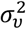 in the last term by its current estimate 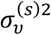 to obtain a close-form expression. Note that in Equation (13), 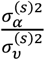 is usually replaced by 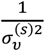 to avoid non-identifiability issues^22^.

When the profile likelihood converges, TGVIS estimates 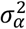 as follows:

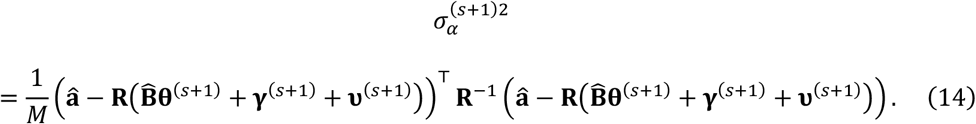

### Bayesian Information Criterion for Summary Data

Based on Equation (3), we define the BIC for summary data:

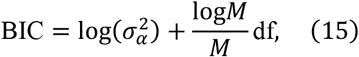

where *M* is the number of IVs and df is the degree of freedom of the model^52^. In practice, 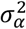 is replaced by its empirical estimate 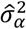, and df is the sum of non-zero causal effect estimates and non-zero direct causal variant estimates. Our default setting assumes *L* can be 2,3,4,5,6,7, or 8 and uses BIC to select the optimal *L* among them. We found that when considering the infinitesimal effect, it tends to capture variants with very small effects that SuSiE does not identify, making it rare for *L* to exceed 8 in practice.

### Pratt index

We use the Pratt index to assess the contribution of a gene-tissue pair. For a general linear model: 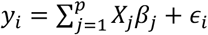, the Pratt index of *x*_*ij*_ is defined as *V*_*j*_ = β_*j*_ × *b*_*j*_, where *b*_*j*_ = cov(*y, X*_*j*_). This definition assumes standardization where E(*y*) = E(*X*_*j*_) = 0 and var(*y*) = var(*X*_*j*_) = 1, 1 ≤ *j* ≤ *p*. The Pratt index measures the contribution of a variable in a linear model because 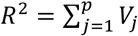 where 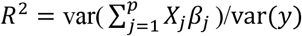. In practice, the Pratt index can be estimated by 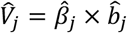, where 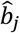 is the sample correlation between *X*_*j*_ and *y*.

The proportion of variance explained (PVE) is defined as 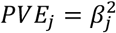, assuming that all variables are standardized. The Pratt index has two key advantages over PVE: (1) Pratt indices are additive across variables, and (2) the sum of Pratt indices is the total trait variance explained by covariates. In contrast, PVE lacks these advantages.

### Pratt Index in TGVIS

Wee show how to yield the Pratt index *V*_*jt*_ in practice. We first estimate the marginal correlation:

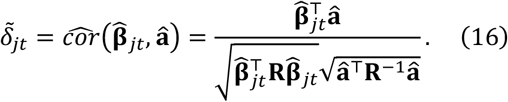

As for the causal effect estimate 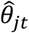, we apply the transformation

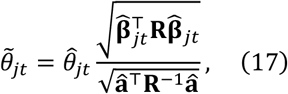

since the Pratt index requires the covariates and trait are all standardized. Thus, the Pratt index of the (*j, t*)th gene-tissue pair is

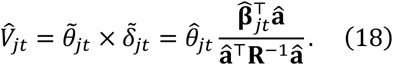

Since Pratt indices are additive, the Pratt index of a credible set is simply calculated as

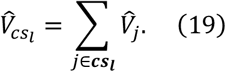

Note that the Pratt index is only comparable within the same locus, as it represents the ratio of the variance explained by the variable to the total variance of the trait.

It is worth comparing the gene-tissue pair, direct causal variant, and infinitesimal effect contributions at a locus. To simplify the estimation, we consider the linear predictors of all gene-tissue pairs and pleiotropy:

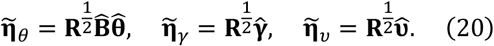

and 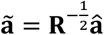, where 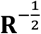 is specified to remove the correlations of 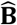 and â. Then, the Pratt indices for the gene-tissue pairs, direct causal variants, and infinitesimal effects are

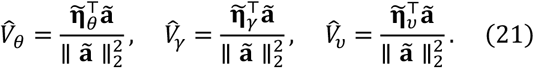

### Threshold of Pratt index

We used the empirical data to determine the threshold for Pratt index to enhance the precision of causal selection. Specifically, we employed K-means clustering with clusters to group the CS-Pratt indices of all gene-tissue pairs and direct variants identified by TGVIS within the 95% credible sets. We hypothesize that one cluster contains credible sets with smaller CS-Pratt values, which are more likely to include falsely causal variables. Interestingly, regardless of whether we focus on gene-tissue pairs, direct causal variants, or both, the minimum value in the cluster with the larger centroid consistently remains at 0.15 (**Figure S34**). Consequently, we set the threshold at CS-Pratt = 0.15 to prioritize the gene-tissue pairs and direct causal variants identified by TGVIS, considering variables with CS-Pratt > 0.15 to have a higher likelihood of being true causal.

### Score test of variance of infinitesimal effects

In implementation, dynamically determining whether to consider the infinitesimal effect is a clever empirical measure. Therefore, we apply the score test of the variance of the random effect in the linear mixed model to test whether the variance of the infinitesimal effect is zero. Specifically, we consider the following hypothesis testing problem:

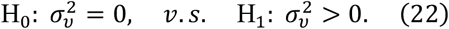

The testing statistics of this hypothesis test is constructed according to Zhang and Lin^53^. Let 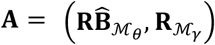 and 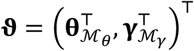. When 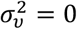 and 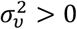, the covariance matrix of 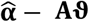 are

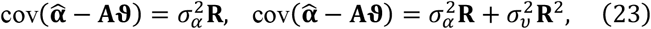

respectively. Similar to estimating σ_υ_, we replace 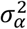 by 1 to avoid non-identifiability. The score described in Zhang and Lin^53^ defined the following three statistics:

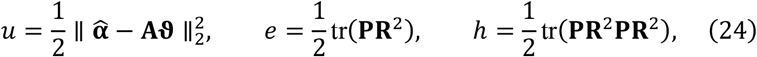

where **P** = **R**^™1^ ™ **R**^™1^**A**(**A**^⊤^**R**^™1^**A**)^™1^**A**^⊤^**R**^™1^. Under the null hypothesis, 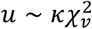 where κ = *h*/(2*e*) and *v* = 2*e*^2^/*h*. If the null hypothesis is accepted, we enforce **υ**^(*s*+1)^ = 0.

### Potential reasons leading to Infinitesimal effects

Here we listed four possible reasons that can lead an infinitesimal effect. First, it has been gradually understood that even within the same ethnic group, such as the European population, different subgroups may have different genetic architectures, leading to different LD structures. Therefore, it is natural to suspect that the LD structures of populations in the GTEx consortium and those in traits GWAS differ, which results in

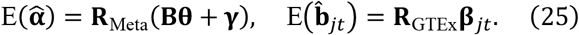

When we try to estimate **β**_*jt*_ using **R**_Meta_, then 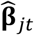 is biased to **β**_*jt*_, which generates infinitesimal effect 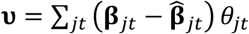. It should be noted that the small sample size in the GTEx consortium can also cause biased eQTL effect estimates, resulting in the appearance of infinitesimal effects. There are other possible sources raising infinitesimal effects, such as (2) the estimation errors of LD matrix, (3) the imputation errors of outcome GWAS effect sizes, and (4) the absence of direct causal variants of outcome, which are shown in **Supplementary Materials**.

### cTWAS and TGFM programs

For cTWAS, since its software is designed to be user-friendly to practical projects, it involves complex settings that are not ideal for simulations, such as requiring a reference panel in BED format and a .db file of eQTL fine-mapping data. Therefore, we directly utilize the principles of cTWAS to develop an R function that employs SuSiE for the first-stage selection of *cis*-regulatory effects and the second-stage selection of causal and horizontally pleiotropic effects.

At the time of writing this paper, the TGFM software has not yet been released. Therefore, we did not consider the first step of cTWAS to estimate two universal prior parameters using the EM algorithm across all loci in the genome. Instead, we restrict cTWAS simulations to a single locus. In addition, we applied the following setting for cTWAS, TGFM and TGVIS: for the prior weight π in SuSiE, we applied π = *p*^™1^ for gene-tissue pairs and π = *M*^™1^ for variants, where *p* represents the number of gene-tissue pairs and *M* the number of variants.

To improve computational efficiency, we applied a slightly different resampling scheme compared the original TGFM. Specifically, we first resampled the eQTL effects from the posterior for 25 times, calculated their mean as 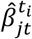 and used these means to estimate 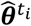 and 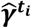. This procedure was repeated 100 times, recording the estimates and PIP for each iteration. We then compute the mean of *t*_1_ × *t*_2_ resampled eQTL effects, 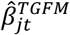, and estimate the empirical 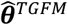 and 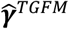. The PIPs of 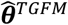 and 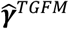 were taken as the empirical PIPs given by SuSiE in each resampling iteration. Finally, we recorded the credible sets of variables from the final step and calculated the PIPs and Pratt indices of credible sets by summing the individual PIPs and Pratt indices of variables within each credible set.

### Simulation Settings

We simulated 20 genes across 5 tissues, resulting in *p* = 100 gene-tissue pairs. Correlations were simulated both within and between genes across tissue. The first and last gene-tissue pairs were designated as causal, with effect sizes of θ_1_ = 1 and θ_100_ = ™1, respectively. The total number of variants was *M* = 400, with only 1,2,3 or 4 of them being eQTLs with non-zero effects for each gene-tissue pair, while the remaining variants were associated with the trait due to LD. We set 4 different sample sizes for the eQTL data (*n*_*eQTL*_ = 100, 200, 400, 800) and a fixed trait GWAS sample size *n*_*trait*_ = 0.5M. Infinitesimal effect were generated from a normal distribution, and gene-tissue pairs, direct causal variants, and infinitesimal effects together were set to explain the trait heritability. For example, when only gene-tissue pairs and infinitesimal effects are present, they each explain 50% of the local heritability for the outcome. When all three are present, each explains 33% of the local heritability for the outcome. The detailed settings, along with corresponding R codes, are provided in Section 2 of the **Supplementary Materials**.

### GWAS summary data

We conducted a meta-analysis on a subset of the 45 metabolic and cardiovascular traits. The publicly available data for these traits are listed in **Table S1**, while the MVP GWAS summary statistics can be accessed through dbGAP under accession number phs001672.v7.p1. For the pleiotropy traits of SBP and DBP, we applied the approach developed in Zhu et al.^32^ using the most recent GWAS summary statistics of SBP and DBP. To perform the meta-analysis, we used METAL^54^. We performed the meta-analysis on the Z-scores, weighting by the sample sizes of the meta-analysis datasets. For binary trait, we always use the effective sample size *n*_eff_. We used CHR:BP (in GRCH37) as the identifier.

### EQTL summary data

We utilized bulk eQTL and sQTL summary statistics from 28 tissues provided by GTEx^12^ (with sample size N ranging from 34.4 (Lymphocytes) to 179.5 (Muscle Skeletal)), as well as additional eQTL summary statistics from tubulointerstitial^33^ (N=311), kidney glomerular^33^ (N=240), and islet^34^ (N=420) tissues (**Table S2**).

### Linkage Disequilibrium Reference Panel

Our study used variants from the UKBB project conducted by Neale’s lab, which initially includes 13 million SNPs. We selected approximately 9.3 million SNPs with a minor allele frequency greater than 0.01 for our analysis. We also identified the top 9,620 unrelated individuals from approximately 500,000 individuals in the UKBB (Field ID: 22828), consisting of 5,205 females and 4,475 males. Data from these 9.3 million SNPs were extracted for these individuals to construct our LD reference panel.

### Clumping and Thresholding

We restricted the studied regions to those within 1MB of the genome-wide significant loci for these traits. These loci were identified using the clumping and thresholding (C+T) method in PLINK^55^: --clump-kb 1000, --clump-p1 5E-8, --clump-p2 5E-8, and --clump-r2 0.01.

We recommend using C+T to filter out variants in high LD, which prevents the inclusion of numerous highly correlated or redundant variants in the analysis, which can unnecessarily complicate the model and result in multiple credible sets consisting of these variants. We evaluated the minimum P-value of each variant across gene-tissue pairs in eQTL/sQTL data. In PLINK, we applied the C+T with the following parameters: --clump-kb 1000, --clump-p1 1E-5, --clump-p2 1E-5, and --clump-r2 0.5. Given that the true causal variant for a trait might not be included in the eQTL/sQTL variants, we combined these variants from outcome GWAS satisfying P < 5E-8 and r^2^<0.5.

### Removing gene-tissue pairs based on significance in S-Predixcan

We used the minimum P-value from S-Predixcan and a modifier accounting for direct causal variants (**Supplementary Materials**) to exclude eGenes/sGene with P > 0.05. These weak filters will eliminate redundant gene-tissue pairs, thereby reducing the model’s dimensionality. Since our goal is fine-mapping the causal gene-tissue pairs and identifying direct causal variants on the GWAS loci with significant signals, it will not induce a winner’s curse.

### Searching causal gene-tissue pairs missed by univariable TWAS

We compared the causal gene-tissue pairs identified by TGVIS and TGFM with the significant gene-tissue pairs identified by S-PrediXcan. We considered genes with P < 0.05/20,000 as significant gene-tissue pairs in tissue specific S-PrediXcan analysis. We did not adjust for number of tissues. We then searched the gene-tissue pairs identified by TGVIS or TGFM but missed by S-PrediXcan.

### Obtaining annotation scores from FAVOR and performing differential annotation tests

We combined the direct causal variants and xQTLs of causal gene-tissue pairs identified by TGVIS or TGFM across all 45 traits into two separate files and uploaded them to the FAVOR online platform^35^ to obtain annotation scores for these variants. We performed Wilcoxon signed-rank test with both “less” and “greater” as alternative hypotheses for determining the direction of shift location and calculated corresponding P-values. We used the R package FDREstimation to convert the P-values to FDR Q-values using the Benjamini–Yekutieli (BY) procedure. Annotations with Q-values less than 0.05 were considered to have significantly different scores.

### Mapping major trait relevant tissues

For TGVIS, a 95% credible set often includes multiple gene-tissue pairs. In such cases, we calculated the proportion of each tissue appearing among these pairs, allowing the number of tissues in a causal credible set of gene-tissue pairs to be fractional. For TGFM, we first removed the gene-tissue pairs with PIPs < 0.5, and then applied the same procedure to map the dominant tissues.

### Enrichment of identified causal genes in lipids silver gene list and druggable gene database

We applied the following strategy to map silver genes. First, we checked each credible set to see if any genes are part of the silver gene list; if so, we counted 1. If no silver gene was present, we then checked if any genes in the credible set were among the nearby genes; if so, we also counted 1. In other words, each credible set of gene-tissue pairs was counted only once. For TGFM, we first removed the gene-tissue pairs with PIPs < 0.5, and then applied the same procedure as TGVIS to count the silver and nearby genes. Similar to the mapping procedure for silver genes, we examined each credible set identified as causal to see if it contained any druggable genes. If a druggable gene is present, we count it once.

We used the following statistics to compare the enrichments of TGVIS and TGFM. For example, regarding TGVIS and a given trait, we consider three metrics: the number of causal genes identified by TGVIS, the overlap between genes identified by TGVIS and those in the drug-target list, and the ratio of these two metrics (referred to as Ratio hereafter). To compare whether TGVIS or TGFM had a higher enrichment across traits, we performed a paired t-test using two vectors of Ratio.

### Colocalization of credible sets

We use colocalization to evaluate the causal credible sets identified by TGFM and TGVIS. Within each region, we select variants from the outcome GWAS with P-values less than 5E-5 and *r*^2^ < 0.81 for colocalization analysis. We perform fine-mapping on both the outcome and the gene-tissue pairs within credible sets using SuSiE, then calculate the posterior probability for hypothesis *H*_4_, i.e., both traits are associated and share the same single causal variant, between each outcome and exposure pair using Coloc-SuSiE. We use a posterior probability of *H*_4_ greater than 0.5 as the threshold to determine colocalization evidence between gene-tissue pairs and the outcome; notably, as long as at least one variant meets this criterion, it suffices.

### Mendelian Randomization using pQTL summary data

We performed both univariable and multivariable MR using pQTLs of protein abundance as IVs to evaluating the identified causal tissue-gene pairs. Because to the lack of tissue-specific protein data, we focused on a subset of pGenes identified in blood tissues provided by Sun et al.^10^. We selected independent, genome-wide significant pQTLs for each protein as IVs. The selection method for independent IVs was C+T (--clump-kb 1000 --clump-p1 5e-8 --clump-p2 5e-8 --clump-r2 0.01 using PLINK), with LD reference panels consisting of the 9,680 individuals and 9.3M variants from UKBB. We applied five univariable MR methods: MRMedian^56^, IMRP^57^, MRCML^58^, MRCUE^59^, and MRBEE^60^. Both MRCUE and MRBEE account for sample overlap, with sample overlap correlations estimated using insignificant variants. We used the R package FDREstimation to convert the P-values obtained by these methods to FDR Q-values, using “BY” as the adjustment method. A pGene was considered significant if it was identified as such by at least four methods. We also conducted an analysis comparing the enrichments of TGVIS and TGFM, where the three corresponding metrics are: the overlap between causal genes identified by TGVIS or TGFM and the pGenes reported in Sun et al.^10^, the number of significant pGenes identified in univariable MR analysis, and the ratio of these two metrics.

## Author contributions

Y.Y and X.Z conceived and designed the study. Y.Y. performed all analysis. Y.Y and X.Z. drafted the manuscript. N.L. edited the manuscript. X.Z. supervised this project.

## Acknowledgments

This work was supported by grant HG011052 and HG011052-03S1 (to X.Z.) from the National Human Genome Research Institute (NHGRI), USA.

## Ethics approval

The study was approved by the institutional review board (IRB number: STUDY20180592) at Case Western Reserve University

## Data Availability

The GWAS summary data, eQTL summary data, and pQTL summary data used in this study can be downloaded from the “Data available” section of the literature listed in **Table S1-S2**. The GTEx summary data can be obtained from https://gtexportal.org/home/downloads/adult-gtex/qtl. The GWAS data in the million veteran program (MVP) are available through database of genotypes and phenotypes (dbGAP) under accession number phs001672.v7.p1. The individuallevel data from the UKBB used for estimating the LD matrix was accessed through Application ID: 81097.

## Code Availability

The code for the analyses presented in this paper is available in the **Supplementary Materials**, complete with step-by-step instructions. The TGVIS R package can be downloaded from https://github.com/harryyiheyang/TGVIS.

## References

1. Graham, S. E. et al. The power of genetic diversity in genome-wide association studies of lipids. Nature 600, 675–679 (2021).

2. Yengo, L. et al. A saturated map of common genetic variants associated with human height. Nature 610, 704–712 (2022).

3. Suzuki, K. et al. Genetic drivers of heterogeneity in type 2 diabetes pathophysiology. Nature 627, 347–357 (2024).

4. Mostafavi, H., Spence, J. P., Naqvi, S. & Pritchard, J. K. Systematic differences in discovery of genetic effects on gene expression and complex traits. Nat. Genet. 55, 1866–1875 (2023).

5. Wallace, C. A more accurate method for colocalisation analysis allowing for multiple causal variants. PLOS Genet. 17, e1009440 (2021).

6. Gamazon, E. R. et al. A gene-based association method for mapping traits using reference transcriptome data. Nat. Genet. 47, 1091–1098 (2015).

7. Gkatzionis, A., Burgess, S. & Newcombe, P. J. Statistical methods for cis-Mendelian randomization with two-sample summary-level data. Genet. Epidemiol. 47, 3–25 (2023).

8. Wainberg, M. et al. Opportunities and challenges for transcriptome-wide association studies. Nat. Genet. 51, 592–599 (2019).

9. Li, Y. I. et al. RNA splicing is a primary link between genetic variation and disease. Science 352, 600–604 (2016).

10. Sun, B. B. et al. Plasma proteomic associations with genetics and health in the UK Biobank. Nature 622, 329–338 (2023).

11. Amariuta, T., Siewert-Rocks, K. & Price, A. L. Modeling tissue co-regulation estimates tissue-specific contributions to disease. Nat. Genet. 55, 1503–1511 (2023).

12. The GTEx Consortium et al. The GTEx Consortium atlas of genetic regulatory effects across human tissues. Science 369, 1318–1330 (2020).

13. Zhao, S. et al. Adjusting for genetic confounders in transcriptome-wide association studies improves discovery of risk genes of complex traits. Nat. Genet. 56, 336–347 (2024).

14. Strober, B. J., Zhang, M. J., Amariuta, T., Rossen, J. & Price, A. L. Fine-mapping causal tissues and genes at disease-associated loci. Preprint at 10.1101/2023.11.01.23297909 (2023).

15. Burgess, S. & Thompson, S. G. Mendelian Randomization. (Chapman and Hall/CRC, 2021). doi:10.1201/9780429324352.

16. Wang, G., Sarkar, A., Carbonetto, P. & Stephens, M. A simple new approach to variable selection in regression, with application to genetic fine mapping. J. R. Stat. Soc. Ser. B Stat. Methodol. 82, 1273–1300 (2020).

17. Cui, R. et al. Improving fine-mapping by modeling infinitesimal effects. Nat. Genet. 56, 162–169 (2024).

18. Zou, Y., Carbonetto, P., Wang, G. & Stephens, M. Fine-mapping from summary data with the “Sum of Single Effects” model. PLOS Genet. 18, e1010299 (2022).

19. Benner, C. et al. FINEMAP: efficient variable selection using summary data from genome-wide association studies. Bioinformatics 32, 1493–1501 (2016).

20. Liang, Y., Nyasimi, F. & Im, H. K. On the problem of inflation in transcriptome-wide association studies. Preprint at 10.1101/2023.10.17.562831 (2023).

21. Sidorenko, J. et al. Genetic architecture reconciles linkage and association studies of complex traits. Nat. Genet. (2024) doi:10.1038/s41588-024-01940-2.

22. Wood, S. N. Fast stable restricted maximum likelihood and marginal likelihood estimation of semiparametric generalized linear models. J. R. Stat. Soc. Ser. B Stat. Methodol. 73, 3–36 (2011).

23. Aschard, H. A perspective on interaction effects in genetic association studies. Genet. Epidemiol. 40, 678–688 (2016).

24. Nielsen, J. B. et al. Biobank-driven genomic discovery yields new insight into atrial fibrillation biology. Nat. Genet. 50, 1234–1239 (2018).

25. Surendran, P. et al. Discovery of rare variants associated with blood pressure regulation through meta-analysis of 1.3 million individuals. Nat. Genet. 52, 1314–1332 (2020).

26. Sinnott-Armstrong, N. et al. Genetics of 35 blood and urine biomarkers in the UK Biobank. Nat. Genet. 53, 185–194 (2021).

27. Pazoki, R. et al. Genetic analysis in European ancestry individuals identifies 517 loci associated with liver enzymes. Nat. Commun. 12, 2579 (2021).

28. Young, W. J. et al. Genetic analyses of the electrocardiographic QT interval and its components identify additional loci and pathways. Nat. Commun. 13, 5144 (2022).

29. Ghouse, J. et al. Genome-wide meta-analysis identifies 93 risk loci and enables risk prediction equivalent to monogenic forms of venous thromboembolism. Nat. Genet. 55, 399–409 (2023).

30. Aragam, K. G. et al. Discovery and systematic characterization of risk variants and genes for coronary artery disease in over a million participants. Nat. Genet. 54, 1803–1815 (2022).

31. Roychowdhury, T. et al. Genome-wide association meta-analysis identifies risk loci for abdominal aortic aneurysm and highlights PCSK9 as a therapeutic target. Nat. Genet. 55, 1831–1842 (2023).

32. Zhu, X., Zhu, L., Wang, H., Cooper, R. S. & Chakravarti, A. Genome-wide pleiotropy analysis identifies novel blood pressure variants and improves its polygenic risk scores. Genet. Epidemiol. 46, 105–121 (2022).

33. Han, S. K. et al. Mapping genomic regulation of kidney disease and traits through high-resolution and interpretable eQTLs. Nat. Commun. 14, 2229 (2023).

34. Viñuela, A. et al. Genetic variant effects on gene expression in human pancreatic islets and their implications for T2D. Nat. Commun. 11, 4912 (2020).

35. Zhou, H. et al. FAVOR: functional annotation of variants online resource and annotator for variation across the human genome. Nucleic Acids Res. 51, D1300–D1311 (2023).

36. Trajanoska, K. et al. From target discovery to clinical drug development with human genetics. Nature 620, 737–745 (2023).

37. GTEx Consortium et al. Estimating the causal tissues for complex traits and diseases. Nat. Genet. 49, 1676–1683 (2017).

38. Arvanitis, M., Tayeb, K., Strober, B. J. & Battle, A. Redefining tissue specificity of genetic regulation of gene expression in the presence of allelic heterogeneity. Am. J. Hum. Genet. 109, 223–239 (2022).

39. Burgess, S., Dudbridge, F. & Thompson, S. G. Combining information on multiple instrumental variables in Mendelian randomization: comparison of allele score and summarized data methods. Stat. Med. 35, 1880–1906 (2016).

40. Grant, A. J. & Burgess, S. An efficient and robust approach to Mendelian randomization with measured pleiotropic effects in a high-dimensional setting. Biostatistics 23, 609– 625 (2022).

41. Barbeira, A. N. et al. Exploring the phenotypic consequences of tissue specific gene expression variation inferred from GWAS summary statistics. Nat. Commun. 9, 1825 (2018).

42. Kirk, T., Ahmed, A. & Rognoni, E. Fibroblast memory in development, homeostasis and disease. Cells 10, 2840 (2021).

43. Wittich, H. et al. Transcriptome-wide association study of the plasma proteome reveals cis and trans regulatory mechanisms underlying complex traits. Am. J. Hum. Genet. 111, 445–455 (2024).

44. Ross, S. D. et al. Clinical outcomes in statin treatment trials: A meta-analysis. Arch. Intern. Med. 159, 1793 (1999).

45. Xu, Y. et al. Rs7206790 and rs11644943 in FTO gene are associated with risk of obesity in Chinese school-age population. PLoS ONE 9, e108050 (2014).

46. Campos, A. I. et al. Boosting the power of genome-wide association studies within and across ancestries by using polygenic scores. Nat. Genet. 55, 1769–1776 (2023).

47. Yang, Y., Lee, D., Chakravarti, A. & Zhu, X. Partitioning tissue-specific heritability and identifying tissue-specific causal genes of blood pressure traits using CRE annotations. Manuscript.

48. Okamoto, J. et al. Probabilistic Fine-mapping of putative causal genes. Preprint at 10.1101/2024.10.27.620482 (2024).

49. Yazar, S. et al. Single-cell eQTL mapping identifies cell type–specific genetic control of autoimmune disease. Science 376, eabf3041 (2022).

50. Claussnitzer, M. et al. FTO obesity variant circuitry and adipocyte browning in humans. N. Engl. J. Med. 373, 895–907 (2015).

51. Zhu, X. & Stephens, M. Bayesian large-scale multiple regression with summary statistics from genome-wide association studies. Ann. Appl. Stat. 11, (2017).

52. Schwarz, G. Estimating the dimension of a model. Ann. Stat. 6, (1978).

53. Zhang, D. & Lin, X. Hypothesis testing in semiparametric additive mixed models. Biostatistics 4, 57–74 (2003).

54. Willer, C. J., Li, Y. & Abecasis, G. R. METAL: fast and efficient meta-analysis of genomewide association scans. Bioinformatics 26, 2190–2191 (2010).

55. Purcell, S. et al. PLINK: A tool set for whole-genome association and population-based linkage analyses. Am. J. Hum. Genet. 81, 559–575 (2007).

56. Yavorska, O. O. & Burgess, S. MendelianRandomization: an R package for performing Mendelian randomization analyses using summarized data. Int. J. Epidemiol. 46, 1734– 1739 (2017).

57. Zhu, X., Li, X., Xu, R. & Wang, T. An iterative approach to detect pleiotropy and perform Mendelian Randomization analysis using GWAS summary statistics. Bioinformatics 37, 1390–1400 (2021).

58. Lin, Z., Xue, H. & Pan, W. Robust multivariable Mendelian randomization based on constrained maximum likelihood. Am. J. Hum. Genet. 110, 592–605 (2023).

59. Cheng, Q., Zhang, X., Chen, L. S. & Liu, J. Mendelian randomization accounting for complex correlated horizontal pleiotropy while elucidating shared genetic etiology. Nat. Commun. 13, 6490 (2022).

60. Lorincz-Comi, N., Yang, Y., Li, G. & Zhu, X. MRBEE: A bias-corrected multivariable Mendelian Randomization method. Hum. Genet. Genomics Adv. 100290 (2024) doi:10.1016/j.xhgg.2024.100290.

