## Supplementary Materials for "Uncovering causal gene-tissue pairs and variants: A multivariable TWAS method controlling for infinitesimal effects"

### 1 Real data analysis

#### 1.1 Data structure

Here we illustrate a real example of how to perform TGVIS and also TGFM in real data analysis of the manuscript. This tutorial starts with the structures of involved data.

##### 1.1.1 LD reference panel

The first dataset is the LD reference panel. This dataset is derived from the 9,680 unrelated individuals we described in the paper, selected from approximately 500,000 imputed individuals in the UK Biobank (UKBB). We refined the data using the bim files from these individuals. The files we shared on Google Drive include all 9.32 million SNPs involved; however, in this tutorial, we will focus only on a subset in the PCSK9 locus. Below is a glimpse of the data structure:

```
library(data.table)
library(dplyr)
variant=readRDS("variant.rds")
variant
```

```
##                       SNP CHR       BP  A1    A2      Freq MarkerName
##     1: 1:54005191_CCACA_C   1 54005191   C CCACA 0.3992490 1:54005191
##     2:    1:54012271_AG_A   1 54012271  AG     A 0.6292130 1:54012271
##     3:    1:54012621_TG_T   1 54012621  TG     T 0.6295230 1:54012621
##     4:    1:54035956_TA_T   1 54035956   T    TA 0.1912090 1:54035956
##     5:   1:54070614_TAC_T   1 54070614 TAC     T 0.9896660 1:54070614
##    ---
## 10782:          rs9919296   1 54578136   C     T 0.0209929 1:54578136
## 10783:          rs9919314   1 54579134   C     T 0.0209763 1:54579134
## 10784:           rs993075   1 56994938   G     C 0.4923560 1:56994938
## 10785:          rs9970807   1 56965664   T     C 0.0921541 1:56965664
## 10786:           rs998154   1 55596384   C     T 0.1690460 1:55596384
```

In this reference panel, `SNP` is the identifier for the variants; `CHR` represents the chromosome; `BP` indicates the base pair position in the hg19 genome build; `A1` is the effect allele as specified in the BED file; `A2` is the other allele; `Freq` denotes the frequency of the effect allele; and `MarkerName` is another unique identifier for the variants in the format CHR:BP. It is important to note that some variants in the UKBB bed file do not have an rsID. For these variants, their `SNP` are in the format CHR:BP:A2:A1.

##### 1.1.2 GWAS summary data

The second dataset is the GWAS summary data, which should include at least the following columns: `SNP`, `A1`, `A2`, `Zscore`, and `N`. In this dataset, `Zscore` represents the Z-score of the marginal effect size estimates from the outcome GWAS, while `N` denotes the sample size. Other statistics can be deduced from `Zscore` and `N`, e.g.,

$$\mathtt{BETA} = \frac{\mathtt{Zscore}}{\sqrt{\mathtt{N}}}, \quad \mathtt{SE} = \frac{1}{\sqrt{\mathtt{N}}}.$$

Below is an example of the dataset's structure:

```r
library(arrow)
LDL=read_parquet("LDL.parquet")
LDL[-5,]
```

```
##                    SNP CHR        BP  A1  A2      Zscore       N
##    1: 1:54388067_CAA_C   1 54388067   C CAA  -0.9536883  389563
##    2: 1:54389774_TAG_T   1 54389774 TAG   T  -0.7761577  487566
##    3:  1:54401303_CT_C   1 54401303   C  CT   1.0402465 1041795
##    4: 1:54406627_ATT_A   1 54406627   A ATT  -3.1568909 1065377
##    5:  1:54430681_GT_G   1 54430681   G  GT  -2.6594631 1058084
##   ---
## 7958:        rs9919142   1 54579153   G   A   1.0001670 1230988
## 7959:        rs9919295   1 54578100   C   T   0.9664437 1231010
## 7960:        rs9919296   1 54578136   C   T   0.9008325 1231009
## 7961:        rs9919314   1 54579134   C   T   0.9813689 1231009
## 7962:         rs998154   1 55596384   C   T -10.8367174 1231250
```

It should be noted that we did not use the original `SNP` identifiers from the GWAS. Instead, we merged the GWAS data with the variant file using the `MarkerName` (CHR:BP) identifiers, and then assigned `SNP` from the variant file to the corresponding entries in the GWAS data. In cases where the GWAS file is based on the hg38 genome build, we use LiftOver to convert it to hg19.

##### 1.1.3 eQTL and sQTL summary data

The next dataset, which has a more complex structure, is the summary data for eQTL and sQTL. We preprocessed the data provided by GTEx and other studies to retain only the following columns: `SNP`, `CHR`, `BP`, `A1`, `A2`, `P`, `Zscore`, `N`, `Gene`, `GeneSymbol`, `Tissue`, `Variable`, and `xQTL`. Here,

- `P`: the P-value of the marginal effect for the xQTL;
- `Gene`: the Ensembl ID of the gene associated with the xQTL;
- `GeneSymbol`: the symbol for the corresponding gene,
- `Tissue`: the tissue in which the gene was sequenced;
- `Variable`: the combination of GeneSymbol (or Gene for sQTL) + Tissue
- `xQTL`: an indicator of sQTL or eQTL.

An example of the dataset structure is shown below:

```r
eQTLsQTL=read_parquet("eQTLsQTL.parquet")%>%
dplyr::select(SNP,CHR,BP,A1,A2,P,Zscore,N,Gene,GeneSymbol,Tissue,Variable,xQTL)
eQTLsQTL
```

```
##                  SNP CHR       BP A1 A2          P     Zscore   N
##        1: rs12090789   1 54387577  T  C 0.78223604 -0.2765702 124
##        2: rs12090789   1 54387577  T  C 0.58952588  0.5399070 124
##        3: rs12090789   1 54387577  T  C 0.54357336  0.6078765 124
##        4: rs12090789   1 54387577  T  C 0.35021166 -0.9351431 124
##        5: rs12090789   1 54387577  T  C 0.59624067 -0.5301882 124
##       ---
```

```
## 15168101:  rs6680237   1 56586870  G  C 0.34938904 -0.9375335 311
## 15168102:  rs6680237   1 56586870  G  C 0.59527600 -0.5316440 420
## 15168103:  rs6680237   1 56586870  G  C 0.86508400 -0.1700180 420
## 15168104:  rs6680237   1 56586870  G  C 0.60456600 -0.5182680 420
## 15168105:  rs6680237   1 56586870  G  C 0.00636787 -2.7430910 420
##                                                       Gene   GeneSymbol
##        1: chr1:52906611:52907868:clu_52882:ENSG00000121310.16 ECHDC2(sQTL)
##        2: chr1:52917615:52921553:clu_52883:ENSG00000121310.16 ECHDC2(sQTL)
##        3: chr1:52914095:52921553:clu_52883:ENSG00000121310.16 ECHDC2(sQTL)
##        4: chr1:52914095:52917544:clu_52883:ENSG00000121310.16 ECHDC2(sQTL)
##        5: chr1:52914095:52916465:clu_52883:ENSG00000121310.16 ECHDC2(sQTL)
##      ---
## 15168101:                                     ENSG00000173406         DAB1
## 15168102:                                     ENSG00000162407        PLPP3
## 15168103:                                     ENSG00000162409       PRKAA2
## 15168104:                                     ENSG00000187889         FYB2
## 15168105:                                     ENSG00000021852          C8B
##                       Tissue
##        1: Adipose_Subcutaneous
##        2: Adipose_Subcutaneous
##        3: Adipose_Subcutaneous
##        4: Adipose_Subcutaneous
##        5: Adipose_Subcutaneous
##      ---
## 15168101:          Kidney_Tube
## 15168102:               Islets
## 15168103:               Islets
## 15168104:               Islets
## 15168105:               Islets
##                                                                       Variable
##        1: chr1:52906611:52907868:clu_52882:ENSG00000121310.16+Adipose_Subcutaneous
##        2: chr1:52917615:52921553:clu_52883:ENSG00000121310.16+Adipose_Subcutaneous
##        3: chr1:52914095:52921553:clu_52883:ENSG00000121310.16+Adipose_Subcutaneous
##        4: chr1:52914095:52917544:clu_52883:ENSG00000121310.16+Adipose_Subcutaneous
##        5: chr1:52914095:52916465:clu_52883:ENSG00000121310.16+Adipose_Subcutaneous
##      ---
## 15168101:                                                         DAB1+Kidney_Tube
## 15168102:                                                            PLPP3+Islets
## 15168103:                                                           PRKAA2+Islets
## 15168104:                                                             FYB2+Islets
## 15168105:                                                              C8B+Islets
##           xQTL
##        1: sQTL
##        2: sQTL
##        3: sQTL
##        4: sQTL
##        5: sQTL
##      ---
## 15168101: eQTL
## 15168102: eQTL
## 15168103: eQTL
## 15168104: eQTL
## 15168105: eQTL
```

It should be noted that entries in the format chr1:52906611:52907868:clu_52882:ENSG00000121310.16 indicate a specific splicing event for the gene ENSG00000121310, as defined by LeafCutter.

In practice, combining multiple sQTL and eQTL datasets is a challenging task. In the section where we explain the key functions, we will detail our strategy for handling and integrating these datasets.

##### 1.1.4 LD refernece panel with individual

We used the UKBB BED file to estimate the LD reference, with a sample size of 9,680. Below is a glimpse of the data structure:

```
UKBBGenotype=readRDS("UKBBGenotype.rds")
UKBBGenotype[1:10,1:5]
```

```
##    rs10047036 rs12728734 rs150256195 rs74510493 rs114570917
## 1           2          2           2          2           2
## 2           2          1           2          2           2
## 3           2          2           2          2           2
## 4           1          1           2          2           2
## 5           2          2           2          2           2
## 6           2          2           2          2           2
## 7           2          2           2          2           2
## 8           2          2           2          2           1
## 9           1          2           2          2           2
## 10          2          2           2          2           2
```

#### 1.2 Step-by-step analysis

##### 1.2.1 Allele harmonisation

In the first step, we adjust the direction of the Z-scores in the GWAS and xQTL summary data to ensure that the effect alleles in these datasets match the effect alleles in our reference panel. This step is crucial because the LD matrix is estimated from this reference panel, and accurate LD estimation is fundamental to all statistical methods based on GWAS summary data. We wrote a function, `allele_harmonise()` in the R package `TGVIS`, to perform this step:

```
library(TGVIS)
LDL=allele_harmonise(ref_panel=variant[,c("SNP","A1","A2")],gwas_data=LDL)
eQTLsQTL=allele_harmonise(ref_panel=variant[,c("SNP","A1","A2")],gwas_data=eQTLsQTL)
eQTLsQTL=eQTLsQTL[LDL,nomatch=0]
setnames(eQTLsQTL,"i.Zscore","Zscore.y")
setnames(eQTLsQTL,"Zscore","Zscore.x")
```

In `allele_harmonise()`, we automatically set `gwas_data` as a data.table with `key="SNP"`, allowing `eQTLsQTL=eQTLsQTL[LDL, nomatch=0]` to efficiently merge the two datasets. The reason for merging these datasets, as described in here, there is a vast and heterogeneous landscape of publicly available GWAS. These studies were conducted on different genotyping platforms, using different imputation schemes, and defined on different releases of the human genome., is that there is a vast and heterogeneous landscape of publicly available GWAS. These studies were conducted on different genotyping platforms, using different imputation schemes, and defined on different releases of the human genome. Therefore, we aim to find the common variants between the GWAS and xQTL summary data to perform the analysis.

##### 1.2.2 Extracting the moderately correlated variants

The next step is to remove highly correlated variants using C+T. Although SuSiE can group highly correlated or statistically duplicated variants into a single group and assign them one effect, including many redundant variants can significantly increase the dimensionality of the model. Therefore, primarily to enhance computational efficiency, we recommend retaining only moderately correlated variants.

We use the smallest p-value across all tissue pairs corresponding to each variant as the input p-value for PLINK to extract a subset of moderately correlated variants. While we will not execute the following steps in this tutorial, we will provide the code for you. You can modify the file paths as needed for your own data.

```
A=eQTLsQTL%>%dplyr::select(SNP,P)
A=A[, .SD[which.min(P)], by=SNP]
A=A[which(A$P<5e-4),]
write.table(A,"Your_path.txt",quote=F, sep="\t", row.name=F)
setwd("Your_path_to_PLINK")
system("./plink --bfile Your_bed_file --clump Your_path.txt
            --clump-field P --clump-kb 1000 --clump-p1 1e-5
            --clump-p2 1e-5 --clump-r2 0.5 --out Your_path")
setwd("Your_analysis_path")
plinkfile=fread("Your_path.clumped")
plinkfile=plinkfile$SNP
```

The most important part in this step is:

- `-clump-kb 1000`: we consider the window size to be 1M,
- `-clump-p1 1e-5`: we use the threshold of 1E-5,
- `-clump-r2 0.5`: the correlation between two variants is in the range $(-\sqrt{0.5}, \sqrt{0.5})$.

Since direct causal variants might not be linked to any gene-tissue pairs, I perform clumping on the outcome GWAS to identify outcome-associated variants:

```
A1=LDL[which(LDL$SNP%in%unique(eQTLsQTL$SNP)),]
A1$P=pchisq(A1$Zscore^2,1,lower.tail=F);
A1=A1[,c("SNP","P")]
A1=A1[which(A1$P<min(5e-8,quantile(A1$P,0.1))),]
write.table(A1,"Your_path.txt",quote=F, sep="\t", row.name=F)
setwd("Your_path_to_PLINK")
system("./plink --bfile Your_bed_file --clump Your_path.txt
            --clump-field P --clump-kb 1000 --clump-p1 1e-5
            --clump-p2 1e-5 --clump-r2 0.5 --out Your_path")
setwd("Your_analysis_path")
plinkfile1=fread("Your_path.clumped")
plinkfile1=plinkfile1$SNP
```

Finally, we merge these two lists of variants and use C+T to remove any highly correlated variants (which are typically few), resulting in the final pool of variants for analysis:

```
A=data.frame(SNP=unique(c(plinkfile,plinkfile1)))
A$P=0.05
write.table(A,"Your_path.txt",quote=F, sep="\t", row.name=F)
```

```
setwd("Your_path_to_PLINK")
system("./plink --bfile Your_bed_file --clump Your_path.txt
            --clump-field P --clump-kb 1000 --clump-p1 1e-5
            --clump-p2 1e-5 --clump-r2 0.5 --out Your_path")
setwd("Your_analysis_path")
plinkfile=fread("Your_path.clumped")
rsid=plinkfile$SNP
```

We have recorded this pool of variants in the PCSK9 locus:

```
rsid=readRDS("SNP_lowLD.rds")
gwas_eQTLsQTL=eQTLsQTL[which(eQTLsQTL$SNP%in%rsid),]
```

#### 1.3 Regularization of LD matrix

Our next step is to estimate a "good" LD matrix. We use the POET-shrinkage method (Fan et al., 2013), as described in this paper, to estimate such an LD matrix. The code is as follows:

```
R0=cor(UKBBGenotype)
R0[is.na(R0)]=0;diag(R0)=1
R0=poet_shrinkage(R0)
R0=(t(R0)+R0)/2
genosnp=colnames(UKBBGenotype)
rownames(R0)=colnames(R0)=genosnp
```

#### 1.4 Construction of design matrix of gene-tissue pairs

Our next step is to extract the design matrix of gene-tissue pairs from the `eQTLsQTL` data.table. We provide the function `make_design_matrix()`, which converts the Z-scores in `eQTLsQTL` into an $M \times p$ design matrix, where $M$ is the number of variants and $p$ is the total number of gene-tissue pairs:

```
bX0=make_design_matrix(eQTLsQTL[,c("SNP","Variable","Zscore.x")])
bX0=bX0[genosnp,]
```

In data.table `eQTLsQTL`, `Zscore.x` represents the Z-score of the xQTL effect, while `Zscore.y` is the Z-score from the outcome GWAS. We match the rows of `bX0` with the LD matrix using the code `bX0=bX0[genosnp,]`.

Our next step is to impute missing values in the Z-scores as 0. Since GTEx only provides the marginal xQTL effect sizes for variants near the gene's TSS, this results in missing values. Before imputing, we remove variants and gene-tissue pairs with excessive missing values. In this analysis, we exclude variants with more than 95% missing values and genes with more than 90% missing values:

```
bX=remove_missing_row_column(bX0,rowfirst=F,rowthres=0.95,colthres=0.9)
genosnp=rownames(bX)
R0=R0[genosnp,genosnp]
bX=as.matrix(bX[genosnp,])
bX[is.na(bX)]=0
```

##### 1.4.1 eQTL selection

Our next step is to use SuSiE for gene-tissue pair eQTL selection. The first step is to extract the average sample size for each gene-tissue pair from eQTLsQTL to use as the input sample size for SuSiE:

```
VariableName=unique(eQTLsQTL$Variable)
NeQTLsQTL=eQTLsQTL[,.(NeQTLsQTL=mean(N)),by=Variable][Variable%in%VariableName,NeQTLsQTL]
names(NeQTLsQTL)=VariableName
NeQTLsQTL=NeQTLsQTL[colnames(bX)]
```

We have encapsulated a for-loop function based on `susie_rss()` to perform eQTL selection for each gene-tissue pair:

```
fiteQTL=eQTLmapping_susie(bX=bX,LD=R0,Nvec=NeQTLsQTL,L=3,pip.thres=0.5,pip.min=0.25)
```

```
##   |                                                                              |
```

```
bXest=fiteQTL$Estimate
ind=which(colSums(abs(bXest))==0)
bXest=bXest[,-ind]
bX=bX[,-ind]
NeQTLsQTL=NeQTLsQTL[colnames(bX)]
```

##### 1.4.2 Performing S-Predixcan and its modifier to remove noise gene-tissue pairs

In this example, our original design matrix includes $M = 381$ variants and $p = 4878$ gene-tissue pairs:

```
dim(bX0)
```

```
## [1]  381 4878
```

After quality control and eQTL selection, we retained $p = 664$ gene-tissue pairs.

```
dim(bX)
```

```
## [1] 379 664
```

However, in many cases, the original number of gene-tissue pairs could be tens of thousands, and even after eQTL selection, there could still be thousands of gene-tissue pairs. Therefore, we perform a univariable TWAS with S-PrediXcan and its modifier to pre-reduce the dimensionality, making TGVIS and TGFM more efficient.

Let's first organize the data and set up a data frame to store the results:

```
bY=eQTLsQTL[,c("SNP","Zscore.y")]
bY=bY[!duplicated(bY$SNP),]
rownames(bY)=bY$SNP
bY=bY[genosnp,]
UVTWAS=matrix(0,ncol(bXest),6)
colnames(UVTWAS)=c("Variable","Type","Est1","P1","Est2","P2")
UVTWAS=as.data.frame(UVTWAS)
UVTWAS[,1]=colnames(bXest)
UVTWAS[,2]=eQTLsQTL$xQTL[match(UVTWAS[,1],eQTLsQTL$Variable)]
UVTWAS[,c(4,6)]=1
```

Next, we execute S-PrediXcan and its modifier:

```
for(i in 1:ncol(bXest)){
errorindicate=0
tryCatch({
bxx=bX[,i]
indx=which(bxx!=0)
bx=bXest[indx,i]
by=bY$Zscore.y[indx]
bxx=bxx[indx]
if(sum(bx!=0)==1){
pleiotropy.rm=which(bx!=0)
}else{
pleiotropy.rm=NULL
}
fitModifier=modified_predixcan(by=by,bxest=bx,LD=R0[indx,indx],
            pleiotropy.rm=pleiotropy.rm,tauvec=seq(3,21,by=3))
fitSpredixcan=modified_predixcan(by=by,bxest=bx,LD=R0[indx,indx],
            pleiotropy.rm=pleiotropy.rm,tauvec=10000000)
UVTWAS[i,3]=fitSpredixcan$theta
UVTWAS[i,4]=pchisq(fitSpredixcan$theta^2/fitSpredixcan$covtheta,1,lower.tail=F)
UVTWAS[i,5]=fitModifier$theta
UVTWAS[i,6]=pchisq(fitModifier$theta^2/fitModifier$covtheta,1,lower.tail=F)
}, error=function(e){
errorindicate=1
})
if(errorindicate == 1) next
}
```

The structure of UVTWAS is:

```
head(UVTWAS)
```

```
##                   Variable Type        Est1        P1       Est2        P2
## 1 ACOT11+Adipose_Visceral eQTL -0.49957076 0.7895277 -1.0746827 0.2311542
## 2    ACOT11+Adrenal_Gland eQTL -2.39903575 0.7147184 -2.9340518 0.3592333
## 3     ACOT11+Artery_Aorta eQTL  0.17665197 0.9181448  0.2538559 0.7569147
## 4    ACOT11+Artery_Tibial eQTL -0.02026495 0.9829979  0.1955781 0.6705236
## 5       ACOT11+Cerebellum eQTL -0.93448220 0.8659363  0.5692827 0.8332270
## 6           ACOT11+Cortex eQTL -0.40462949 0.8721248 -0.2022807 0.8692957
```

As described in the manuscript, we only consider data with an P-value greater than 0.5. We systematically scan each locus of the GWAS trait.

```r
Genelist=UVTWAS%>%mutate(pvthres=0.05) %>%
dplyr::filter(P1 < pvthres | P2 < pvthres) %>%
pull(Variable)%>%unique()
```

This leaves $p = 52$ gene-tissue pairs:

```r
length(Genelist)
```

```
## [1] 52
```

We then organize the data:

```r
bX=bX[,Genelist]
bXest=bXest[,Genelist]
bY=eQTLsQTL[,c("SNP","Zscore.y")]%>%as.data.frame(.)
bY=bY[!duplicated(bY$SNP),]
rownames(bY)=bY$SNP
bY=bY[genosnp,]
```

##### 1.4.3 Performing TGFM and TGVIS

The main procedures for TGFM and TGVIS are relatively straightforward, as shown below:

```r
fittgfm=tgfm(by=bY$Zscore.y,bX=bX,LD=R0, Nvec=c(mean(LDL$N),NeQTLsQTL[Genelist]),
         causal.sampling.time=100,eqtl.sampling.time=25,L.causal=10)
```

Before executing TGVIS, we do not recommend removing potential candidates for direct causal variants. Specifically, if a gene is solely an eQTL, we suggest not including these eQTLs as candidates for direct causal variants. We have defined a function, `findUniqueNonZeroRows()`, to identify the indices of these variants. The code for this function will be provided in GitHub.

Next, we execute TGVIS:

```r
fittgvis=tgvis(by=bY$Zscore.y,bXest=bXest,LD=R0,Noutcome=mean(LDL$N),
                 L.causal.vec=c(1:10),varinf.upper.boundary=0.25,pip.min=0.1,
                 pleiotropy.rm=findUniqueNonZeroRows(bXest))
```

It should be pointed out that TGVIS requires the esimates of the joint xQTL effect `bXest` while TGFM requires the marginal xQTL effect estimate `bX`.

Finally, we organize the results, which is a bit more complex. The principle is to retain only those gene-tissue pairs and direct variants that are included in the 95% credible set. As for TGFM:

```r
thetagamma=c(fittgfm$theta[which(fittgfm$theta!=0)],
         fittgfm$gamma[which(fittgfm$gamma!=0)])
if(length(thetagamma)>0){
se.mrjones=c(fittgfm$theta.se[which(fittgfm$theta!=0)],
         fittgfm$gamma.se[which(fittgfm$gamma!=0)])
pip.mrjones=c(fittgfm$theta.pip[which(fittgfm$theta!=0)],
          fittgfm$gamma.pip[which(fittgfm$gamma!=0)])
pratt.mrjones=c(fittgfm$theta.pratt[which(fittgfm$theta!=0)],
```

```
            fittgfm$gamma.pratt[which(fittgfm$gamma!=0)])
cs.mrjones=c(fittgfm$theta.cs[which(fittgfm$theta!=0)],
         fittgfm$gamma.cs[which(fittgfm$gamma!=0)])
cs.pip.mrjones=c(fittgfm$theta.cs.pip[which(fittgfm$theta!=0)],
             fittgfm$gamma.cs.pip[which(fittgfm$gamma!=0)])
TGFMResult=data.frame(Variable=names(thetagamma),estimate=thetagamma,
                  se=se.mrjones,pip=pip.mrjones,pratt=pratt.mrjones,
                  cs=cs.mrjones,cs.pip=cs.pip.mrjones)
TGFMResult$Type=c(rep("TissueGene",length(which(fittgfm$theta!=0))),
              rep("SNP",length(which(fittgfm$gamma!=0))))
TGFMResult$CHR=1
TGFMResult$BP=55.5e5
TGFMResult=TGFMResult%>%group_by(cs)%>%mutate(cs.pratt=sum(pratt))%>%ungroup()
TGFMResult=TGFMResult%>%group_by(cs)%>%mutate(cs.pip=sum(pip))%>%ungroup()
cs0=which(TGFMResult$cs==0)
if(length(cs0)>0){
TGFMResult$cs.pratt[cs0]=TGFMResult$pratt[cs0]
TGFMResult$cs.pip[cs0]=TGFMResult$pip[cs0]
}
}else{
TGFMResult=NULL
}
TGFMResult$xQTL=TGFMResult$Variable
if(sum(TGFMResult$Type=="TissueGene")>0){
TGFMResult$xQTL[which(TGFMResult$Type=="TissueGene")]=get_nonzero_rows(fittgfm$bXest,
               TGFMResult$Variable[which(TGFMResult$Type=="TissueGene")])$NonzeroRows
}
row.names(TGFMResult)=NULL
TGFMResult=TGFMResult%>%dplyr::select(Variable,cs,cs.pip,cs.pratt,xQTL,
                 CHR,BP,Type,estimate,se,pip,pratt)%>%arrange(.,cs,Type,Variable)
print(TGFMResult)
```

```
## # A tibble: 11 x 12
##    Variable    cs cs.pip cs.pratt xQTL    CHR     BP Type  estimate     se   pip
##    <chr>    <dbl>  <dbl>    <dbl> <chr> <dbl>  <dbl> <chr>    <dbl>  <dbl> <dbl>
##  1 rs11591~     1  1      0.524   rs11~     1 5.55e6 SNP     -76.6  1.02   1
##  2 PCSK9+W~     2  1      0.204   rs12~     1 5.55e6 Tiss~     6.46 0.258  1
##  3 rs11206~     3  1      0.0387  rs11~     1 5.55e6 SNP      18.9  0.352  1
##  4 rs11546~     4  1      0.0129  rs11~     1 5.55e6 SNP      -1.89 0.0411 0.121
##  5 chr1:54~     4  1      0.0129  rs11~     1 5.55e6 Tiss~   -10.5  0.228  0.879
##  6 rs24954~     5  1.00   0.0376  rs24~     1 5.55e6 SNP      15.3  0.666  1.00
##  7 rs15011~     6  1.00   0.0220  rs15~     1 5.55e6 SNP      13.4  0.748  1.00
##  8 Affx-52~     7  0.987  0.00873 Affx~     1 5.55e6 SNP     -10.8  0.781  0.987
##  9 rs39767~     8  1     -0.0259  rs39~     1 5.55e6 SNP      17.6  1.61   1
## 10 rs24793~     9  0.714  0.0179  rs24~     1 5.55e6 SNP       9.30 4.86   0.714
## 11 rs26472~    10  0.940 -0.00298 rs26~     1 5.55e6 SNP       9.75 2.37   0.940
## # i 1 more variable: pratt <dbl>
```

As for TGVIS:

```
thetagamma=c(fittgvis$theta[which(fittgvis$theta!=0)],
         fittgvis$gamma[which(fittgvis$gamma!=0)])
if(length(thetagamma)>0){
```

```r
se.mrjones=c(fittgvis$theta.se[which(fittgvis$theta!=0)],
         fittgvis$gamma.se[which(fittgvis$gamma!=0)])
pip.mrjones=c(fittgvis$theta.pip[which(fittgvis$theta!=0)],
          fittgvis$gamma.pip[which(fittgvis$gamma!=0)])
pratt.mrjones=c(fittgvis$theta.pratt[which(fittgvis$theta!=0)],
            fittgvis$gamma.pratt[which(fittgvis$gamma!=0)])
cs.mrjones=c(fittgvis$theta.cs[which(fittgvis$theta!=0)],
         fittgvis$gamma.cs[which(fittgvis$gamma!=0)])
cs.pip.mrjones=c(fittgvis$theta.cs.pip[which(fittgvis$theta!=0)],
             fittgvis$gamma.cs.pip[which(fittgvis$gamma!=0)])
TGVIResult=data.frame(Variable=names(thetagamma),estimate=thetagamma,
                  se=se.mrjones,pip=pip.mrjones,pratt=pratt.mrjones,
                  cs=cs.mrjones,cs.pip=cs.pip.mrjones)
TGVIResult$Type=c(rep("TissueGene",length(which(fittgvis$theta!=0))),
              rep("SNP",length(which(fittgvis$gamma!=0))))
TGVIResult$CHR=1
TGVIResult$BP=55.5e6
TGVIResult=TGVIResult%>%group_by(Type, cs)%>%mutate(cs.pratt=sum(pratt))%>%ungroup()
}else{
TGVIResult=NULL
}
TGVIResult$xQTL=TGVIResult$Variable
if(sum(TGVIResult$Type=="TissueGene")>0){
TGVIResult$xQTL[which(TGVIResult$Type=="TissueGene")]=
get_nonzero_rows(bXest,TGVIResult$Variable[which(TGVIResult$Type=="TissueGene")])$NonzeroRows
}
row.names(TGVIResult)=NULL
TGVIResult=TGVIResult%>%dplyr::select(Variable,cs,cs.pip,cs.pratt,xQTL,
          CHR,BP,Type,estimate,se,pip,pratt)%>%arrange(.,cs,Type,Variable)
print(TGVIResult)
```

```
## # A tibble: 3 x 12
##   Variable      cs cs.pip cs.pratt xQTL    CHR     BP Type  estimate    se   pip
##   <chr>      <dbl>  <dbl>    <dbl> <chr> <dbl>  <dbl> <chr>    <dbl> <dbl> <dbl>
## 1 rs11591147     1      1   0.492  rs11~     1 5.55e7 SNP      -71.8 1.02      1
## 2 PCSK9+Who~     2      1   0.170  rs12~     1 5.55e7 Tiss~     5.61 0.149     1
## 3 rs11206517     3      1   0.0398 rs11~     1 5.55e7 SNP       19.4 0.974     1
## # i 1 more variable: pratt <dbl>
```

##### 1.4.4 The Pratt indices of gene-tissue pairs, direct causal variants, and infinitesimal effect

We have defined a function, `R2_partition`, to calculate the Pratt indices for gene-tissue pairs, direct causal variants, and infinitesimal effects. For TGFM, the calculation is:

```r
TGFMPratt=R2_partition(y=bY$Zscore.y,LD=R0,
                   eta.theta=c(R0%*%fittgfm$bXest%*%fittgfm$theta),
                   eta.gamma=c(R0%*%fittgfm$gamma))
TGFMPratt
```

```
##         r1       r2        r3
## 1 0.834437 0.211783 0.6226539
```

Here, $r1$ is the total Pratt index, $r2$ is the Pratt index of gene-tissue pairs $r3$ is the Pratt index of direct causal variants.

```
TGVIPratt=R2_partition(y=bY$Zscore.y,LD=R0,
                       eta.theta=c(R0%*%bXest%*%fittgvis$theta),
                       eta.gamma=c(R0%*%fittgvis$gamma),
                       eta.upsilon=c(R0%*%fittgvis$upsilon))
TGVIPratt
```

```
##          r1        r2       r3        r4
## 1 0.9466211 0.1674825 0.529892 0.2492466
```

Here, $r4$ is the Pratt index of infinitesimal effects.

#### 1.5 Colocalization

Next, we will perform colocalization to evaluate the results of the causal credible sets identified by the two methods. First, we will apply SuSiE on the outcome:

```
library(susieR)
library(coloc)
```

```
## Warning: package 'coloc' was built under R version 4.3.3
```

```
## This is coloc version 5.2.3
```

```
fitoutcome=susie_rss(z=bY$Zscore.y,R=R0,n=mean(LDL$N),L=30)
fitoutcome=susie_rss(z=bY$Zscore.y,R=R0,n=mean(LDL$N),L=sum(fitoutcome$pip>0.5))
plot(fitoutcome$pip)
```

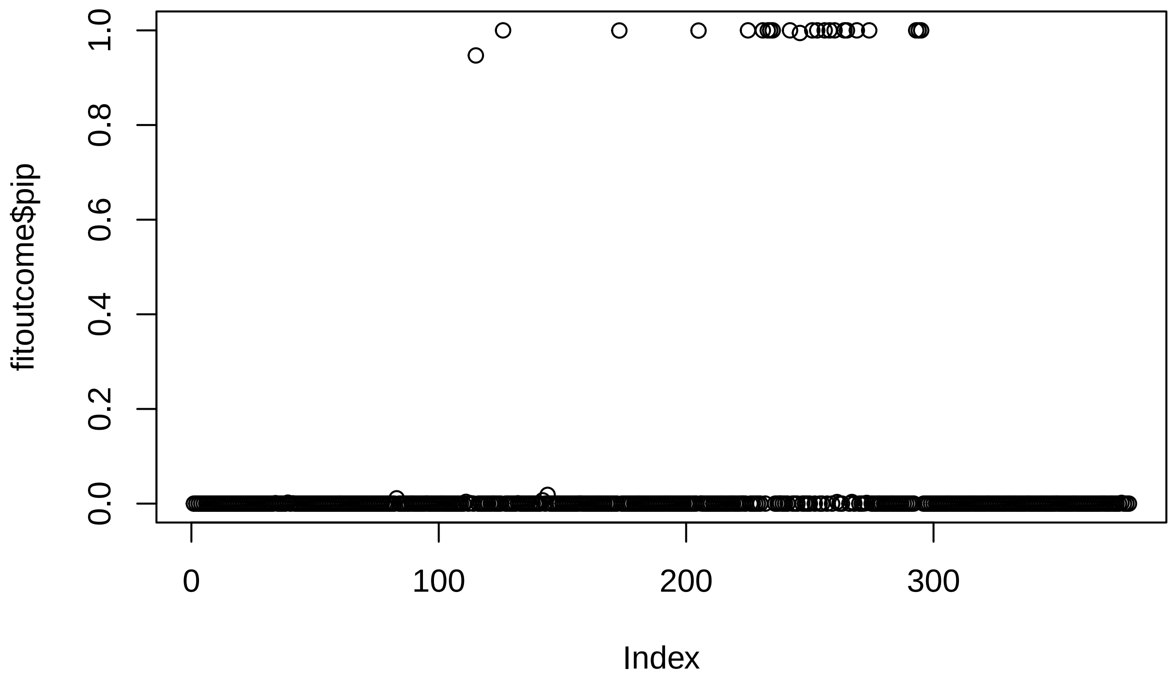

```
sum(fitoutcome$pip>0.5)
```

```
## [1] 23
```

We found a total of 23 variants with PIP > 0.5 in this region. Next, we will apply SuSiE to the PCSK9-WholeBlood data:

```
library(susieR)
library(coloc)
fitexposure=susie_rss(z=bX[,"PCSK9+Whole_Blood"],R=R0,n=NeQTLsQTL["PCSK9+Whole_Blood"],L=8)
fitexposure=susie_rss(z=bX[,"PCSK9+Whole_Blood"],R=R0,n=NeQTLsQTL["PCSK9+Whole_Blood"],
                      L=sum(fitoutcome$pip>0.5))
plot(fitexposure$pip)
```

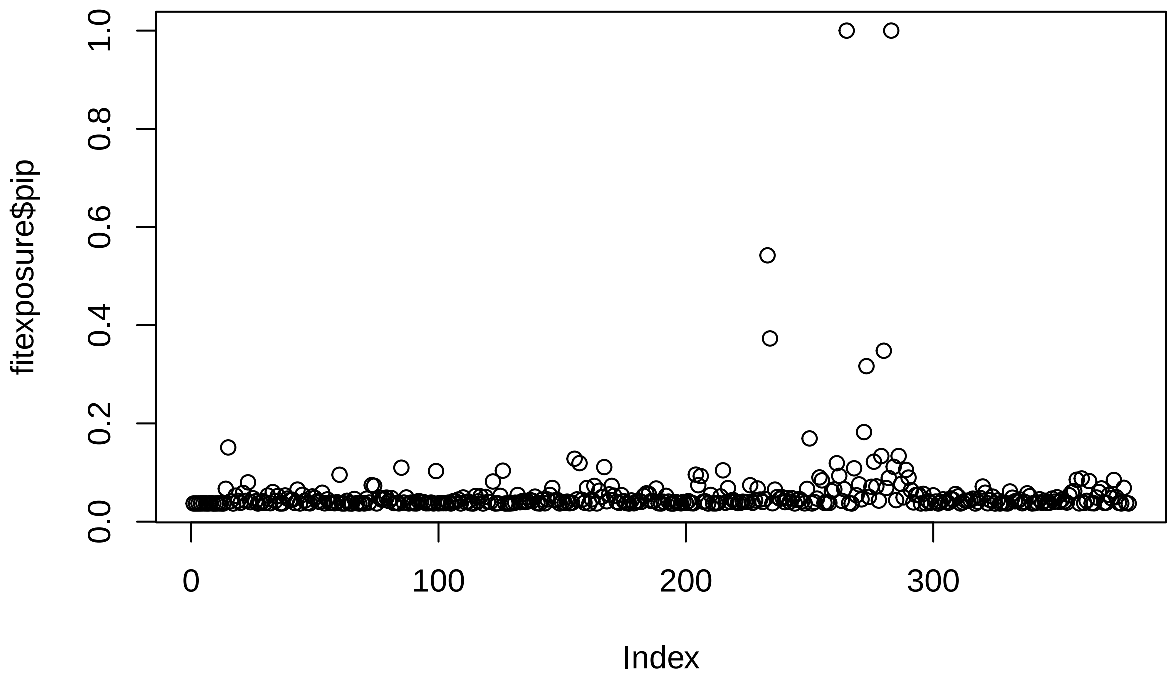

```r
sum(fitexposure$pip>0.5)
```

```
## [1] 3
```

Finally, we will use Coloc-SuSiE to evaluate the results:

```r
susie.res=coloc.susie(fitoutcome,fitexposure)
sum(susie.res$summary$PP.H4.abf>0.5)
```

```
## [1] 1
```

We identified a Coloc variant shared between LDL and PCSK9-WholeBlood.

It should be noted that in our main analysis, we selected all variants with P < 5E-5 from the outcome GWAS for analysis (r2<0.81), which may result in slight differences in the outcomes.

### 2 Simulation

Now we show how did we perform the simulation.

#### 2.1 Data structure

In practice, GTEx only supplies association data between variants and gene expression for SNPs located within $\pm 1$ Mb of a gene center. This structure results in missing values for SNPs outside this range. As an example, we determined the missing values for each SNP (ordered by BP position on the y-axis) in a multivariable TWAS analysis involving 30 genes on chromosome 22 (ordered by BP position on the x-axis).

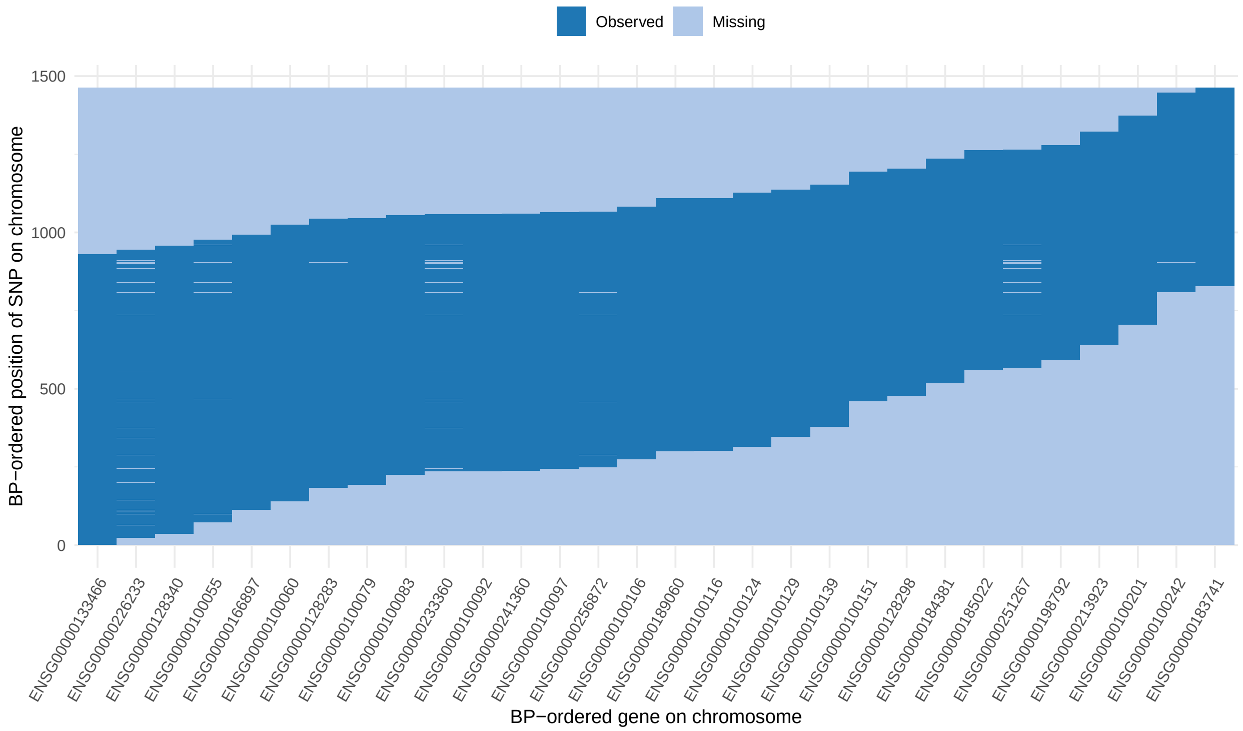

To simulate this scenario, we set the number of variants to $M = 400$ and the number of gene-tissue pairs to $p = 100$, comprising $J = 20$ genes, each with $T = 5$ possible tissues. For each gene-tissue pair, we defined a region spanning 50 variants, within which the gene's eQTLs are located. The sampling regions for eQTLs across different gene-tissue pairs are shown below, with some overlap between them:

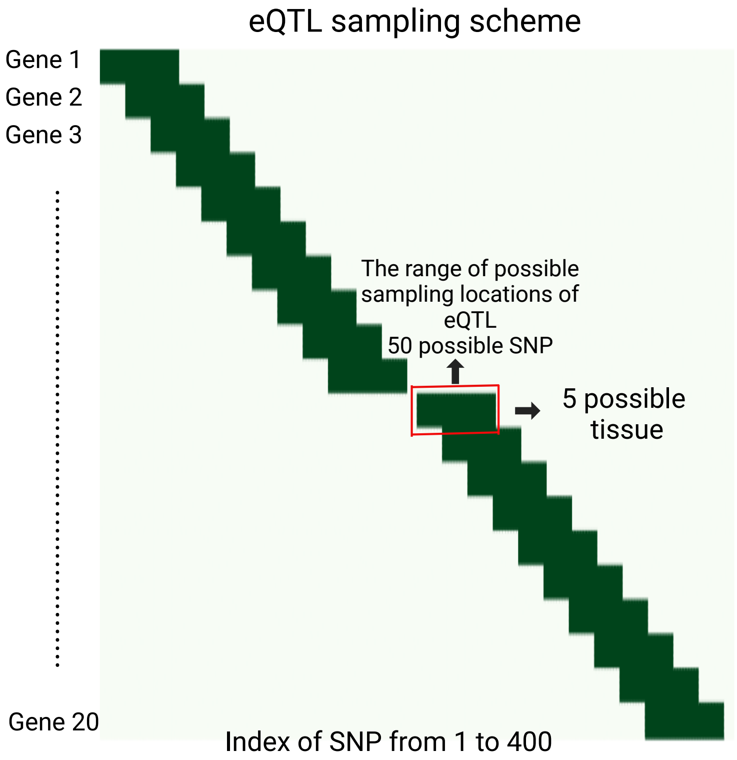

Our next step is to generate correlated direct eQTL effects for each gene-tissue pair. Given the sparsity of potential eQTLs, generating these correlated eQTL effects is quite challenging. Our generation procedure is as follows: For each variant, we generate a latent variable of dimension $p = 100$, denoted as $\boldsymbol{\omega}_j \sim \mathcal{N}(\mathbf{0}, \boldsymbol{\Sigma}_\omega)$, where $\Sigma_\omega$ is a matrix with a Kronecker product structure. This matrix is constructed using the code `Somega=kronecker(ARcov(p=20,-0.5),CScov(p=5,0.5))`, and its structure is as follows:

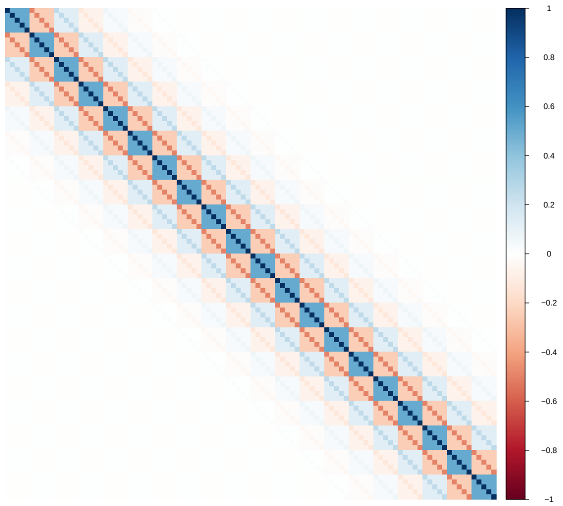

This matrix attempts to simulate the following scenario: For a given gene, the corresponding normal latent variable $\boldsymbol{\omega}_j$ exhibits a compound symmetry structure with an internal correlation of $\rho = 0.5$ within the gene. Between different genes, the correlation follows an order-1 autoregressive (AR(1)) structure with a correlation coefficient of $\rho = -0.5$. As a result, genes that are farther apart have lower correlations, while those that are closer together have higher correlations.

We generate a total of $M' = 50$ such latent variables, $\boldsymbol{\omega}_1, \ldots, \boldsymbol{\omega}_{50}$, since for each gene-tissue pair, the possible range for eQTLs is limited to 50 variants (represented by the dark green area in the figure). We define $\boldsymbol{\Omega} = (\boldsymbol{\omega}_1^\top, \boldsymbol{\omega}_2^\top, \ldots, \boldsymbol{\omega}_{50}^\top)^\top$ as a $50 \times 100$ matrix, where $\boldsymbol{\omega}^j$ represents the $j$-th column of this matrix (and $\boldsymbol{\omega}_j$ is its $j$-th row).

When setting the number of causal eQTLs to $k$ (e.g., $k = 2$), we retain the $k$ largest entries in $\boldsymbol{\omega}^j$ and set the others to 0. We then position this $M' = 50$-dimensional vector within the corresponding possible sampling range for this gene-tissue pair, with all other positions set to 0. Due to the correlations between columns, the top $k$ largest entries are likely to be shared, resulting in genetic correlation. The figure below shows the genetic correlation matrix between these 100 gene-tissue pairs in a single simulation, calculated as follows:

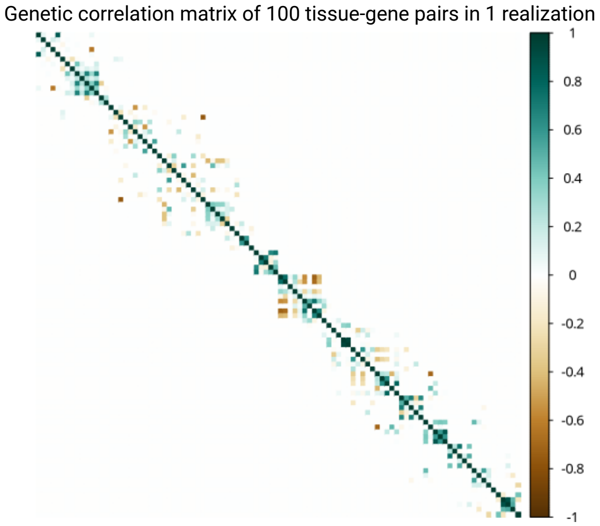

The final one is the LD matrix. The structure of the LD matrix is also of Kronecker product. In practice, we apply $r^2 = 0.5$ within the C+T model, meaning that the maximum correlation scale is approximately 0.7. We generate the LD matrix using the command `LD=kronecker(ARcov(20,0.7),CScov(20,0.5))`, which implies that the LD matrix is block-wise. Within each block, the correlation follows a compound symmetry structure with an internal correlation of $\rho = 0.5$. Additionally, there is correlation between blocks, following an AR(1) structure with a correlation coefficient of $\rho = 0.7$. We consider a total of 20 blocks, each with a size of 20. A visualization of this matrix is shown below:

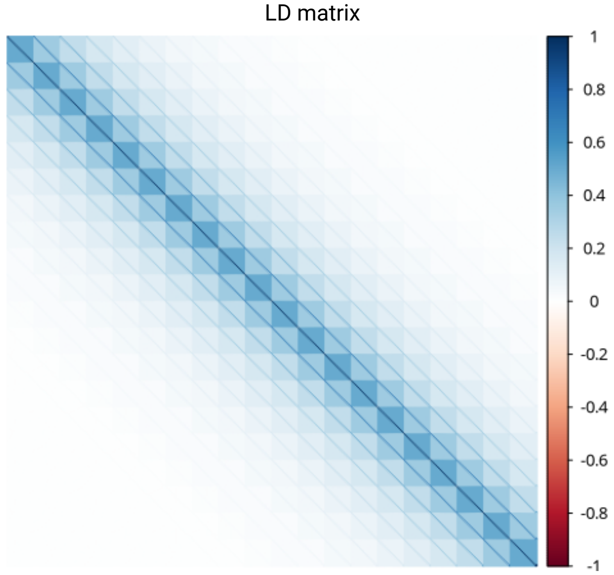

#### 2.2 Step-by-step simulation

We will demonstrate the simulation steps one by one. First, we begin with the script setup.

```
source("function_in_simulation.R")
M=400
p=100
n0=0.5e6 # sample size of outcome GWAS
n1=200 # sample size of eQTL
hx=rep(0.3,100) # fix the heritability of gene-tissue pair 0.3
hy=0.005 # fix the local heritablility of outcome to be 0.005
UHP=1
## unbalanced horizontal pleiotropy
## equivalent to direct causal variants
UHPh2=1
## variance of UHP compared to gene-tissue pair
BHP=1
## balanced horizontal pleiotropy
## equivalent to infinitesimal effect
BHPh2=1
## variance of BHP compared to gene-tissue pair
non.zero.frac=0.06
## The fraction of eQTL (number = 0.06*50=3)
```

Next, we generate the data using the following codes:

```
simuCis=as.matrix(readRDS("simueQTL.rds"))
## The range of variants in sampling eQTL
LD=kronecker(ARcov(20,0.7),CScov(20,0.5))
## The LD matrx
C=matrixsqrt(LD)$w
## The square root of the LD matrix: LD = C%*%C
Somega=kronecker(ARcov(p=20,-0.5),CScov(p=5,0.5))
## The correlation matrix of latent variables
Sxx=ARcov(p=p,-0.5);
## The correlation matrix of estimation error of eQTL summary data
theta0=rep(0,p);theta0[1]=1;theta0[p]=-1
## Generate the causal effects of gene-tissue pairs
B=MASS::mvrnorm(M,rep(0,p),Somega)
## Generate genetic effect
G=cis.sparse(n=nrow(B)/8,p=ncol(B),rho=non.zero.frac,
         Cis=simuCis,Sbb=Somega,min.frac=0.999*non.zero.frac)
barplot(G[,1])
```

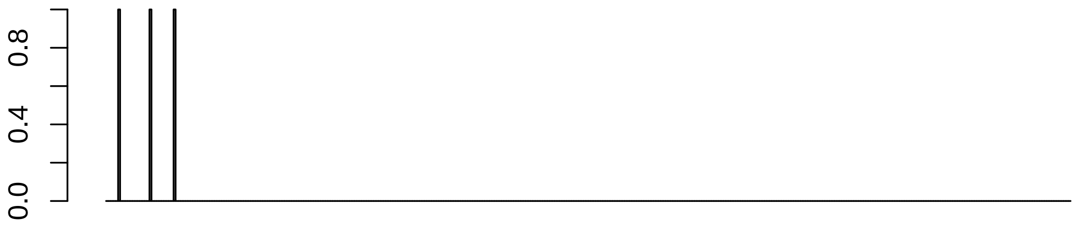

```
## Generate the location of eQTL. 1 means there is a eQTL
```

The next step is rescaling the genetic effect such that `sum(B[,i]^2)=hx=0.3`:

```
B=B*G
B=h.standard(B,h=hx)
c(sum(B[,1]^2),sum(B[,100]^2))
```

```
## [1] 0.3 0.3
```

```
generate.error=verrorvar(Sbb=Somega,Suu=Sxx,Suv=rep(0,p),hx=hx,theta=theta0,
                         hy=hy,pleiotropy.var=UHPh2)
## Suv=rep(0,p) means the eQTL data and outcome GWAS data are nearly independent
Svv=generate.error$Syy
Duv=c(rep(1,p),sqrt(Svv))
Sigmauv=diag(Duv)%*%Matrix::bdiag(Sxx,1)%*%diag(Duv)
Vxy=as.matrix(Sigmauv)
```

We directly generate the GWAS summary data and eQTL summary data based on the model (2) and (3) shown in the main body of the paper. Consider a matrix normal distribution $\mathcal{MN}(\mathbf{M}, \mathbf{G}_1, \mathbf{G}_2)$, where $\mathbf{M}$ is the mean matrix, $\mathbf{G}_1$ is the covariance matrix of rows of any random matrix following this distribution, and $\mathbf{G}_2$ is the covariance matrix of columns. This distribution is essentially equalization to a standard multivariate normal distribution:

$$\mathbf{X} \sim \mathcal{MN}(\mathbf{M}, \mathbf{G}_1, \mathbf{G}_2) \iff \mathtt{vec}(X) \sim \mathcal{N}(\mathtt{vec}(\mathbf{M}), \mathbf{G}_1 \otimes \mathbf{G}_2).$$

The main model in simulation is

$$\hat{\mathbf{A}} = (\hat{\boldsymbol{a}}, \hat{\boldsymbol{b}}_1, \ldots, \hat{\boldsymbol{b}}_p) \sim \mathcal{MN}(\mathbf{R}(\boldsymbol{\alpha}, \boldsymbol{\beta}_1, \ldots, \boldsymbol{\beta}_p), \mathbf{R}, \boldsymbol{\Sigma}_{xy}),$$

where

$$\boldsymbol{\alpha} = \sum_{j=1}^{p} \boldsymbol{\beta}_j \theta_j + \boldsymbol{\gamma} + \boldsymbol{\upsilon}.$$

In this model, $\boldsymbol{\Sigma}_{xy}$ represents the estimation error covariance matrix among $\hat{\boldsymbol{a}}, \hat{\boldsymbol{b}}_1, \ldots, \hat{\boldsymbol{b}}_p$. Lorincz-Comi et al. have provided a detailed expression of this matrix, which depends on (1) the covariance matrix of $\boldsymbol{y}, \boldsymbol{x}_1, \ldots, \boldsymbol{x}_p$, and (2) the sample overlap between the GWAS summary data and the eQTL summary data. According this summarized-statistics based model, we generate the summarized data as follows:

```r
E=MASS::mvrnorm(M,rep(0,p+1),Vxy);
E=t(t(E)-colMeans(E))
E=E%*%diag(sqrt(c(rep(1/n1,p),1/n0)))
E1=C%*%E
bX=LD%*%B+E1[,1:p]
by=LD%*%B%*%theta0+E1[,p+1]
bX=as.matrix(bX)*simuCis
by=as.vector(by)
plot(bX[which(bX[,1]!=0),1],by[which(bX[,1]!=0)])
```

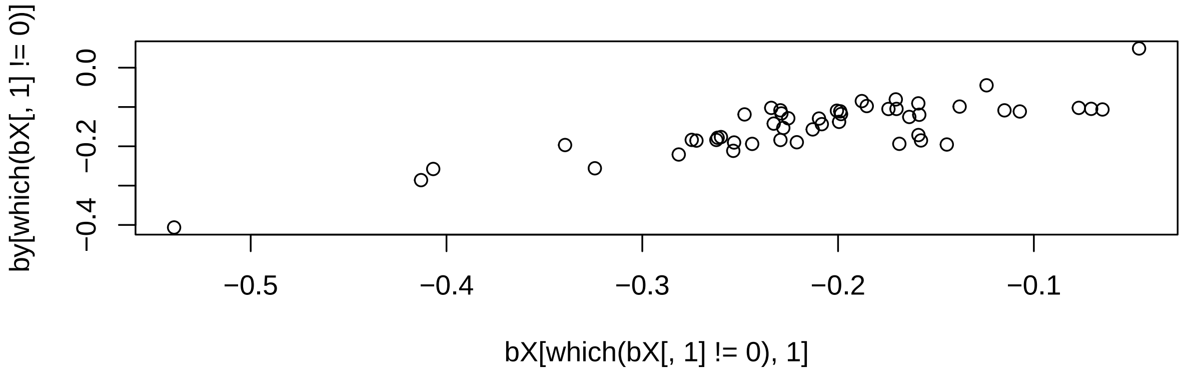

Next, we generate the UHP (direct causal effects) and BHP (infinitesimal effects). For the UHP, we assume they are of the same size:

```r
uhp_effect=rep(0.5,M)
UHPind=sample(which(rowSums(B==0)==100),2)
## We only generate UHP for the variants without being eQTL of any gene-tissue pair
uhp_effect[-UHPind]=0
barplot(uhp_effect)
```

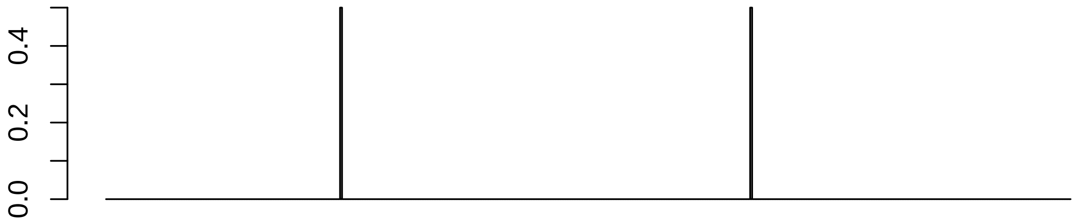

```
if(UHP==1){
s1=as.vector(B%*%theta0)
ratio=var(uhp_effect)/var(s1)/UHPh2
by=by+c(LD%*%uhp_effect/sqrt(ratio))
}
bhp_effect=rnorm(M,0,1)
if(BHP==1){
if(UHP==1){bhp_effect[which(uhp_effect!=0)]=0}
s1=as.vector(B%*%theta0)
ratio=var(bhp_effect)/var(s1)/BHPh2
by=by+c(LD%*%bhp_effect/sqrt(ratio))
}
```

Next, we converted the marignal effect estimates to Z-scores:

```
byse=rep(sd(E1[,p+1]),M)
bXse=matrix(1,M,p)
for(j in 1:p) bXse[,j]=sd(E1[,j])
adjust.coef=bXse*(1/byse)
adjust.coef=adjust.coef[1,]
bX=bX/bXse
bXse=bXse/bXse
by=by/byse
byse=byse/byse
plot(bX[which(bX[,1]!=0),1],by[which(bX[,1]!=0)])
```

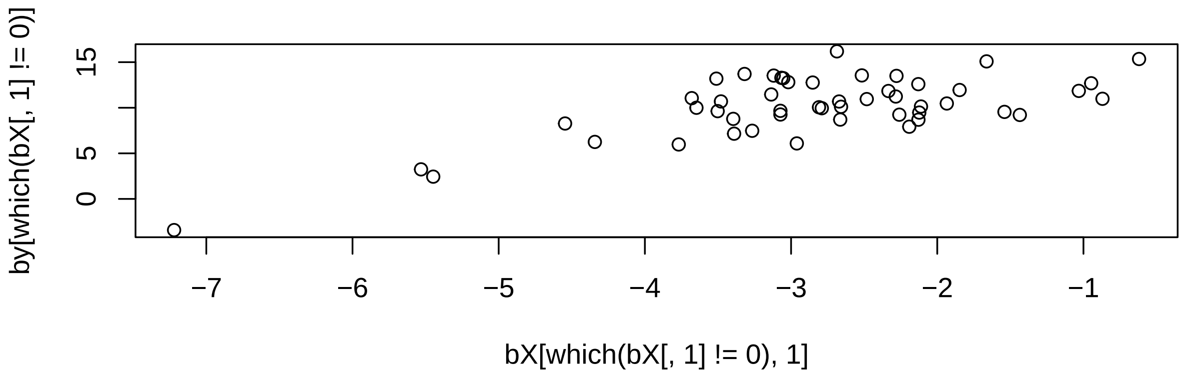

Note that the using Z-score rather than the effect size estimates will res-cale the causal effect, and the principle is

$$y = x\beta + \epsilon \Rightarrow \frac{y}{\|y\|_2} = \frac{x}{\|x\|_2}\beta^* + \epsilon,$$

where $\beta^* = \beta \times \|x\|_2/\|y\|_2$. We record this ratio $\|x\|_2/\|y\|_2$ and re-scale $\hat{\beta}^*$ when evaluating the results.

#### 2.3 Performing the five methods

We first performing the *cis*IVW method using the R package `MendelianRandomization` (Yavorska and Burgess, 2017):

```r
library(MendelianRandomization)
datainput=mr_mvinput(bx=as.matrix(bX),by=by,bxse=as.matrix(bXse),
                  byse=byse,correlation=as.matrix(LD))
fitivw=mr_mvivw(datainput,correl=T)
barplot(fitivw@Estimate/adjust.coef,main="CisIVW")
```

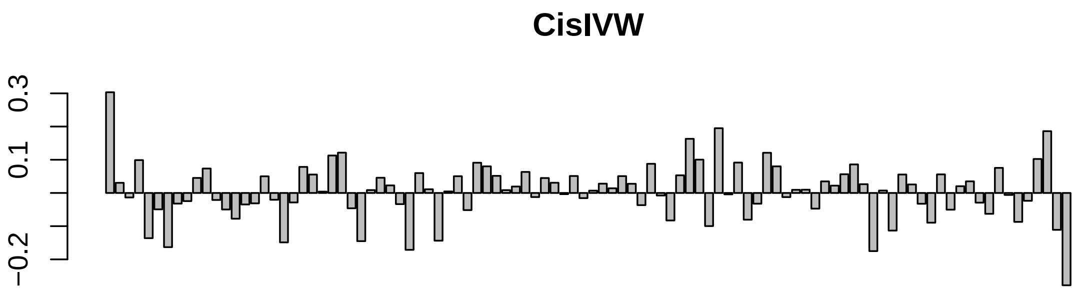

As for Grant2022, we use the R package `ncvreg` (Breheny and Huang, 2011) to perform lasso:

```r
library(ncvreg)
## ncvreg is very simlar to glmnet
## ncvreg directly provides the BIC of each model
fitlasso=ncvreg(X=bX,y=by,penalty="lasso")
barplot(fitlasso$beta[-1,which.min(BIC(fitlasso))]/adjust.coef,main="Grant2022")
```

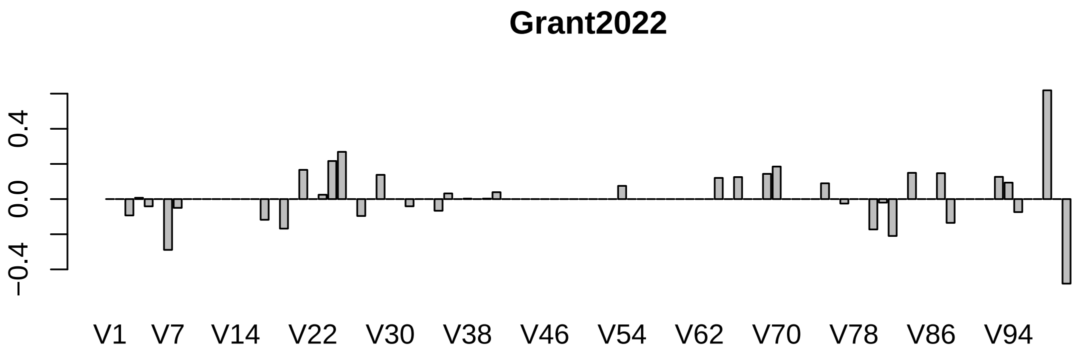

As for TGFM,

```r
library(TGVISelector)
```

```
##
## Attaching package: 'TGVISelector'
```

```
## The following objects are masked from 'package:TGVIS':
##
##     allele_harmonise, ctwas, eQTLmapping_susie, make_design_matrix,
##     modified_predixcan, poet_shrinkage, R2_partition,
##     remove_missing_row_column, tgfm
```

```r
fittgfm=tgfm(by,bX,LD,Nvec=c(n0,rep(n1,p)),
         L.causal=6,L.eqtl=sum(B[,1]!=0)+1,
         causal.sampling.time=25,eqtl.sampling.time=100)
barplot(fittgfm$theta,main="TGFM")
```

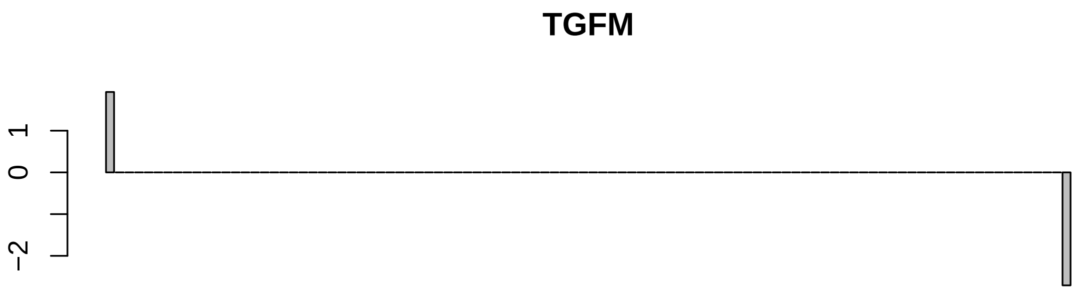

As for cTWAS and TGVIS, we need to first select the eQTL for each gene-tissue pairs:

```r
fiteQTL=eQTLmapping_susie(bX=bX,LD=LD,Nvec=rep(n1,p),pip.thres=0.5,L=sum(B[,1]!=0)+1,pip.min=0.2)
```

```
##   |                                                                              |
```

```r
bXest=fiteQTL$Estimate
print(sum(B[,1]!=0)+1)
```

```
## [1] 4
```

```r
barplot(bXest[,1])
```

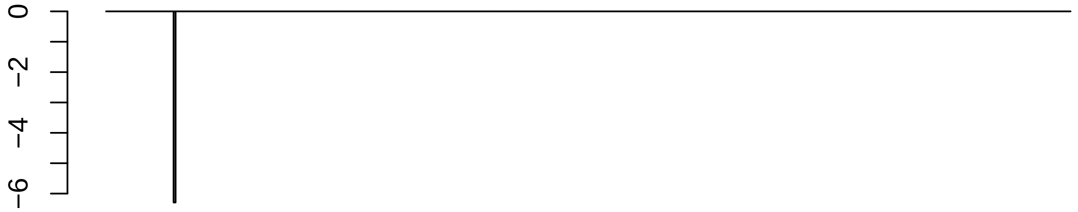

This implies although the true number of eQTL is 3, SuSiE could find less number of eQTL sometimes.

The code of performing cTWAS is

```
fitctwas=ctwas(by,bXest=bXest,LD,Noutcome=n0,L.causal=6)
barplot(fitctwas$theta,main="cTWAS")
```

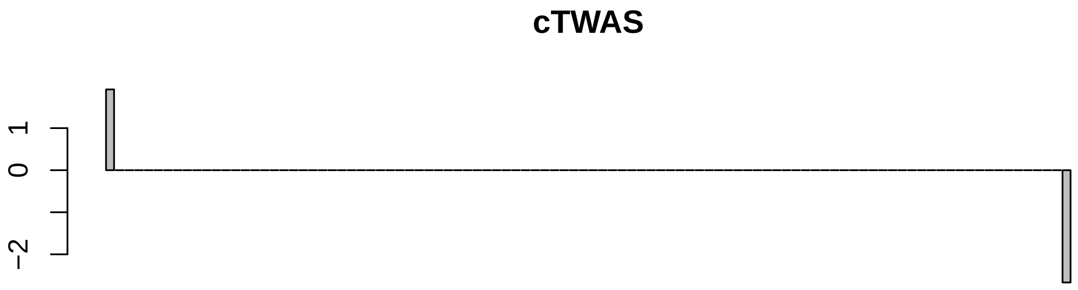

The code for performing TGVIS is:

```
fittgvis=tgvis(by=by,bXest=bXest,LD=LD,Noutcome=n0,
                 pleiotropy.rm=findUniqueNonZeroRows(bXest),
                 L.causal.vec=c(2:6),pip.min=0.05)
barplot(fittgvis$theta,main="TGVIS")
```

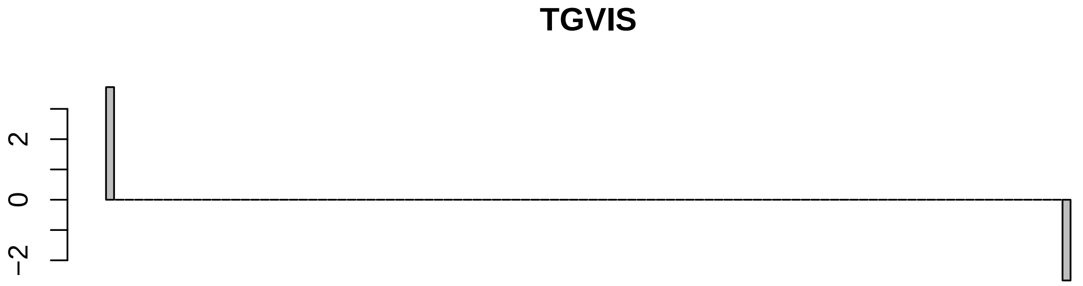

### 3 Descriptions of Functions in the TGVIS Package

In this section, we provide a detailed introduction to the functions within the TGVIS package, based on statistical principles. We will explain the statistical models behind these codes step-by-step to help readers understand our approach.

#### 3.1 cTWAS

We begin with an analysis of the cTWAS function, as cTWAS serves as the prototype for both TGFM and TGVIS.

```
ctwas <- function(by, bXest, LD, Noutcome, L.causal = 10, pip.thres.cred = 0.95)
```

The cTWAS method has only six inputs. by represents the Z-scores of the outcome GWAS. bXest is the estimated eQTL effect sizes obtained using SuSiE (or other methods). LD is the linkage disequilibrium matrix, Noutcome is the sample size of the outcome GWAS, L.causal is the total number of potential causal gene-tissue pairs and direct causal variants, and pip.thres.cred is the CS-PIP threshold, which we set at 0.95.

```
n <- length(by)
p <- dim(bXest)[2]
Theta <- matrixInverse(LD)
XR <- cbind(matrixMultiply(LD, bXest), LD)
Xty <- c(t(bXest) %*% by, by)
XtX <- Matrix::bdiag(matrixMultiply(t(bXest) %*% LD, bXest), LD)
XtX <- as.matrix(XtX)
XtX[1:p, -c(1:p)] <- matrixMultiply(t(bXest), LD)
XtX[-c(1:p), c(1:p)] <- t(XtX[1:p, -c(1:p)])
dXtX <- diag(XtX)
dXtX[is.na(dXtX)] <- 1
dXtX[dXtX == 0] <- 1
XtX[is.na(XtX)] <- 0
diag(XtX) <- dXtX
Xadjust <- diag(XtX)
XtX <- cov2cor(XtX)
```

```
XtX=(t(XtX)+XtX)/2
XtyZ <- Xty / sqrt(Xadjust)
```

The main purpose of the above code is to compute the inputs for `susie_rss()`, specifically: `z` and `R`. More specifically, we consider the design matrix or covariance matrix $\mathbf{G} = (\mathbf{R}\hat{\mathbf{B}}, \mathbf{R})$, where $\hat{\mathbf{B}} = (\hat{\beta} jt) M \times JT$represents the eQTL effect sizes estimated using SuSiE. The inner product of this design matrix is $\mathbf{G}^\top \mathbf{R}^{-1} \mathbf{G}$. On the other hand, the marginal covariance Z-scores between $\mathbf{G}$ and $\mathbf{y}$ are calculated as $\mathbf{G}^\top \hat{\boldsymbol{\alpha}} / \mathrm{diag}(\mathbf{G}^\top \mathbf{R}^{-1} \mathbf{G})$. In the code, $\hat{\boldsymbol{\alpha}}$corresponds to `by`, $\mathbf{G}$ corresponds to `XR`, and $\mathbf{G}^\top \hat{\boldsymbol{\alpha}} / \mathrm{diag}(\mathbf{G}^\top \mathbf{R}^{-1} \mathbf{G})$ corresponds to `XtyZ`, while `XtX` corresponds to $\mathbf{G}^\top \mathbf{R}^{-1} \mathbf{G}$. The rest of the code ensures no bugs occur.

```
prior.weight.theta <- rep(1 / p, p)
prior.weight.gamma <- rep(1 / n, n)
prior_weights <- c(prior.weight.theta, prior.weight.gamma)
```

It ensures that gene-tissue pairs and direct causal variants have different prior distributions of $\boldsymbol{\pi}$.

```
fit.causal <- susie_rss(z = XtyZ, R = XtX, n = Noutcome, L = L.causal,
                        residual_variance = 1, estimate_prior_method = "EM",
                        prior_weights = prior_weights, intercept = FALSE,
                        max_iter = 300)
```

This code uses `susie_rss` to estimate gene-tissue causal effects and direct causal effects. We use the EM algorithm to estimate the prior distributions of these effects, restricting the estimation to the locus of interest. In contrast, Zhao et al. (2024) appears to estimate two universal distributions across all loci. This is the key difference between our `ctwas` function and the approach in Zhao et al. (2024).

```
fit.causal.sampling <- susie.resampling(alpha = fit.causal$alpha,
                                mu = fit.causal$mu, mu2 = fit.causal$mu2)
fit.causal$beta.se <- fit.causal.sampling$sd
```

The remaining part of this function is dedicated to preparing the output. One important point to highlight is that we estimate the standard errors using resampling. The R program SuSiE provides three$L \times p$dimensional matrices: $\mathbf{A}$, $\mathbf{Mu}$, and $\mathbf{Mu2}$, which are used to estimate the standard errors of the involved parameter estimates. We applied the same resampling scheme as Strober et al. (2023), generating $\tilde{\delta}_l \sim \mathrm{Multi}(\mathbf{A}_l)$, where $\mathbf{A}_l$ is the $l$-th row of $\mathbf{A}$. Next, we generated $\tilde{b}_{jl} \sim N(\mathbf{Mu}_{jl}, \mathbf{Mu2}_{jl} - \mathbf{Mu}_{jl}^2)$. Finally, we obtained a posterior sample of $\beta_j$ by:

$$\tilde{\beta}_j = \sum_{l=1}^{L} \tilde{b}_{jl} \tilde{\delta}_{jl},$$

We estimated the empirical standard deviation as the standard deviation of $\hat{\beta}_j$ from the independent sampling scheme. This approach is also applied to $\hat{\theta}_{jt}$and$\hat{\gamma}_m$once the iteration converges. Since this is not computationally expensive, we performed 100 resamplings.

#### 3.2 TGFM

A unique step in TGFM is the resampling using the posterior distributions of the eQTL effect estimates provided by SuSiE. Therefore, unlike cTWAS, TGFM leverages these posterior distributions obtained from SuSiE and stores them for further analysis.

```
for(i in 1:p){
errorindicator <- FALSE
indx <- which(bX[,i] != 0)
eQTLList[[i]]=list(alpha=matrix(0.5,1,p),mu=matrix(0,1,p),mu2=matrix(1,1,p),index.causal=1,indx=indx)
tryCatch({
a <- LD[indx, indx]
fit <- susie_rss(z = bX[indx, i], R = a, n = Nvec[i + 1], L = L.eqtl,  estimate_prior_method="EM")
####################### We don't consider the credible set including too many variables ################
index.causal = intersect(unique(susie_get_cs_index(fit)),which(fit$pip>eqtl.thres))
eQTLList[[i]]=list(alpha=fit$alpha,mu=fit$mu,mu2=fit$mu2,index.causal=index.causal,indx=indx)
}, error = function(e){
cat("Error in iteration", i, ": ", e$message, "\n")
errorindicator <- TRUE
})
if(errorindicator) next
}
```

We found that SuSiE's `susie_get_cs()` sometimes generates credible sets that contain a large number of variables, which we believe are likely generated incorrectly. Therefore, we use `index.causal = intersect(unique(susie_get_cs_index(fit)), which(fit$pip > eqtl.thres))` to remove these variables. The `eqtl.thres` is typically set to a very low value, and we have set it to 0.01 in this context.

Our next step is to perform resampling. To improve computational efficiency, we resample the eQTL effect sizes according to the number of PIP resampling iterations, generating eQTL effect sizes for each resampling iteration using a for loop. These eQTL effect sizes are averaged over the number of eQTL resampling iterations. We set the number of PIP resampling iterations to 100, with 25 resampling iterations for each eQTL effect size. Finally, we calculate the average effect size from the 100 PIP resampling iterations. This effect size is then used in the overall TGFM estimation.

```
ZArray= array(0,c(n,p,causal.sampling.time))
ZAll= matrix(0,n,p)
colnames(ZAll)=colnames(bX)
rownames(ZAll)=rownames(bX)
for (j in 1:p){
z = bX[eQTLList[[j]]$indx, 1] * 0
indj = unique(eQTLList[[j]]$index.causal)
z = tgfm.resampling(alpha = eQTLList[[j]]$alpha, mu = eQTLList[[j]]$mu,
                    mu2 = eQTLList[[j]]$mu2, sampling =
                    causal.sampling.time*eqtl.sampling.time) * sqrt(Nvec[j + 1])
zall = colMeans(z)
zall[abs(zall)<eqtl.thres]=0
if(length(indj)>0){
zall[-indj]=0
}
ZAll[eQTLList[[j]]$indx,j]=zall
for(jj in 1:causal.sampling.time){
indjj=c((eqtl.sampling.time*(jj-1)+1):(eqtl.sampling.time*jj))
zjj=z[indjj,]
zjj=colMeans(zjj)
zjj[abs(zjj)<eqtl.thres]=0
if(length(indj)>0){
zjj[-indj]=0
}
ZArray[eQTLList[[j]]$indx,j,jj]=zjj
```

```
}
}
```

Below is the main function of TGFM.

```
eXi = ZAll
Xty = c(t(eXi) %*% by, by)
XtX = Matrix::bdiag(matrixMultiply(t(eXi) %*% LD, eXi), LD)
XtX = as.matrix(XtX)
XtX[1:p, -c(1:p)] = matrixMultiply(t(eXi), LD)
XtX[-c(1:p), c(1:p)] = t(XtX[1:p, -c(1:p)])
dXtX = diag(XtX); dXtX[is.na(dXtX)] = 1; dXtX[dXtX == 0] = 1;
XtX[is.na(XtX)] = 0; diag(XtX) = dXtX
Xadjust=diag(XtX)
XtX=cov2cor(XtX)
XtX=(t(XtX)+XtX)/2
XtyZ=Xty/sqrt(Xadjust)
prior.weight.theta=rep(1/p,p)
prior.weight.gamma=rep(1/n,n)
prior_weights=c(prior.weight.theta,prior.weight.gamma)
fit.causal = susie_rss(z=XtyZ,R=XtX,n=Nvec[1], L = L.causal,
                       residual_variance = 1, estimate_prior_method="EM",
                       prior_weights=prior_weights, intercept=F,max_iter=300)
```

Next, we apply susie_rss to the data recorded from the previous PIP resampling. For computational efficiency, we limit each resampling iteration to a maximum of 10 iterations.

```
AA = AB = matrix(0, causal.sampling.time,n+p)
for (i in 1:causal.sampling.time) {
eXi = ZArray[,,i]
Xty = c(t(eXi) %*% by, by)
XtX = Matrix::bdiag(matrixMultiply(t(eXi) %*% LD, eXi), LD)
XtX = as.matrix(XtX)
XtX[1:p, -c(1:p)] = matrixMultiply(t(eXi), LD)
XtX[-c(1:p), c(1:p)] = t(XtX[1:p, -c(1:p)])
dXtX = diag(XtX); dXtX[is.na(dXtX)] = 1; dXtX[dXtX == 0] = 1;
XtX[is.na(XtX)] = 0; diag(XtX) = dXtX
Xadjusti=diag(XtX)
XtX=cov2cor(XtX)
XtX=(t(XtX)+XtX)/2
XtyZ=Xty/sqrt(Xadjusti)
fit.causali = susie_rss(z=XtyZ,R=XtX,n=Nvec[1],L=L.causal,
                       estimate_prior_method="EM",s_init=fit.causal,
                       prior_weights=prior_weights,intercept=F,max_iter=10)
AA[i,] = fit.causali$pip
AB[i,] = coef(fit.causali)[-1]
}
```

It is important to note that the credible sets generated in the resampling process will not necessarily match those produced by the main TGFM function. We only record the individual PIPs from each resampling iteration, and then use the average of these PIPs to infer the CS-PIP in the main TGFM procedure.

The remaining steps are consistent with those in cTWAS, so they are not discussed here.

##### 3.3 TGVIS

TGVIS has some specific inputs, such as `L.causal.vec = c(1:6)`, which indicates that we consider up to 6 gene-tissue pairs or direct causal variants. After accounting for the infinitesimal effect, we rarely observe credible sets with more than 5 variables in practice.

We do not want the infinitesimal effect to characterize the majority of the outcome's local genetic variants; therefore, we set varinf.upper.boundary = 0.25. This indicates that the prior variance of the infinitesimal effect cannot exceed one-quarter of the local genetic variance of the outcome.

```
varinf.upper.boundary=varinf.upper.boundary*sum(by*(Theta%*%by))/n
```

The difference from cTWAS is that the inputs to `susie_rss` for TGVIS are:

```
res.beta=by-matrixVectorMultiply(LD,upsilon)
Xty=c(t(bXest)%*%res.beta,res.beta[pleiotropy.keep])/sqrt(XtXadjust)
fit.causal=susie_rss(z=XtyZ,R=XtX,n=Noutcome,L=max(1,L.causal.vec[i]),
                     estimate_prior_method="EM",max_iter=inner.iter,
                     intercept=F,standardize=F,prior_weights=prior_weights)
```

Here, `pleiotropy.keep` represents the indices of variants we consider having a direct causal effect. We do not consider a variant to have a direct causal effect if it is the only eQTL for a particular gene-tissue pair.

Next, we explain how to estimate the infinitesimal effect. We first use a score test to determine whether the infinitesimal effect needs to be considered. Since the score test requires specifying fixed effects, we force all variables not included in the 95% credible set to be set to zero.

```
causal.cs=group.pip.filter(pip.summary=summary(fit.causal)$var,pip.thres.cred=pip.min)
pip.alive=causal.cs$ind.keep
beta[-pip.alive]=0
res.upsilon=by-matrixVectorMultiply(XR,beta)
```

Next, we perform the score test. We only consider the infinitesimal effect if the score test p-value is below the threshold or if the infinitesimal effect was considered in the first five iterations.

```
pv=inf.test(res.inf=res.upsilon,LD=LD,LD2=LD2,Theta=Theta,A=XR[,which(fit.causal$pip>pip.min)])
######################## Performing REML ###############################
upsilon=by*0
if(pv<pv.thres|iter<5){
for(ii in 1:3){
Hinv=1/(Dvec+1/varinf)
upsilon=matrixVectorMultiply(Umat,outcome*Hinv)
for(jj in 1:3){
df=sum(Hinv)
varinf=min((sum(upsilon^2)+df)/n,varinf.upper.boundary)
pv=ifelse(varinf<varinf.lower.boundary,0.5,pv)
}
}
}
error=norm(beta-beta1,"2")/sqrt(length(beta))
iter=iter+1
}
```

Next, we perform the BIC calculation. Specifically, we use the extended BIC Chen and Chen (2012), which is defined by the formula:

$$\text{EBIC} = \log\hat{\sigma}_\alpha^2 + \frac{\log M + f_1\log(p+M)}{M}\text{df}_1 + \frac{\log M + f_2\log M}{M}\text{df}_2,$$

where$\text{df}_1$is the number of gene-tissue pairs and direct causal variants, i.e.,$L$, while$\text{df}_2$is the degrees of freedom for the infinitesimal effect, defined as:

$$\text{df}_2 = \text{trace}\bigg(\Big(\mathbf{R} + \frac{1}{\hat{\sigma}_\nu^2}\mathbf{I}\Big)^{-1}\mathbf{R}\bigg).$$

```
df=sum(Dvec*Hinv)
res=by-matrixVectorMultiply(XR,beta)-matrixVectorMultiply(LD,upsilon)
rss=sum(res*matrixVectorMultiply(Theta,res))
Bicvec[i]=log(rss)+(log(n)+ebic.beta*log(dim(XtX)[1]))/n*L.causal.vec[i]
          +(ebic.upsilon*log(n)+log(n))/n*df
```

The rest code is identical to that in cTWAS and TGFM.

#### 3.4 S-Predixcan and Its Modifier

To prioritize potentially causal gene-tissue pairs, we perform a univariable TWAS analysis using S-Predixcan on all gene-tissue pairs within each region. This step helps filter out gene-tissue pairs that are unlikely to have a causal relationship with the trait of interest.

First, we define$\vartheta_{jt}$as the marginal effect in the univariable TWAS model, which is given by:

$$\vartheta_{jt} = \sum_{j'=1}^{J}\sum_{t'=1}^{T}\left(\frac{1}{M}\beta_{j't'}^\top\beta_{jt}\right)\theta_{j't'},$$

due to the correlation term $\frac{1}{M}\beta_{j't'}^\top\beta_{jt}$. The S-Predixcan estimate is calculated as:

$$\hat{\vartheta}_{jt} = \frac{\hat{\beta}_{jt}^\top\hat{a}}{\hat{\beta}_{jt}^\top R\hat{\beta}_{jt}},$$

and its covariance is:

$$\text{var}(\hat{\vartheta}_{jt}) = \frac{(\hat{a} - R\hat{\beta}_{jt}\hat{\vartheta}_{jt})^\top R^{-1}(\hat{a} - R\hat{\beta}_{jt}\hat{\vartheta}_{jt})}{\hat{\beta}_{jt}^\top R\hat{\beta}_{jt}}.$$

We slightly modify the S-Predixcan to account for horizontal pleiotropy, although a similar idea has been considered in the literature. The fine-mapping model employed in this analysis is as follows:

$$\hat{a} \sim N(R\hat{\beta}_{jt}\vartheta_{jt} + R\gamma_{jt}, \sigma_{\alpha'}^2 R),$$

Given an estimate $\vartheta_{jt}^{(s)}$ in the $s$-th iteration, the univariable TWAS screener updates $\gamma_{jt}$ with lasso according to:

$$\hat{a} - R\hat{\beta}_{jt}\vartheta_{jt}^{(s)} \sim N(R\gamma_{jt}, \sigma_{\alpha'}^2 R).$$

Given $\gamma_{jt}^{(s)}$, the univariable TWAS screener updates$\vartheta_{jt}$by:

$$\vartheta_{jt}^{(s)} = \frac{\hat{\beta}_{jt}^\top(\hat{a} - R\gamma_{jt}^{(s)})}{\hat{\beta}_{jt}^\top R\hat{\beta}_{jt}}.$$

The variance of $\hat{\vartheta}_{jt}$is approximated by:

$$\text{var}(\hat{\vartheta}_{jt}) = \frac{(\hat{a} - R\hat{\beta}_{jt}\hat{\vartheta}_{jt} - R\hat{\gamma}_{jt})^\top R^{-1}(\hat{a} - R\hat{\beta}_{jt}\hat{\vartheta}_{jt} - R\hat{\gamma}_{jt})}{\hat{\beta}_{jt}^\top R\hat{\beta}_{jt}}.$$

Note that for univariable TWAS analysis, we do not consider the infinitesimal effect, because the goal is just to provide roughly accurate statistics that can screen the noncausal gene-tissue pairs. Also, we call this variance approximated because it seems underestimated due to multiple uncontrolled issues such as the uncertainty of $\hat{\gamma}_{jt}$. We believe this is acceptable because we use this screener merely to rule out some candidates, thereby reducing the model to a reasonable scale.

In addition, as the real data of GTEx eQTL/sQTL summary data will only provide the summarized statistics within a 1MB region whose center is the transcription start site (TSS), some elements of $\hat{a}$ for a gene-tissue pair are exactly zero because the variants are selected within a 2MB region whose center is the GWAS hit. Hence, we only analyzed the non-zero part of $\hat{a}$:

$$\hat{a}_{M_{jt}} \sim N(R_{M_{jt}M_{jt}}\hat{\beta}_{M_{jt}}\vartheta_{jt} + R_{M_{jt}M_{jt}}\gamma_{M_{jt}}, \sigma_{\alpha'}^2 R_{M_{jt}M_{jt}}), \quad (52)$$

where the notation $M_{jt}$ refers to the index set of non-zero elements in $\hat{\beta}_{jt}$.

```
modified_predixcan=function(by,bxest,LD,tauvec=seq(3,10,by=1),
                        rho.gamma=1.5,max.iter=15,max.eps=0.005,
                        ebic.factor=2,normmax=2,pleiotropy.rm=NULL){
n=length(by)
pleiotropy.keep=setdiff(c(1:n),pleiotropy.rm)
dx=matrixVectorMultiply(LD,bxest)
Theta=matrixInverse(LD)
yinv=c(matrixVectorMultiply(Theta,by))
xtx=sum(bxest*dx)
xty=sum(by*bxest)
theta.ini=xty/xtx
Thetarho=matrixInverse(LD[pleiotropy.keep,pleiotropy.keep]
          +rho.gamma*diag(length(pleiotropy.keep)))
tauvec=sort(tauvec,decreasing=F)
w=length(tauvec)
Btheta=c(1:w)
Bgamma=matrix(0,n,w)
Bbic=c(1:w)
for(sss in c(w:1)){
theta=theta.ini
gamma=by*0
gamma1=gamma
u=rho.gamma*(gamma-gamma1)
theta1=theta*0
error=1
iter=1
while(error>max.eps&iter<max.iter){
```

```
theta1=theta
theta=(xty-sum(dx*gamma))/xtx
res=c(by-dx*theta-u+rho.gamma*gamma1)
gamma[pleiotropy.keep]=c(matrixVectorMultiply(Thetarho,res[pleiotropy.keep]))
gamma1=mcp(gamma+u/rho.gamma,tauvec[sss],ga=3)
u=u+rho.gamma*(gamma-gamma1)
u[pleiotropy.rm]=0
iter=iter+1
if(iter>3){
error=abs(theta-theta1)
}
Btheta[sss]=theta
Bgamma[,sss]=gamma1
r=vec(by-dx*theta-matrixVectorMultiply(LD,gamma1))
df=sum(gamma1!=0)
rss=sum(r*(matrixVectorMultiply(Theta,r)))
Bbic[sss]=n*log(rss)+log(n)*(1+ebic.factor)*df
}
}

star=which.min(Bbic)
theta=Btheta[star]
gamma=Bgamma[,star]
eta=bxest*theta

indgamma=which(gamma!=0)
effn=n-length(indgamma)
if(sum(indgamma)>0){
Z=cbind(bxest,diag(n)[,indgamma])
Hinv=matrixMultiply(t(Z),matrixMultiply(LD,Z))
Hinv=MASS::ginv(Hinv)
r=vec(yinv-bxest*theta-gamma)
varr=mean(r*matrixVectorMultiply(LD,r))*n/(effn-1)
covg=varr*Hinv
covg=covg[1,1]
covtheta=covg
}
if(sum(indgamma)==0){
r=vec(yinv-bxest*theta)
varr=mean(r*matrixVectorMultiply(LD,r))
covg=varr/xtx*n/(n-1)
covtheta=covg
}

A=list()
A$theta=theta
A$gamma=gamma
A$covtheta=as.numeric(covtheta)
A$Bic=Bbic
A$Btheta=Btheta
A$Bgamma=Bgamma
A$Eta=eta
return(A)
}
```

##### 3.5 POET Shrinkage Estimate

We applied regularization to the high-dimensional LD matrix. Our motivation was that in some regions with stronger LD, the LD matrix estimated from the reference panel with approximately 10K individuals has a larger condition number (the ratio of the maximum eigenvalue to the minimum eigenvalue). Specifically, we utilized a modification of Principal Orthogonal Complement Thresholding (POET) (Fan et al., 2013) to address this issue. POET considers an eigenvalue decomposition of the sample LD matrix of individuals' genotypes:

$$\hat{R} = \sum_{k=1}^{p} d_k U_k U_k^\top = \sum_{k=1}^{K} d_k U_k U_k^\top + E,$$

where $d_1 \geq d_2 \geq \cdots \geq d_p$are the eigenvalues of $\hat{R}$, $U_k$is the corresponding eigenvector of $d_k$,$K$is a cutoff, and$E$is the residual matrix. To improve the condition of $\hat{R}$, the standard POET applies a covariance-thresholding method on$E$, while we considered a linear shrinkage of $E$:

$$\tilde{E} = \alpha E + (1 - \alpha)\text{diag}(E).$$

The extended POET estimate was:

$$\tilde{R} = \sum_{k=1}^{K} d_k U_k U_k^\top + \tilde{E}.$$

We use $\tilde{R}$ in the corresponding data.

We utilized the Dynamic Eigenvalue Difference Ratio (DDR) to select $K$ (Cavicchioli et al., 2016):

$$K = \arg \min_{K_{\min} \leq k \leq K_{\max}} \frac{d_k - d_{k+1}}{d_{k+1} - d_{k+2}},$$

where $K_{\min} = 2$ and $K_{\max} = \min(15, p/2)$. On the other hand, we adopted finite sample positive definiteness for the selection of $\alpha$(Fan et al., 2013):

$$\alpha = \inf\{a : \text{minimum eigenvalue of } aE + (1 - a)\text{diag}(E) > \tau\},$$

where $\tau = 0.001$ was a given tolerance.

```
poet_shrinkage=function(LD,KMax=min(15,round(nrow(LD)/2)),
                        lamvec=seq(0.025,0.25,by=0.025),minvalue=1e-3){
LD[is.na(LD)]=0;
diag(LD)=1
LD[abs(LD)<0.0001]=0
eig=matrixEigen(LD)
U=eig$vectors
d=eig$values
z=c()
for(j in 2:KMax){
z[j-1]=(d[j-1]-d[j])/(d[j]-d[j+1])
}
pck=which.max(z)+1
Uk=U[,1:pck];dk=d[1:pck]
```

```
hatc=(sum(diag(LD))-sum(dk))/(ncol(LD)-pck)
P=matrixMultiply(Uk,t(Uk)*dk)
E=LD-P;e=diag(E);e[e<0]=max(hatc,0.01);diag(E)=e
eigenvec=lamvec
for(i in 1:length(lamvec)){
E1=E*(1-lamvec[i])+diag(diag(E))*lamvec[i]
eigenvec=min(matrixEigen(E1)$values)
if(eigenvec>minvalue) break
}
hatLD=P+E1
return(cov2cor(hatLD))
}
```

##### 3.6 Statistical Principle of Infinitesimal Effects

We believe that in an ideal case, infinitesimal effects should not exist. In a local genome region, a few specific variants with cis-regulatory effects influence the outcome through molecular phenotypes like gene expression and splicing events, termed gene-tissue mediated variants. Unspecified components, such as DNA methylation and chromatin accessibility, cause variants to act as direct causal or non-gene-expression mediated variants. While unknown biological mechanisms may underlie infinitesimal effects, we discuss the statistical mechanisms that could generate them.

First, estimation errors in the LD matrix could generate infinitesimal effects. Specifically, let $\mathbf{R}$ be the true LD matrix and $\hat{\mathbf{R}}$ be its estimate. The expectations of outcome GWAS effects without infinitesimal effects is:

$$\mathrm{E}(\hat{\boldsymbol{\alpha}}) = \mathbf{R}(\mathbf{B}\boldsymbol{\theta} + \boldsymbol{\gamma}) = \hat{\mathbf{R}}(\mathbf{B}\boldsymbol{\theta} + \boldsymbol{\gamma}) + (\mathbf{R} - \hat{\mathbf{R}})(\mathbf{B}\boldsymbol{\theta} + \boldsymbol{\gamma}).$$

If $\hat{\mathbf{R}}$is biased or misspecified, $(\mathbf{R} - \hat{\mathbf{R}})(\mathbf{B}\boldsymbol{\theta} + \boldsymbol{\gamma})$ could be non-negligible and act as an infinitesimal effect. This bias term cannot be represented as $\mathbf{R}\boldsymbol{\nu}$. This explains why, for many loci with low local heritability of traits, neither causal gene-tissue pairs nor direct causal variants were identified, and the Pratt index of infinitesimal effects was also low. That is, the low local heritability of the outcome could make the scale of $\mathbf{R}(\mathbf{B}\boldsymbol{\theta} + \boldsymbol{\gamma})$ not significantly larger than $(\mathbf{R} - \hat{\mathbf{R}})(\mathbf{B}\boldsymbol{\theta} + \boldsymbol{\gamma})$, resulting in a low signal-to-noise ratio, and hence causing the TGVIS to fail.

Second, imputation errors in outcome GWAS effect estimates can generate infinitesimal effects. Let $\tilde{\boldsymbol{\alpha}}$ be the actual outcome effect estimates in which some entries are imputed, and $\hat{\boldsymbol{\alpha}}$ be the theoretical outcome effect estimates. Then

$$\mathrm{E}(\tilde{\boldsymbol{\alpha}}) = \mathbf{R}(\mathbf{B}\boldsymbol{\theta} + \boldsymbol{\gamma}) + \mathrm{E}(\tilde{\boldsymbol{\alpha}} - \hat{\boldsymbol{\alpha}}).$$

Also, imputation error (or imputation bias) cannot be represented as$R\nu$and hence can cause the TGVIS to fail in practice.

Third, estimation errors or estimation biases in eQTL effect sizes lead to infinitesimal effects, which is usually caused by the misspecified LD matrices. It has been gradually understood that even within the same ethnic group, such as the European population, different subgroups share different genetic architectures, leading to different LD structures. Therefore, it is natural to suspect that the LD structures of populations in the GTEx consortium and those in traits GWAS differ, which results in

$$\mathrm{E}(\hat{\boldsymbol{\alpha}}) = \mathbf{R}_{\mathrm{Meta}}(\mathbf{B}\boldsymbol{\theta} + \boldsymbol{\gamma}), \quad \mathrm{E}(\hat{b}_{jt}) = \mathbf{R}_{\mathrm{GTEx}}\boldsymbol{\beta}_{jt}.$$

When we try to estimate $\boldsymbol{\beta}_{jt}$ using $\mathbf{R}_{\mathrm{Meta}}$, then $\hat{\boldsymbol{\beta}}_{jt}$is biased relative to$\boldsymbol{\beta}_{jt}$, which generates the infinitesimal effect $\boldsymbol{\nu} = \sum_{jt}(\boldsymbol{\beta}_{jt} - \hat{\beta}_{jt})\boldsymbol{\theta}_{jt}$. It should be noted that the small sample size in the GTEx consortium can also cause biased eQTL effect estimates, resulting in the appearance of infinitesimal effects.

Lastly, the absence of causal variants can cause infinitesimal effects. In the absence of causal variants, $\boldsymbol{\gamma}$ is not a sparse vector. Variants in strong LD with causal variants can be good proxies, while variants in weak LD with causal variants exhibit infinitesimal effects.

### 4 Mendelian Randomization

#### 4.1 Step-by-stey analysis

We provide a case example to illustrate how we performed univariable MR analysis to validate the causal genes identified in cis eQTL/sQTL analysis.

First, the so-called pQTL summary data is essentially the same as the GWAS summary data of a protein, with the following data structure:

```
PCSK9=read_parquet("PCSK9.parquet")
```

```
## Warning: Potentially unsafe or invalid elements have been discarded from R metadata.
## i Type: "externalptr"
## > If you trust the source, you can set ‘options(arrow.unsafe_metadata = TRUE)‘ to preserve them.
```

```
head(PCSK9)
```

```
##      MarkerName A1 A2    Zscore     N           P         SNP CHR        BP
## 1:  10:10041620  C  A  2.965897 34090 0.003018007   rs2762638  10  10083583
## 2:  10:10041742  T  G  2.966180 34090 0.003015329   rs2804081  10  10083705
## 3:  10:10045068  A  G  2.821558 34090 0.004779125   rs1540956  10  10087031
## 4: 10:100590740  C  G  3.179501 34090 0.001475310   rs4917903  10 102350497
## 5:  10:10109165  G  A  2.980699 34090 0.002875908  rs77473009  10  10151128
## 6: 10:101097541  A  G -2.824193 34090 0.004739842 rs117081694  10 102857298
```

Due to the large number of pGenes, we retained only the variants that met the criterion of P<0.05. Similarly, the outcome GWAS is:

```
LDL=read_parquet("LDLMR.parquet")
```

```
## Warning: Potentially unsafe or invalid elements have been discarded from R metadata.
## i Type: "externalptr"
## > If you trust the source, you can set ‘options(arrow.unsafe_metadata = TRUE)‘ to preserve them.
```

```
head(LDL)
```

```
##                      SNP CHR        BP  A1    A2   Freq     Zscore       N
## 1: 10:104850621_TAATA_T  10 104850621   T TAATA 0.0483 2.00815820 1093568
## 2: 10:106935726_TAGAG_T  10 106935726   T TAGAG 0.0998 1.33092322 1093566
## 3:   10:107002336_TTA_T  10 107002336 TTA     T 0.1030 1.35251330  935935
## 4:    10:107786863_TG_T  10 107786863   T    TG 0.0481 0.09892816 1092471
## 5:    10:107794764_AG_A  10 107794764   A    AG 0.0475 0.25930177 1063453
## 6: 10:107802797_TAAAC_T  10 107802797   T TAAAC 0.0497 0.95689729  487567
##            BETA           SE
## 1: 1.920326e-03 0.0009562625
## 2: 1.272713e-03 0.0009562633
## 3: 1.398037e-03 0.0010336587
## 4: 9.464877e-05 0.0009567424
## 5: 2.514469e-04 0.0009697077
## 6: 1.370403e-03 0.0014321313
```

Our next step is to use the `filter_align()` function from the MRBEE package for allele harmonization:

```
library(MRBEE)
```

```
##
## Attaching package: 'MRBEE'

## The following object is masked from 'package:TGVIS':
##
##     filter_align
```

```
GWAS=MRBEE::filter_align(gwas_data_list=list(LDL=LDL[,c("SNP","A1","A2","Zscore","N")],
                                             PCSK9=PCSK9[,c("SNP","A1","A2","Zscore","N")]),
                         ref_panel=PCSK9[,c("SNP","A1","A2")],allele_match=T)
```

```
## Adjusting effect allele according to reference panel...
## Finding common SNPs...
## Aligning data to common SNPs and ordering...
## Filtering complete.
```

```
LDL=GWAS$LDL
PCSK9=GWAS$PCSK
print(dim(LDL))
```

```
## [1] 74669     7
```

```
print(dim(PCSK9))
```

```
## [1] 74669     7
```

Our next step is to use the insignificant GWAS variants to estimate the correlation matrix of the estimation errors:

```
ind=which(abs(LDL$Zscore)<0.5)
Rxy=cor(cbind(PCSK9$Zscore[ind],LDL$Zscore[ind]))
Rxy
```

```
##           [,1]      [,2]
## [1,] 1.0000000 0.1089105
## [2,] 0.1089105 1.0000000
```

The next step is to select IVs using the C+T clumping method. Our parameters are set as `--clump-kb 1000 --clump-p1 5e-8 --clump-p2 5e-8 --clump-r2 0.01`:

```
plink=readRDS("LDLPCSK9IV.rds")
length(plink$SNP)
```

```
## [1] 24
```

We organize the data:

```
LDL=LDL[which(LDL$SNP%in%plink$SNP),]
PCSK9=PCSK9[which(PCSK9$SNP%in%plink$SNP),]
LDL$BETA=LDL$Zscore/sqrt(LDL$N);LDL$SE=1/sqrt(LDL$N)
PCSK9$BETA=PCSK9$Zscore/sqrt(PCSK9$N);PCSK9$SE=1/sqrt(PCSK9$N)
```

We perform univariable MR:

```
library(MRBEE)
library(MR.CUE)
library(IMRP)
library(MendelianRandomization)
fitMRBEE=MRBEE.IMRP.UV(by=LDL$BETA,bx=PCSK9$BETA,byse=LDL$SE,bxse=PCSK9$SE,Rxy=Rxy,
                        var.est="variance")

fitMRCUE=MRCUEIndep(Gammah=LDL$BETA,gammah=PCSK9$BETA,se2=LDL$SE,se1=PCSK9$SE,rho=Rxy[1,2])

INPUT=mr_input(by=LDL$BETA,bx=PCSK9$BETA,byse=LDL$SE,bxse=PCSK9$SE)

fitMED=mr_median(INPUT,iterations=100)

fitCML=mr_cML(INPUT,num_pert=100,MA=F,n=mean(PCSK9$N))

fitIMRP=MR_pleio(BetaOutcome="by",BetaExposure="bx",SdOutcome="byse",SdExposure="bxse",
                 data=data.frame(by=LDL$BETA,bx=PCSK9$BETA,byse=LDL$SE,bxse=PCSK9$SE),
                 SignifThreshold=0.05/length(PCSK9$SNP),rho=Rxy[1,2])
```

Finally, we summarize the results:

```
A=data.frame(Estimate=c(fitMED@Estimate,fitIMRP$CausalEstimate,fitMRCUE$beta.hat,
                        fitCML@Estimate,fitMRBEE$theta),
             SE=c(fitMED@StdError,fitIMRP$SdCausalEstimate,fitMRCUE$beta.se,
                  fitCML@StdError,sqrt(fitMRBEE$vartheta)))
A$P=pchisq((A$Estimate/A$SE)^2,1,lower.tail=F)
A$Method=c("MRMedian","IMRP","MRCUE","MRCML","MRBEE")
A$IV=paste(plink$SNP,collapse=",")
A$Rxy=Rxy[1,2]
A=as_tibble(A)
print(A)
```

```
## # A tibble: 5 x 6
##   Estimate      SE         P Method   IV                                   Rxy
##      <dbl>   <dbl>     <dbl> <chr>    <chr>                              <dbl>
## 1    0.320 0.00844 4.24e-315 MRMedian rs11591147,rs472495,rs174568,rs1260~ 0.109
## 2    0.310 0.0137  1.57e-112 IMRP     rs11591147,rs472495,rs174568,rs1260~ 0.109
## 3    0.380 0.135   4.89e-  3 MRCUE    rs11591147,rs472495,rs174568,rs1260~ 0.109
## 4    0.143 0.0184  6.87e- 15 MRCML    rs11591147,rs472495,rs174568,rs1260~ 0.109
## 5    0.337 0.0237  8.38e- 46 MRBEE    rs11591147,rs472495,rs174568,rs1260~ 0.109
```

### 5 Supplemental Simulation Results

#### 5.1 Pratt index estimation

Here, we present additional simulations. Our primary focus is on whether the Pratt index can accurately estimate the contributions of gene-tissue pairs, direct causal variants, and infinitesimal effects. To better illustrate the properties of the Pratt index, we consider three key measures:

- Practical Estimation: The Pratt index estimate obtained from the actual data.
- Oracle Estimation: Variable selection often introduces shrinkage bias, especially when small effects are compressed to zero by SuSiE, making it difficult to accurately evaluate the Pratt index. To address this, we consider directly using the true non-zero gene-tissue pairs $\mathbf{B}_{\mathcal{M}_\theta}$ and the true direct causal variants $\mathbf{R}_{\mathcal{M}_\gamma}$ (in cases with no direct causal variant, two variants are randomly selected to ensure smooth program execution).
- True Values of Pratt Indices: The actual Pratt index values for each component.

We begin by fixing the local heritability of the outcome at $h_y^2 = 5 \times 10^{-3}$, a value achievable for the first few loci on each chromosome of the cardiovascular and metabolic traits we are studying. We will then demonstrate, step by step, how the three components and total Pratt index estimations behave under different numbers of eQTLs in this setting.

It can be observed in Figure S1 that Pratt index estimates tend to underestimate the true values, and the degree of underestimation is largely unaffected by the eQTL sample size. This is due to the estimation error present in the outcome GWAS marginal effect estimates, which increases the uncertainty in the model. This uncertainty is only reduced by (1) an increase in the local heritability of the outcome, or (2) an increase in the GWAS sample size. Both factors would increase the Z-scores of the outcome GWAS marginal effect sizes, while the estimation error of the Z-score remains fixed at 1. This issue has been discussed in various studies, such as those on LDSC (Bulik-Sullivan et al., 2015) and MRBEE (Lorincz-Comi et al., 2024).

Figures S2-S4 show the Pratt index estimations when the number of eQTL is 2, 3, and 4, respectively. A notable difference arises when the eQTL sample size is small: due to the low power in such cases, SuSiE tends to overlook smaller-scale eQTL effects, causing these overlooked eQTL to be mischaracterized as direct causal variants or described within the infinitesimal effect. Since the total heritability of each gene-tissue pair is fixed at $h_1^2 = \cdots = h_{100}^2 = 0.3$, the average effect of each eQTL decreases as the number of eQTLs increases. Consequently, in practice, some smaller effects may be incorrectly compressed to zero. This introduces a third source factor, in addition to the local heritability of the outcome and the GWAS sample size, that contributes to the underestimation of the Pratt index from gene-tissue pairs.

To validate our hypothesis that increasing the local heritability of the outcome can mitigate the underestimation of the Pratt index, we fixed the eQTL sample size at 200 and increased the local heritability of the outcome from $1 \times 10^{-3}$ to $5 \times 10^{-3}$. We did not consider increasing the sample size of the outcome, as this is equivalent to increasing the local heritability: increasing the local heritability by a factor of $k$ is mathematically equivalent to increasing the sample size by a factor of $k$. Figures S5-S8 clearly demonstrate that increasing the local heritability of the outcome significantly alleviates the underestimation of the Pratt index.

In conclusion, we find that the Pratt index is not an unbiased estimator; it tends to underestimate the true Pratt index. Generally, this underestimation is due to estimation errors in the marginal effect sizes of the outcome GWAS, leading to measurement error bias (Yi, 2016; Lorincz-Comi et al., 2024). Enhancing the reliability score (Yi, 2016) can reduce this measurement error bias, which can be achieved by (1) increasing the local heritability of the outcome or, equivalently, (2) increasing the GWAS sample size for the outcome. Fortunately, compared to overestimating the Pratt index, underestimating it results in more conservative inferences, potentially leading to the omission of some causal gene-tissue pairs with CS-Pratt $< 0.15$. In the future, with larger sample sizes for the outcome GWAS, we can revisit these cases for further analysis.

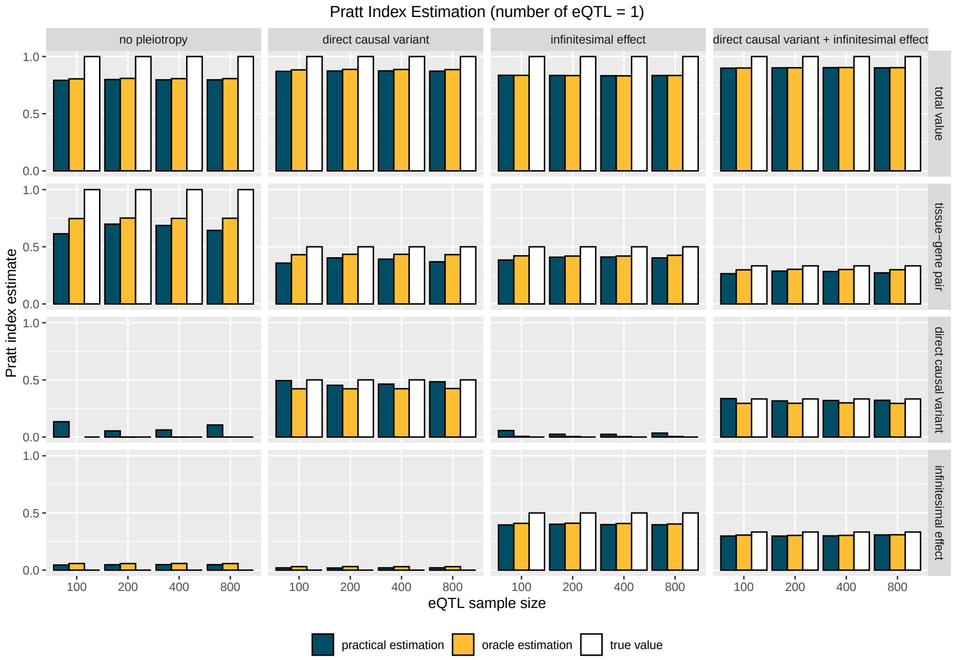

Figure S1: This figure shows how the practical estimation, oracle estimation, and true values of Pratt indices for gene-tissue pairs, direct causal variants, and infinitesimal effects change as **the eQTL sample size** increases, with **the number of eQTL to 1**.

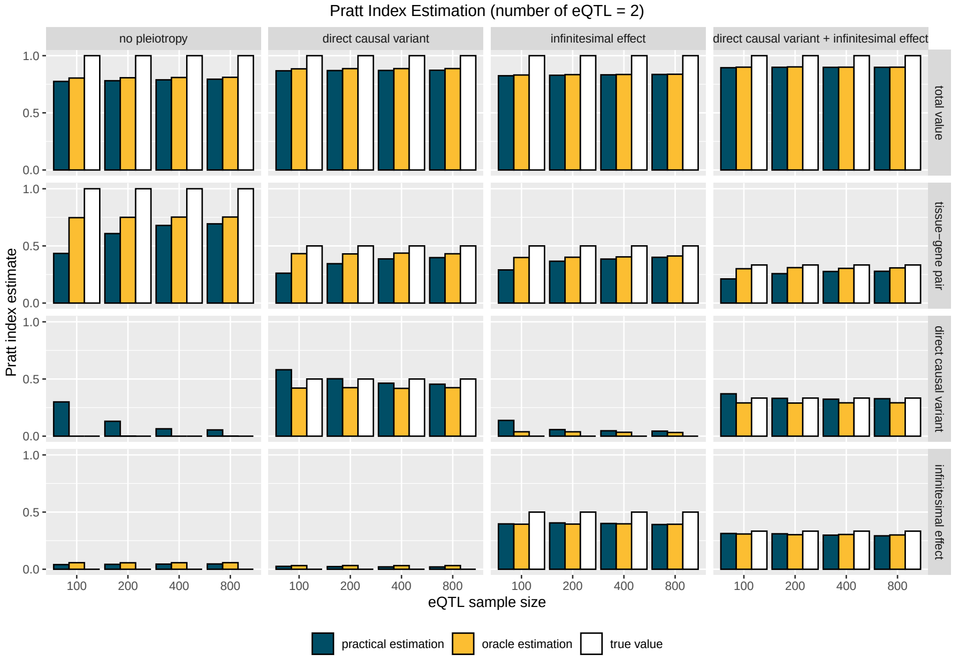

Figure S2: This figure shows how the practical estimation, oracle estimation, and true values of Pratt indices for gene-tissue pairs, direct causal variants, and infinitesimal effects change as **the eQTL sample size** increases, with **the number of eQTL to 2**.

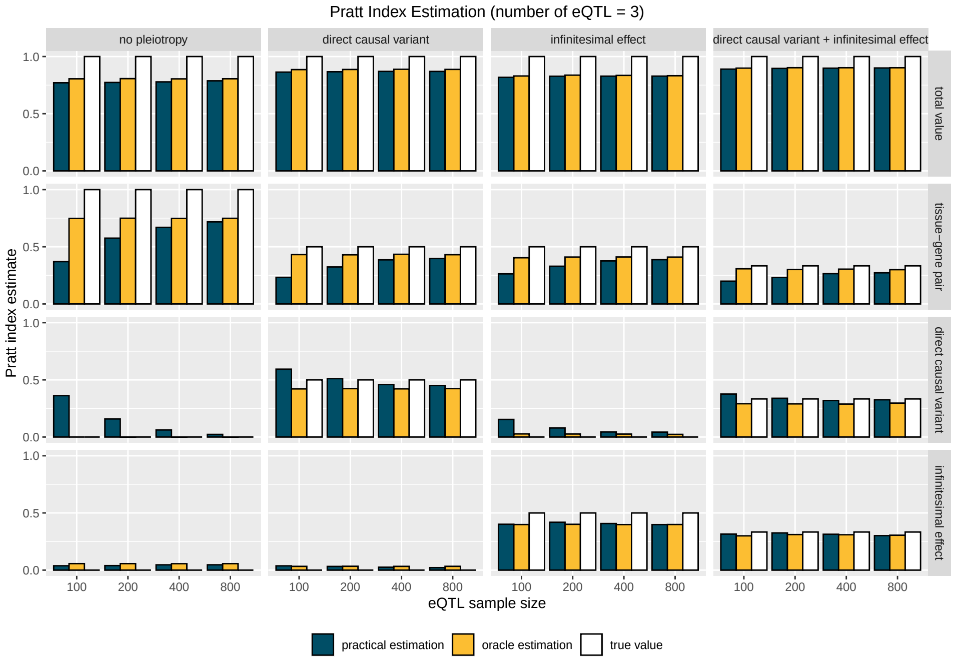

Figure S3: This figure shows how the practical estimation, oracle estimation, and true values of Pratt indices for gene-tissue pairs, direct causal variants, and infinitesimal effects change as **the eQTL sample size** increases, with **the number of eQTL to 3**.

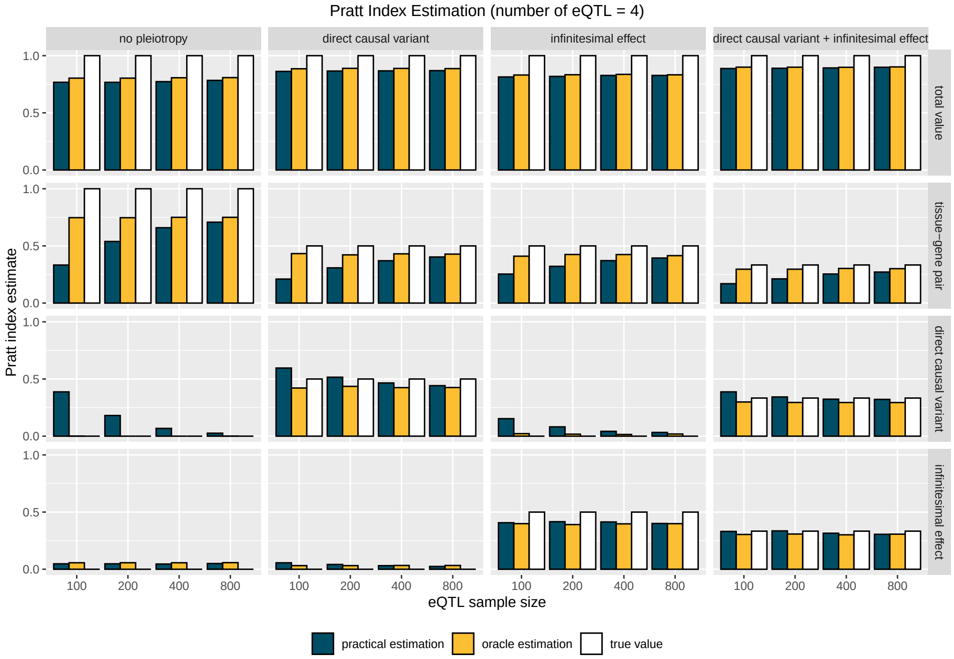

Figure S4: This figure shows how the practical estimation, oracle estimation, and true values of Pratt indices for gene-tissue pairs, direct causal variants, and infinitesimal effects change as **the eQTL sample size** increases, with **the number of eQTL to 4**.

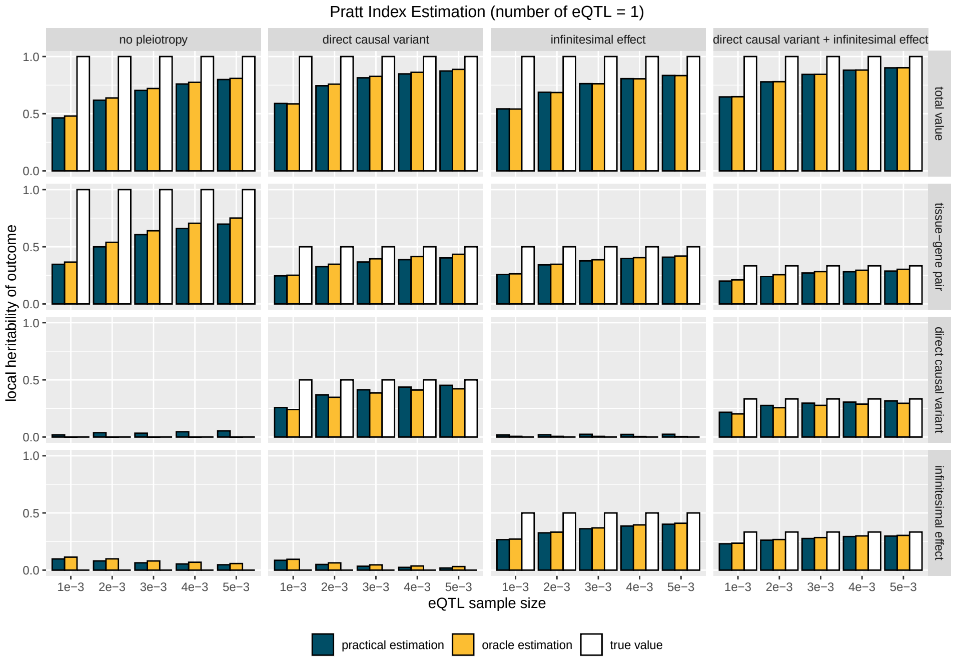

Figure S5: This figure shows how the practical estimation, oracle estimation, and true values of Pratt indices for gene-tissue pairs, direct causal variants, and infinitesimal effects change as **local heritability of the outcome** increases, with **the number of eQTL to 1**.

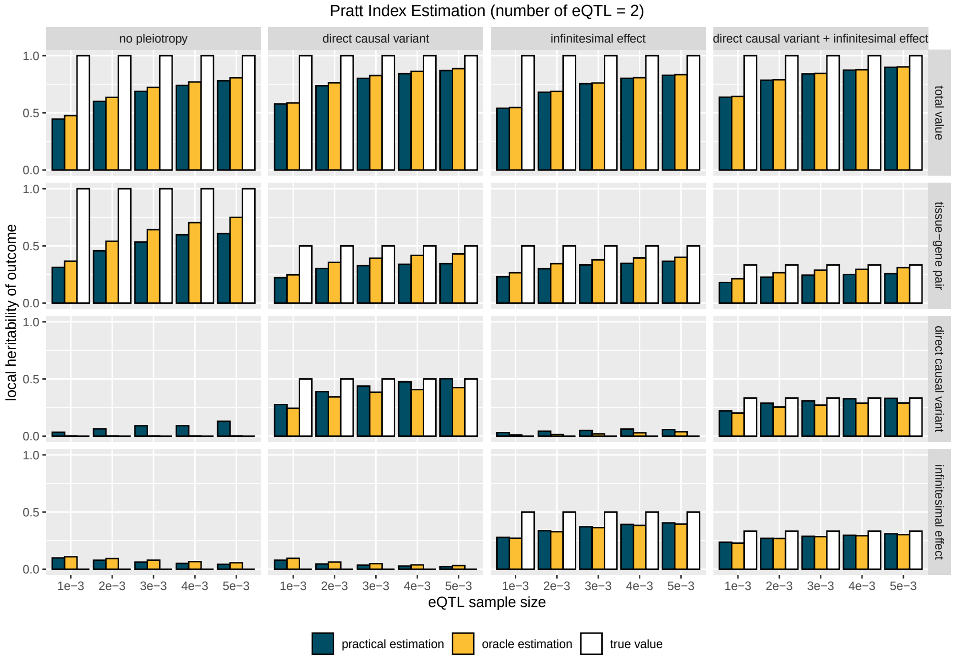

Figure S6: This figure shows how the practical estimation, oracle estimation, and true values of Pratt indices for gene-tissue pairs, direct causal variants, and infinitesimal effects change as **local heritability of the outcome** increases, with **the number of eQTL to 2**.

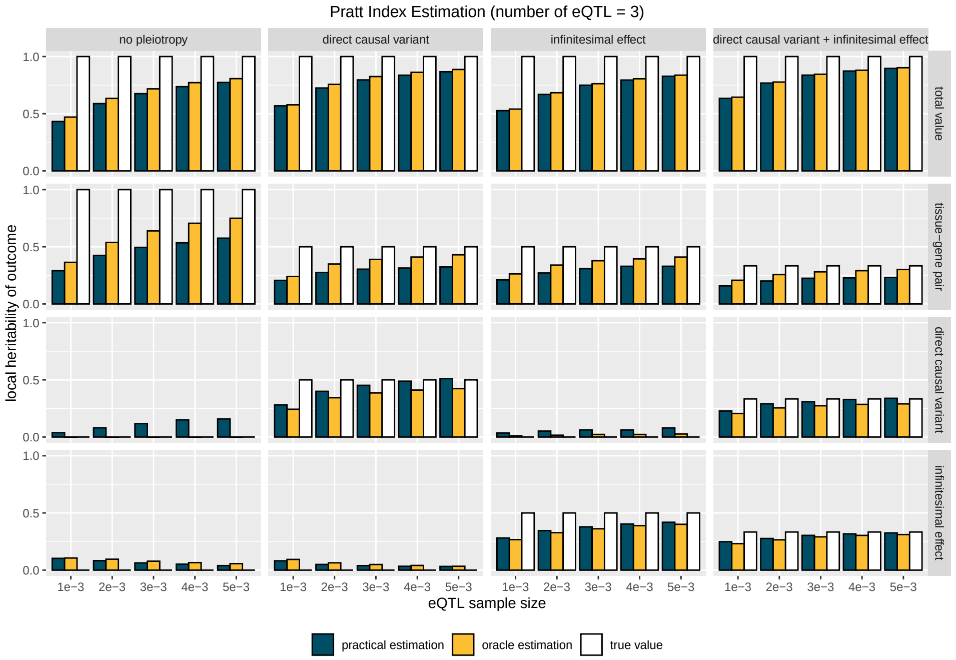

Figure S7: This figure shows how the practical estimation, oracle estimation, and true values of Pratt indices for gene-tissue pairs, direct causal variants, and infinitesimal effects change as **local heritability of the outcome** increases, with **the number of eQTL to 3**.

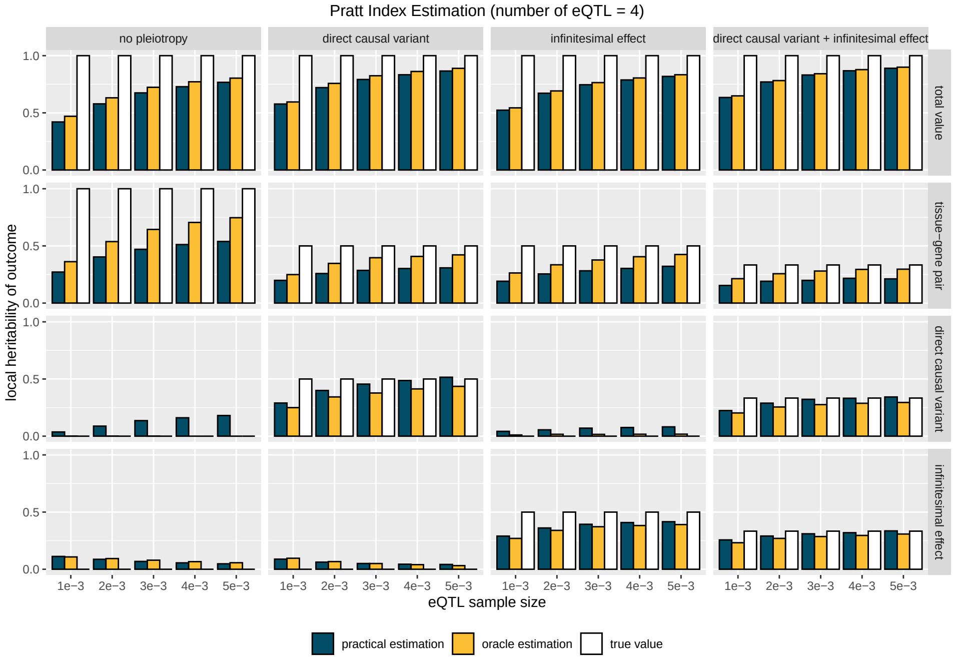

Figure S8: This figure shows how the practical estimation, oracle estimation, and true values of Pratt indices for gene-tissue pairs, direct causal variants, and infinitesimal effects change as **local heritability of the outcome** increases, with **the number of eQTL to 4**.

#### 5.2 Main simulation in other scenarios

Next, we present the results for additional scenarios related to Figure 2 from the main text, as shown in Figures S9-S15 below:

#### 5.3 Comparison of eQTL selecton

There are also some differences between TGVIS and TGFM during the eQTL selection stage. Specifically, according to the description of TGFM, it considers all variants in the 95% credible set as causal variants and applies parameter bootstrap according to the posterior distribution provided by SuSiE. In contrast, TGVIS directly selects eQTLs within the 95% credible set using the effect sizes provided by SuSiE. We have observed that the function `susie_get_cs()` in SuSiE sometimes identifies credible sets containing a large number of variants. To prevent this, we impose an additional requirement that each variant's individual PIP must exceed a certain threshold (e.g., 0.25), which avoids situations where more than four eQTLs in a credible set share the PIP equally. Since we have already used the C+T approach to remove highly correlated variants before performing eQTL selection, ideally, each credible set should contain only one variant. We therefore believe that our procedure of eQTL selection should be more robust.

We conducted a simple simulation to evaluate the two procedures of eQTL selection. We found that, in most cases, both methods are accurate. However, when the eQTL sample size is smaller, the TGFM approach tends to produce more outliers (Figure S16-S17). Therefore, we comment in the paper that under conditions of smaller eQTL sample sizes, the TGFM approach introduces more uncertainty, which can actually reduce the precision of fine-mapping gene-tissue pairs in multivariable TWAS. The R code is as follows:

```
tgfm.resampling=function(alpha,mu,mu2,sampling=500){
L=dim(mu)[1]
n=dim(mu)[2]
a=c(mu[1,]*0)
M=matrix(0,sampling,n)
for(j in 1:L){
phi=t(rmultinom(sampling,1,alpha[j,]))
varr=c(mu2[j,])-c(mu[j,])^2
varr[varr<0]=0
N=MASS::mvrnorm(n=sampling,mu=mu[j,],Sigma=diag(varr))
N=N*phi
M=M+N
}
return(M)
}

susie_get_cs_index=function(res,coverge=0.95,min_abs_corr=0.5){
A=susie_get_cs(res,coverage=coverge,min_abs_corr=min_abs_corr)
if(is.null(A)!=1){
A=A$cs
s=c()
for(i in 1:length(A)){
s=c(s,A[[i]])
}
}else{
s=c()
}
return(s)
}
```

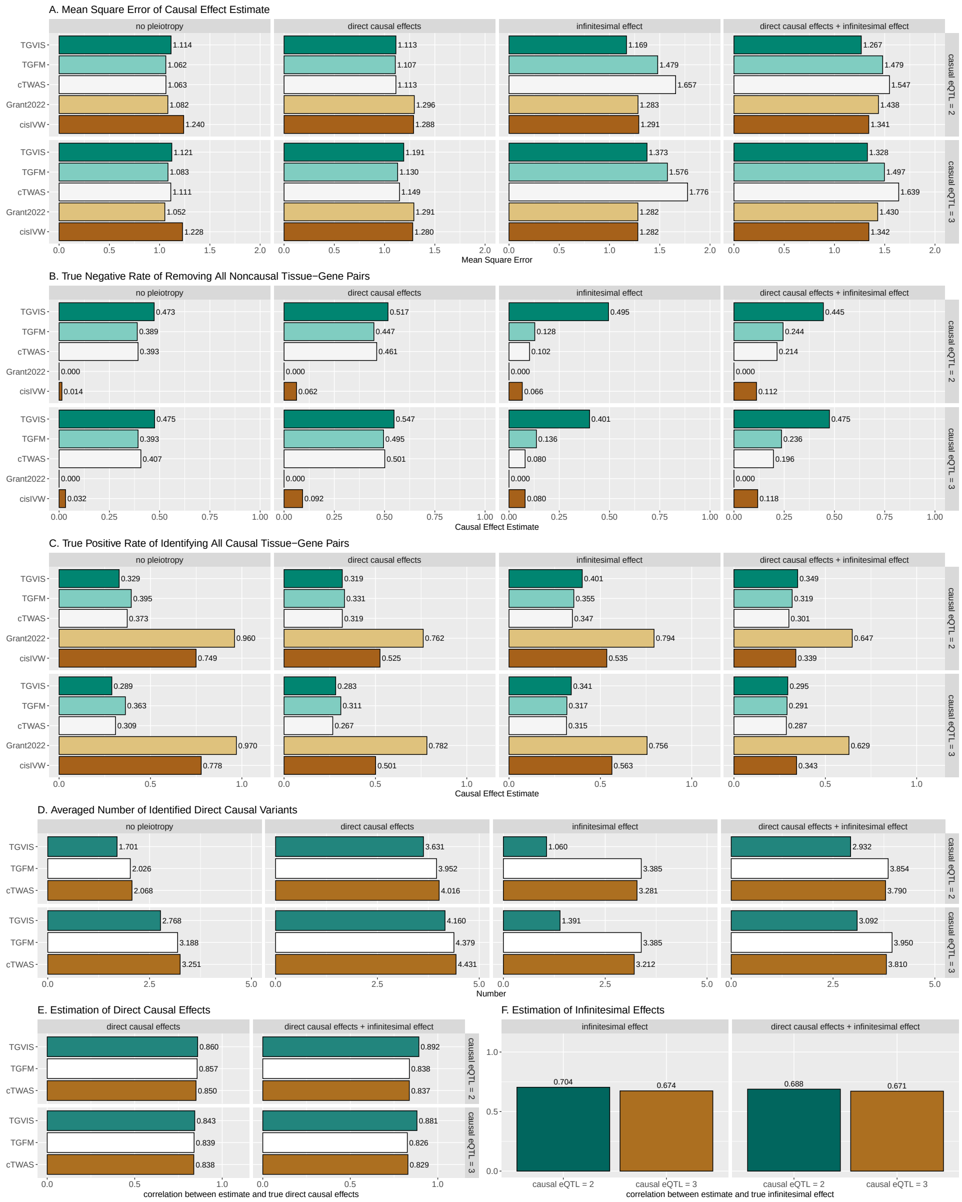

Figure S9: Simulation results comparing the performance of TGVIS, TGFM, cTWAS, Grant2022, and cisIVW in a scenario with eQTL sample size = 100, number of eQTL = 2 and 3. A: The MSE of causal effect estimates under no pleiotropy and in the presence of direct causal variants, infinitesimal effects, and both. B: The true negative rates of removing all the 98 noncausal gene-tissue pairs under different scenarios. This means that if a method incorrectly identifies any non-causal pairs as causal, the true negative events (correctly identified non-causal pairs) will not be counted or considered. C: Bar plots display the true positive rates of identifying all 2 causal gene-tissue pairs under different scenarios. D: The averaged number of identified direct causal variants. The true numbers of no-pleiotropy, direct-causal-variant, infinitesimal-effects, and direct-causal-variant and infinitesimal-effects cases are 0, 2, 0, 2. E: The empirical correlation of the true direct causal effect vector and its estimate cor($\hat{\boldsymbol{\gamma}}$, $\boldsymbol{\gamma}$). F: The empirical correlation of the true infinitesimal effect vector and its estimate cor($\hat{\boldsymbol{\upsilon}}$, $\boldsymbol{\upsilon}$).

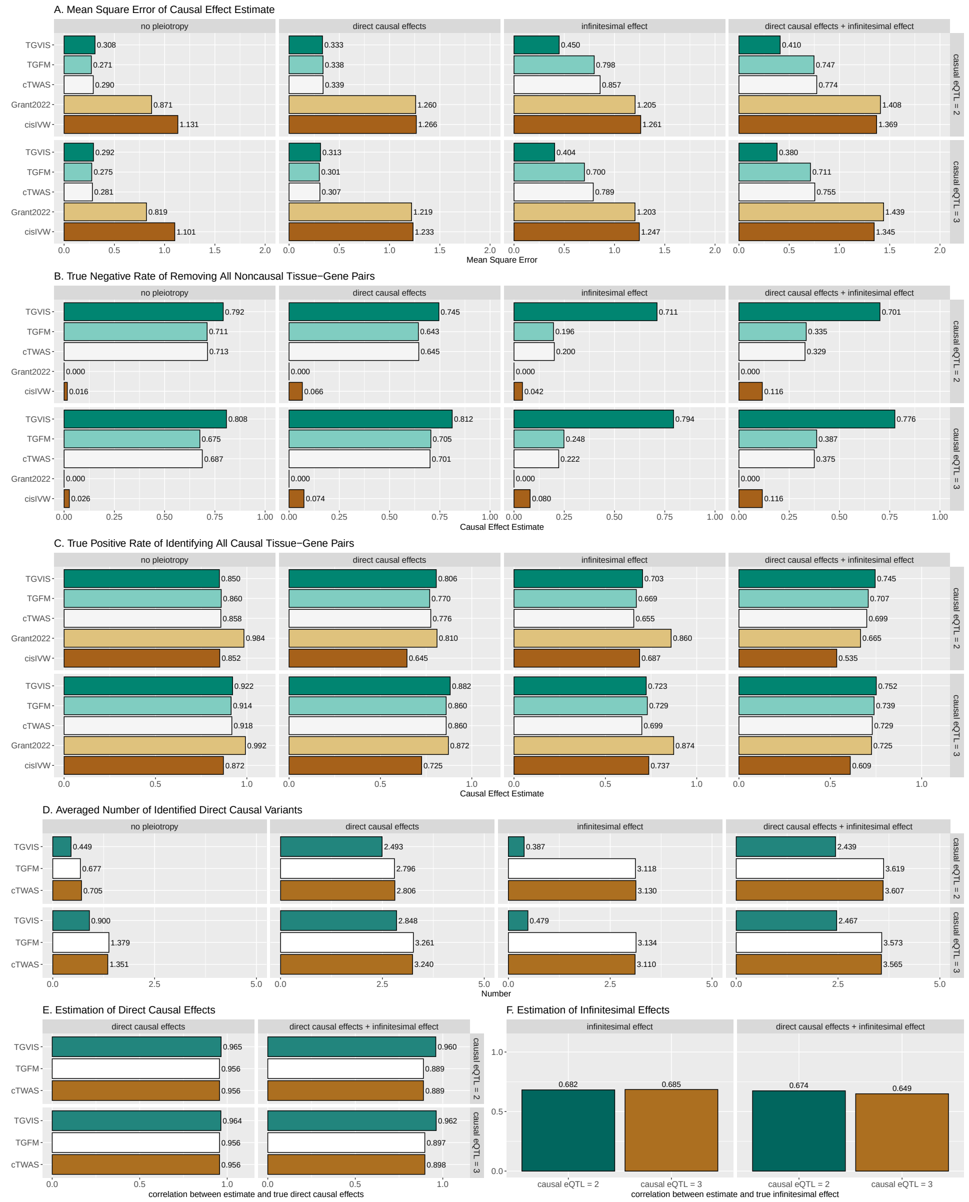

Figure S10: Simulation results comparing the performance of TGVIS, TGFM, cTWAS, Grant2022, and cisIVW in a scenario with eQTL sample size = 400, number of eQTL = 2 and 3. A: The MSE of causal effect estimates under no pleiotropy and in the presence of direct causal variants, infinitesimal effects, and both. B: The true negative rates of removing all the 98 noncausal gene-tissue pairs under different scenarios. This means that if a method incorrectly identifies any non-causal pairs as causal, the true negative events (correctly identified non-causal pairs) will not be counted or considered. C: Bar plots display the true positive rates of identifying all 2 causal gene-tissue pairs under different scenarios. D: The averaged number of identified direct causal variants. The true numbers of no-pleiotropy, direct-causal-variant, infinitesimal-effects, and direct-causal-variant and infinitesimal-effects cases are 0, 2, 0, 2. E: The empirical correlation of the true direct causal effect vector and its estimate cor($\hat{\boldsymbol{\gamma}}$, $\boldsymbol{\gamma}$). F: The empirical correlation of the true infinitesimal effect vector and its estimate cor($\hat{\boldsymbol{\upsilon}}$, $\boldsymbol{\upsilon}$).

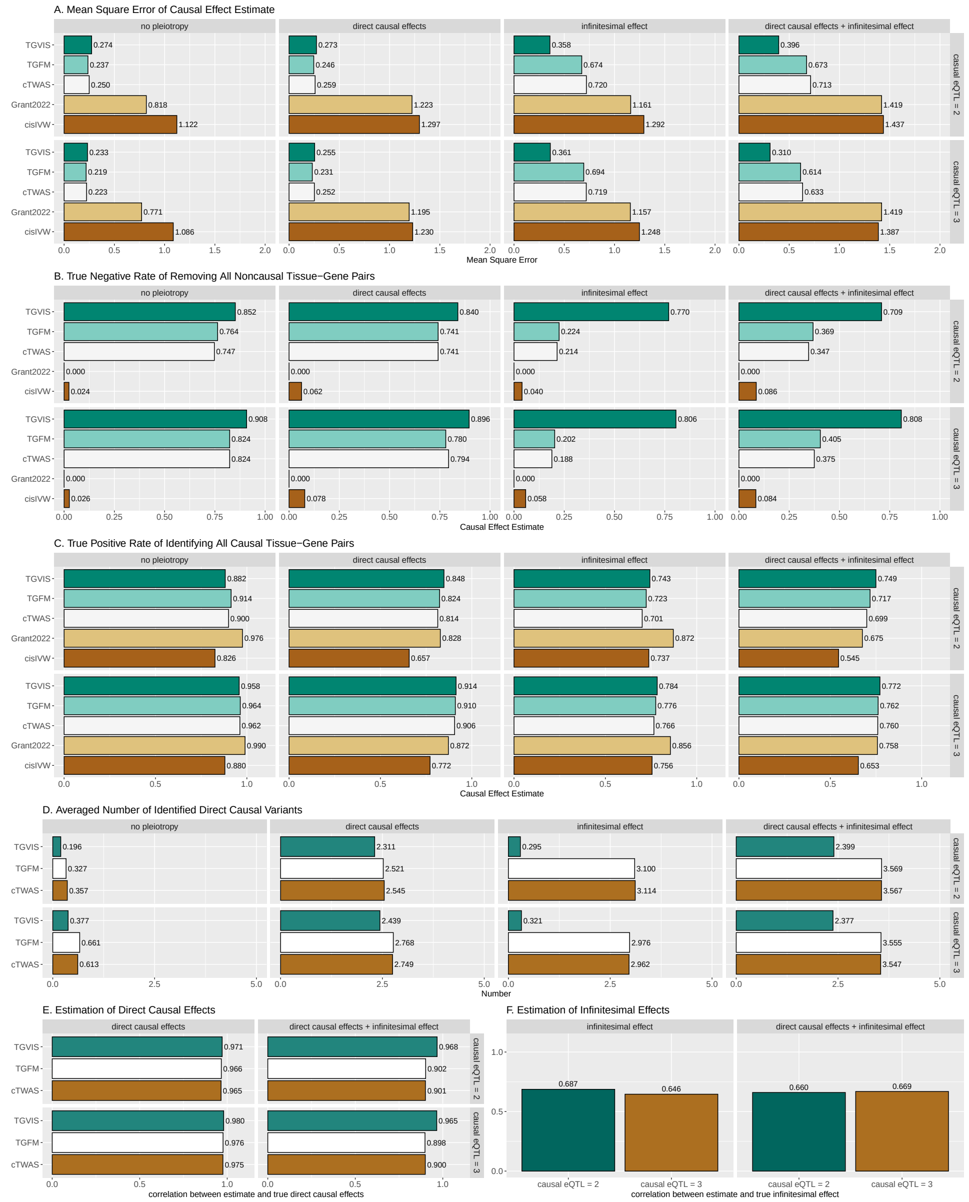

Figure S11: Simulation results comparing the performance of TGVIS, TGFM, cTWAS, Grant2022, and cisIVW in a scenario with eQTL sample size = 800, number of eQTL = 2 and 3. A: The MSE of causal effect estimates under no pleiotropy and in the presence of direct causal variants, infinitesimal effects, and both. B: The true negative rates of removing all the 98 noncausal gene-tissue pairs under different scenarios. This means that if a method incorrectly identifies any non-causal pairs as causal, the true negative events (correctly identified non-causal pairs) will not be counted or considered. C: Bar plots display the true positive rates of identifying all 2 causal gene-tissue pairs under different scenarios. D: The averaged number of identified direct causal variants. The true numbers of no-pleiotropy, direct-causal-variant, infinitesimal-effects, and direct-causal-variant and infinitesimal-effects cases are 0, 2, 0, 2. E: The empirical correlation of the true direct causal effect vector and its estimate cor($\hat{\boldsymbol{\gamma}}$, $\boldsymbol{\gamma}$). F: The empirical correlation of the true infinitesimal effect vector and its estimate cor($\hat{\boldsymbol{v}}$, $\boldsymbol{v}$).

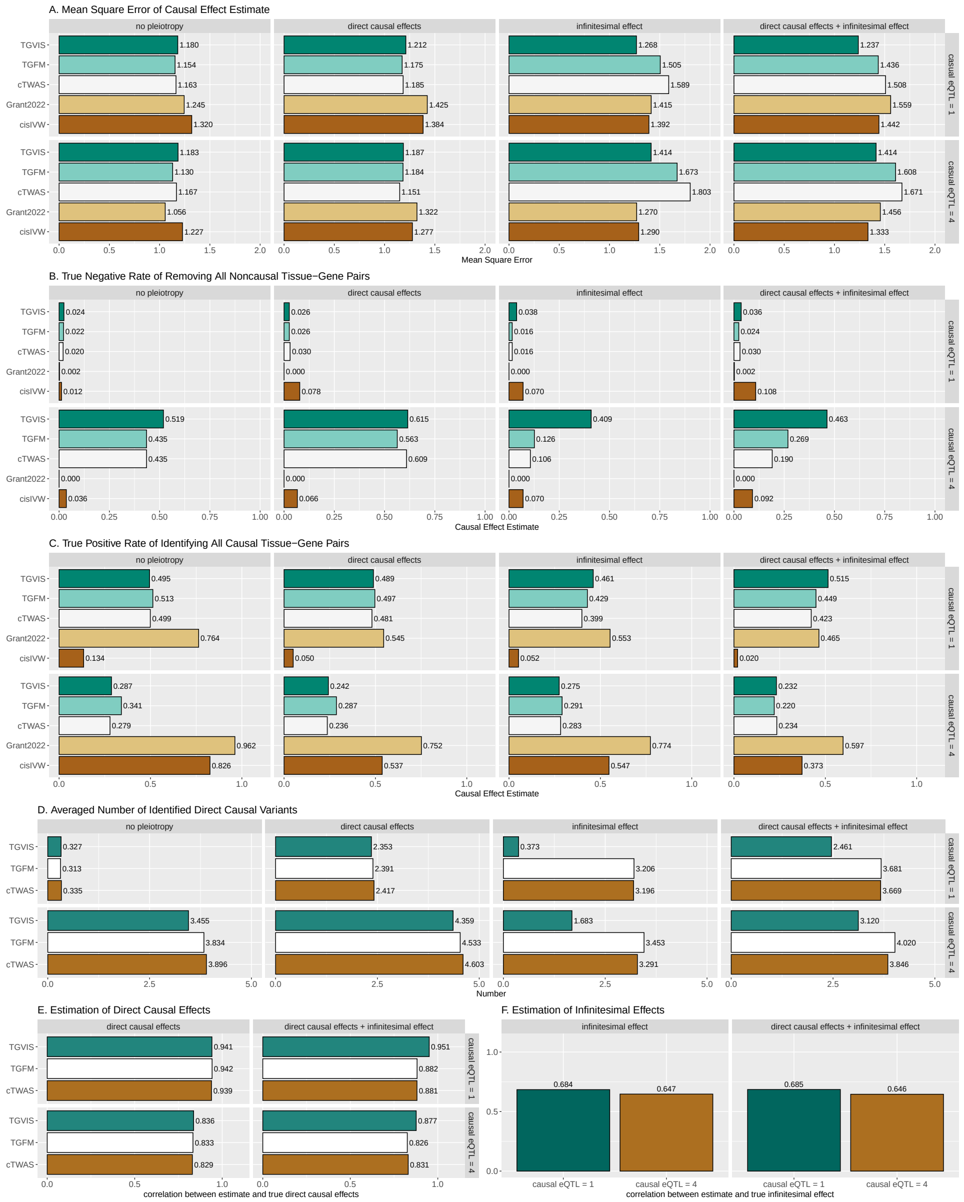

Figure S12: Simulation results comparing the performance of TGVIS, TGFM, cTWAS, Grant2022, and cisIVW in a scenario with eQTL sample size = 100, number of eQTL = 1 and 4. A: The MSE of causal effect estimates under no pleiotropy and in the presence of direct causal variants, infinitesimal effects, and both. B: The true negative rates of removing all the 98 noncausal gene-tissue pairs under different scenarios. This means that if a method incorrectly identifies any non-causal pairs as causal, the true negative events (correctly identified non-causal pairs) will not be counted or considered. C: Bar plots display the true positive rates of identifying all 2 causal gene-tissue pairs under different scenarios. D: The averaged number of identified direct causal variants. The true numbers of no-pleiotropy, direct-causal-variant, infinitesimal-effects, and direct-causal-variant and infinitesimal-effects cases are 0, 2, 0, 2. E: The empirical correlation of the true direct causal effect vector and its estimate $\text{cor}(\hat{\boldsymbol{\gamma}}, \boldsymbol{\gamma})$. F: The empirical correlation of the true infinitesimal effect vector and its estimate $\text{cor}(\hat{\boldsymbol{v}}, \boldsymbol{v})$.

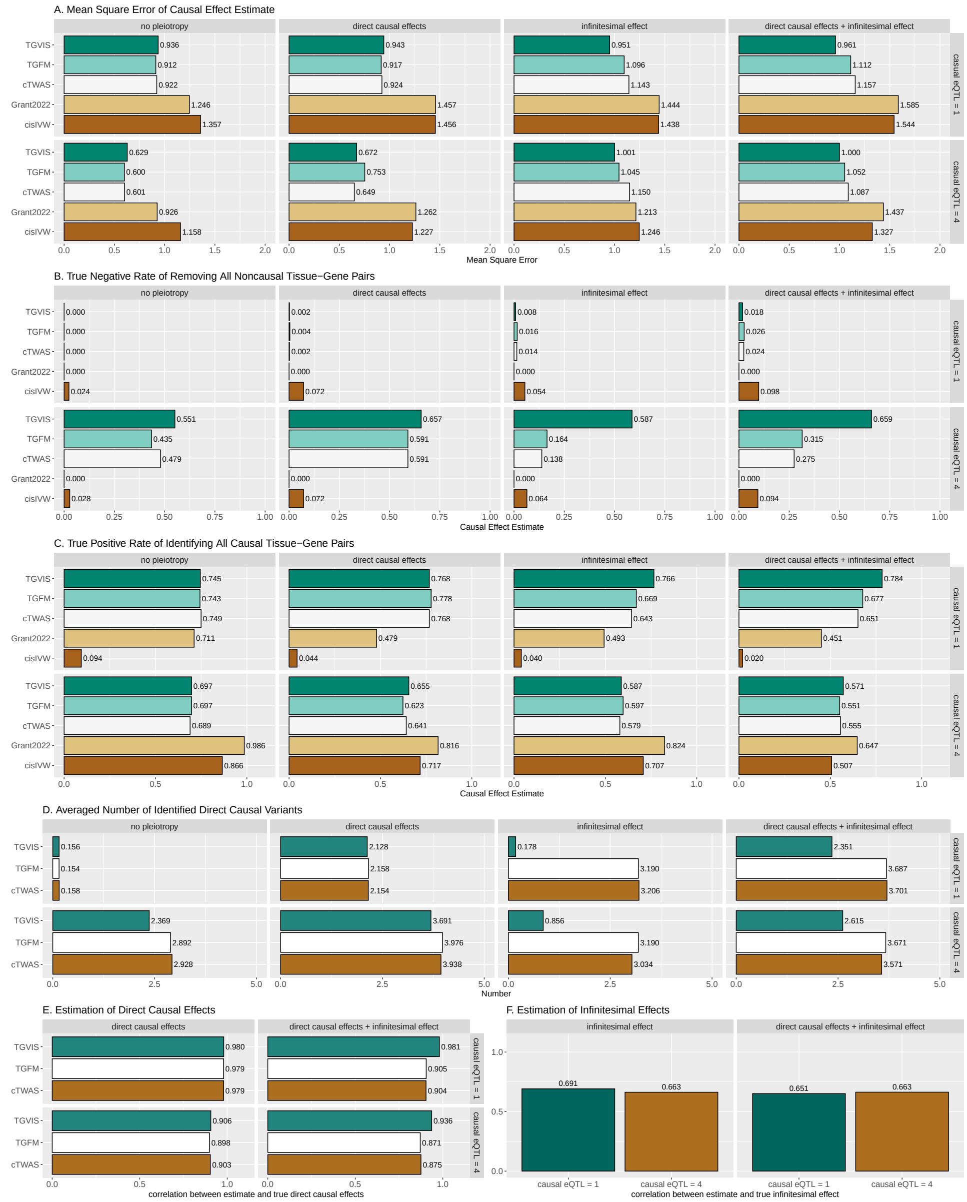

Figure S13: Simulation results comparing the performance of TGVIS, TGFM, cTWAS, Grant2022, and cisIVW in a scenario with eQTL sample size = 200, number of eQTL = 1 and 4. A: The MSE of causal effect estimates under no pleiotropy and in the presence of direct causal variants, infinitesimal effects, and both. B: The true negative rates of removing all the 98 noncausal gene-tissue pairs under different scenarios. This means that if a method incorrectly identifies any non-causal pairs as causal, the true negative events (correctly identified non-causal pairs) will not be counted or considered. C: Bar plots display the true positive rates of identifying all 2 causal gene-tissue pairs under different scenarios. D: The averaged number of identified direct causal variants. The true numbers of no-pleiotropy, direct-causal-variant, infinitesimal-effects, and direct-causal-variant and infinitesimal-effects cases are 0, 2, 0, 2. E: The empirical correlation of the true direct causal effect vector and its estimate $\text{cor}(\hat{\boldsymbol{\gamma}}, \boldsymbol{\gamma})$. F: The empirical correlation of the true infinitesimal effect vector and its estimate $\text{cor}(\hat{\boldsymbol{v}}, \boldsymbol{v})$.

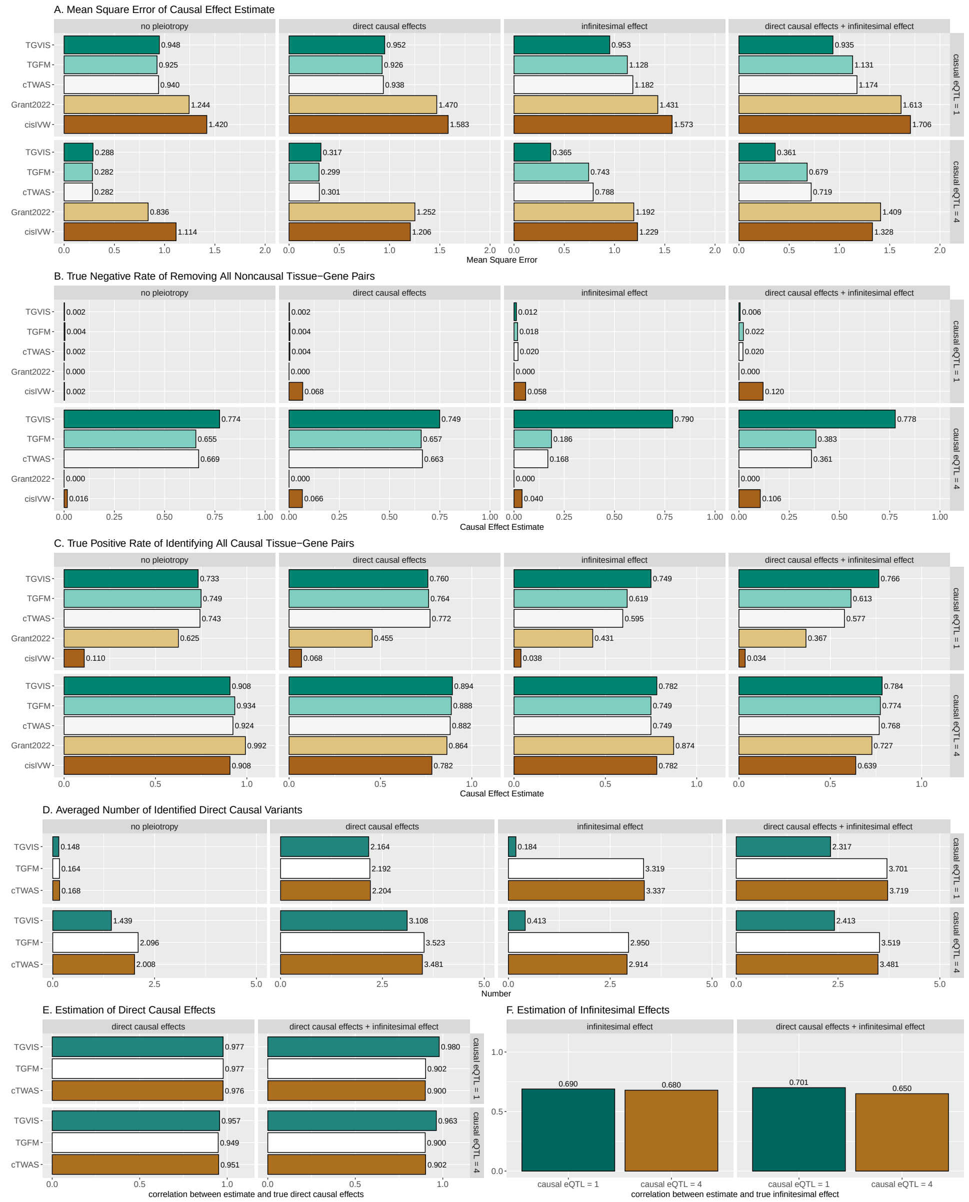

Figure S14: Simulation results comparing the performance of TGVIS, TGFM, cTWAS, Grant2022, and cisIVW in a scenario with eQTL sample size = 400, number of eQTL = 1 and 4. A: The MSE of causal effect estimates under no pleiotropy and in the presence of direct causal variants, infinitesimal effects, and both. B: The true negative rates of removing all the 98 noncausal gene-tissue pairs under different scenarios. This means that if a method incorrectly identifies any non-causal pairs as causal, the true negative events (correctly identified non-causal pairs) will not be counted or considered. C: Bar plots display the true positive rates of identifying all 2 causal gene-tissue pairs under different scenarios. D: The averaged number of identified direct causal variants. The true numbers of no-pleiotropy, direct-causal-variant, infinitesimal-effects, and direct-causal-variant and infinitesimal-effects cases are 0, 2, 0, 2. E: The empirical correlation of the true direct causal effect vector and its estimate cor($\hat{\boldsymbol{\gamma}}$, $\boldsymbol{\gamma}$). F: The empirical correlation of the true infinitesimal effect vector and its estimate cor($\hat{\boldsymbol{v}}$, $\boldsymbol{v}$).

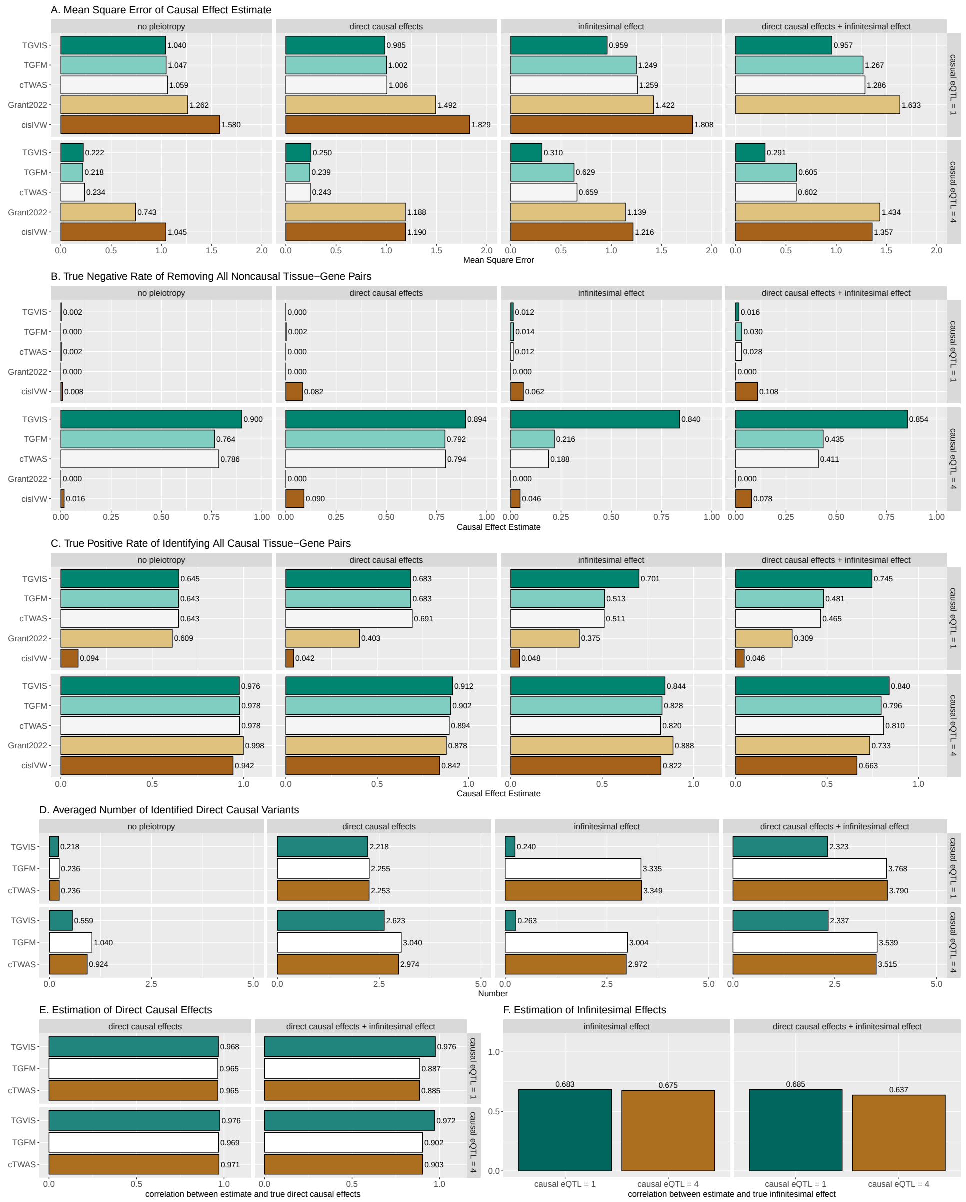

Figure S15: Simulation results comparing the performance of TGVIS, TGFM, cTWAS, Grant2022, and cisIVW in a scenario with eQTL sample size = 800, number of eQTL = 1 and 4. A: The MSE of causal effect estimates under no pleiotropy and in the presence of direct causal variants, infinitesimal effects, and both. B: The true negative rates of removing all the 98 noncausal gene-tissue pairs under different scenarios. This means that if a method incorrectly identifies any non-causal pairs as causal, the true negative events (correctly identified non-causal pairs) will not be counted or considered. C: Bar plots display the true positive rates of identifying all 2 causal gene-tissue pairs under different scenarios. D: The averaged number of identified direct causal variants. The true numbers of no-pleiotropy, direct-causal-variant, infinitesimal-effects, and direct-causal-variant and infinitesimal-effects cases are 0, 2, 0, 2. E: The empirical correlation of the true direct causal effect vector and its estimate $\mathrm{cor}(\hat{\boldsymbol{\gamma}}, \boldsymbol{\gamma})$. F: The empirical correlation of the true infinitesimal effect vector and its estimate $\mathrm{cor}(\hat{\boldsymbol{v}}, \boldsymbol{v})$.

```
LDlist=readRDS("~/MRJones_CRE/LD.rds")
LDsqrtlist=readRDS("~/MRJones_CRE/LDsqrt.rds")
blockdim=unlist(lapply(LDlist,nrow))
blockindexend=cumsum(blockdim)
blockindexstart=blockindexend-blockdim+1

par(mfrow=c(3,3))
N=c(50,100,200)
NeQTL=c(1:3)
for(n in N){
for(neQTL in NeQTL){
h2=0.1
MSE=matrix(0,200,2)
for(iter in 1:200){
Blist=Alist=list()
i=sample(10,1,replace=F)
G=LDlist[[i]]
G=G*0.67+diag(diag(G))*0.33
b=0*G[,1]
b[sample(nrow(G),neQTL)]=rnorm(neQTL,0,1)
b=b/sqrt(sum(b^2))*h2
hatb=c(G%*%b)+MASS::mvrnorm(n=1,mu=rep(0,nrow(G)),Sigma=G)/sqrt(n)
z=hatb*sqrt(n)
fit=susieR::susie_rss(z=z,R=G,n=n,L=5)
## Get the 95% credible set for TGVIS resampling
beta1=coef(fit)[-1]
index.causal=intersect(unique(susie_get_cs_index(fit)),which(fit$pip>0.25))
if(length(index.causal)>0){
beta1[-index.causal]=0
}else{
beta1=beta1*(fit$pip>0.5)
}
## Get the 95% credible set for TGFM resampling
beta2=colMeans(tgfm.resampling(alpha=fit$alpha,mu=fit$mu,mu2=fit$mu2,sampling=100))
index.causal=intersect(unique(susie_get_cs_index(fit)),which(fit$pip>0.05))
if(length(index.causal)>0){
beta2[-index.causal]=0
}else{
beta2=beta2*0
}
MSE[iter,]=c(norm(b-beta1,"2"),norm(b-beta2,"2"))
}
colnames(MSE)=c("TGVIS","TGFM")
boxplot(MSE,main=glue("neQTL={n}, h2=0.1, number={neQTL}",ylab="2-norm of eQTL estimate"))
}
}
```

### 6 Supplemental Data Analysis Results

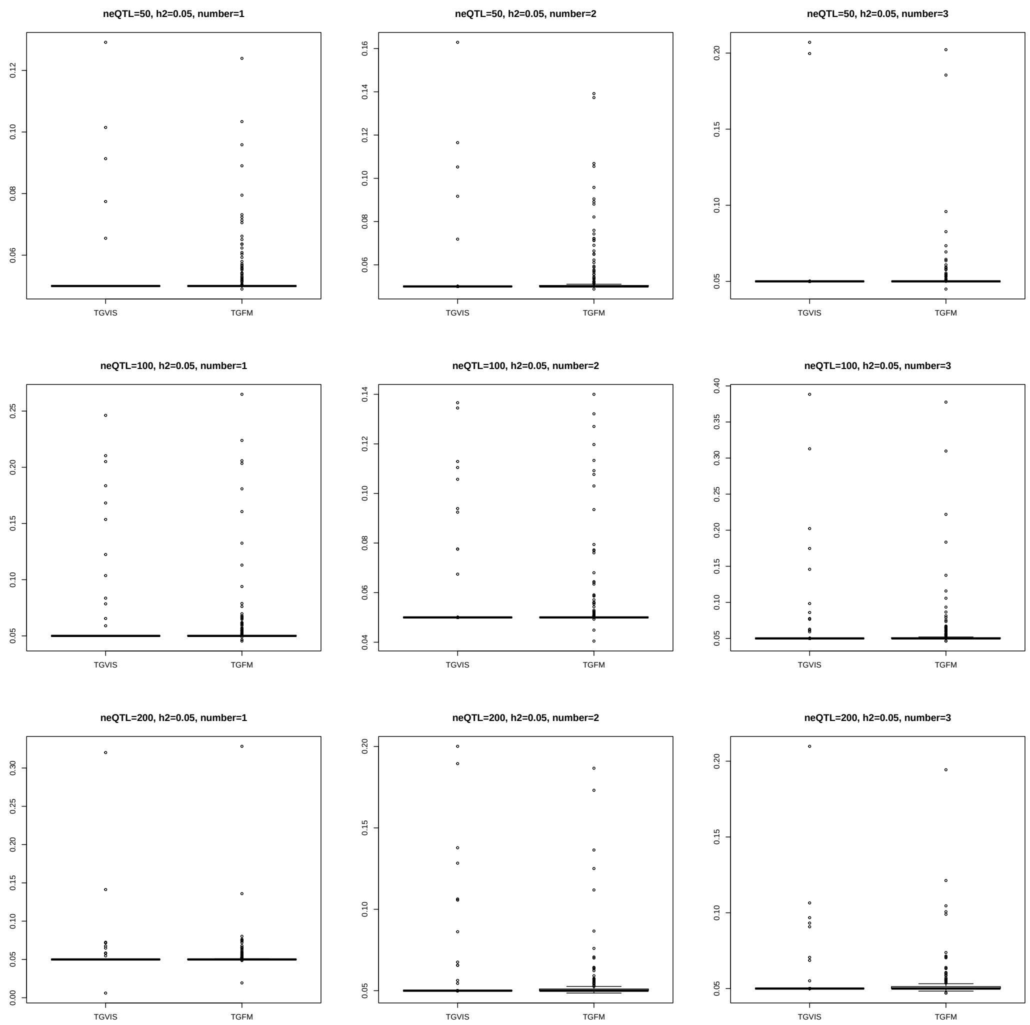

Figure S16: This figure shows the boxplot of the 2-norm of the difference between the eQTL effect estimates obtained by TGVIS and TGFM and the true values, i.e., $||\hat{\boldsymbol{\beta}}_{\text{TGVIS}} - \boldsymbol{\beta}||_2$ and $||\hat{\boldsymbol{\beta}}_{\text{TGFM}} - \boldsymbol{\beta}||_2$, across 100 simulations. The simulations are conducted with the heritability of gene-tissue pairs fixed at $h^2_{\text{tissue-gene}} = 0.05$, varying the number of eQTLs (number) and the eQTL sample size (neQTL).

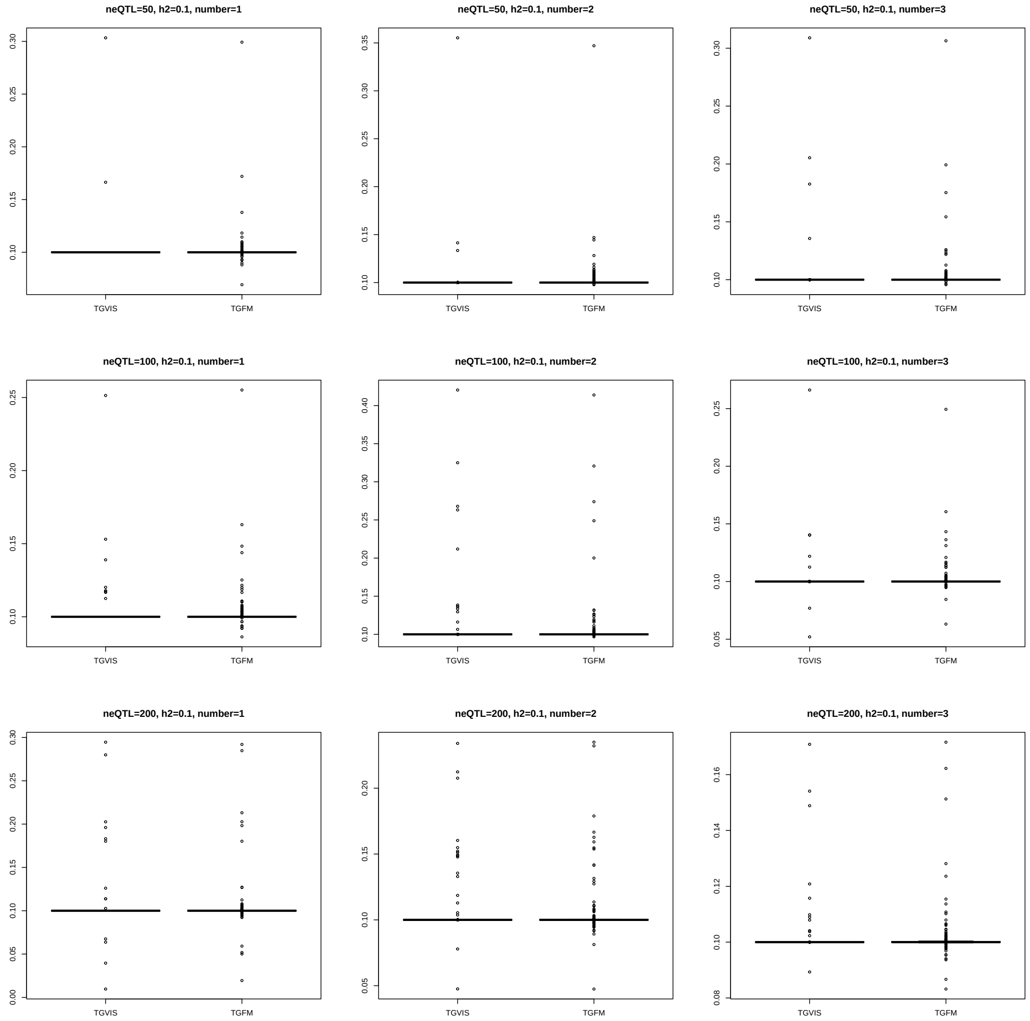

Figure S17: This figure shows the boxplot of the 2-norm of the difference between the eQTL effect estimates obtained by TGVIS and TGFM and the true values, i.e., $||\hat{\boldsymbol{\beta}}_{\mathrm{TGVIS}} - \boldsymbol{\beta}||_2$ and $||\hat{\boldsymbol{\beta}}_{\mathrm{TGFM}} - \boldsymbol{\beta}||_2$, across 100 simulations. The simulations are conducted with the heritability of gene-tissue pairs fixed at $h^2_{\text{tissue-gene}} = 0.1$, varying the number of eQTLs (number) and the eQTL sample size (neQTL).

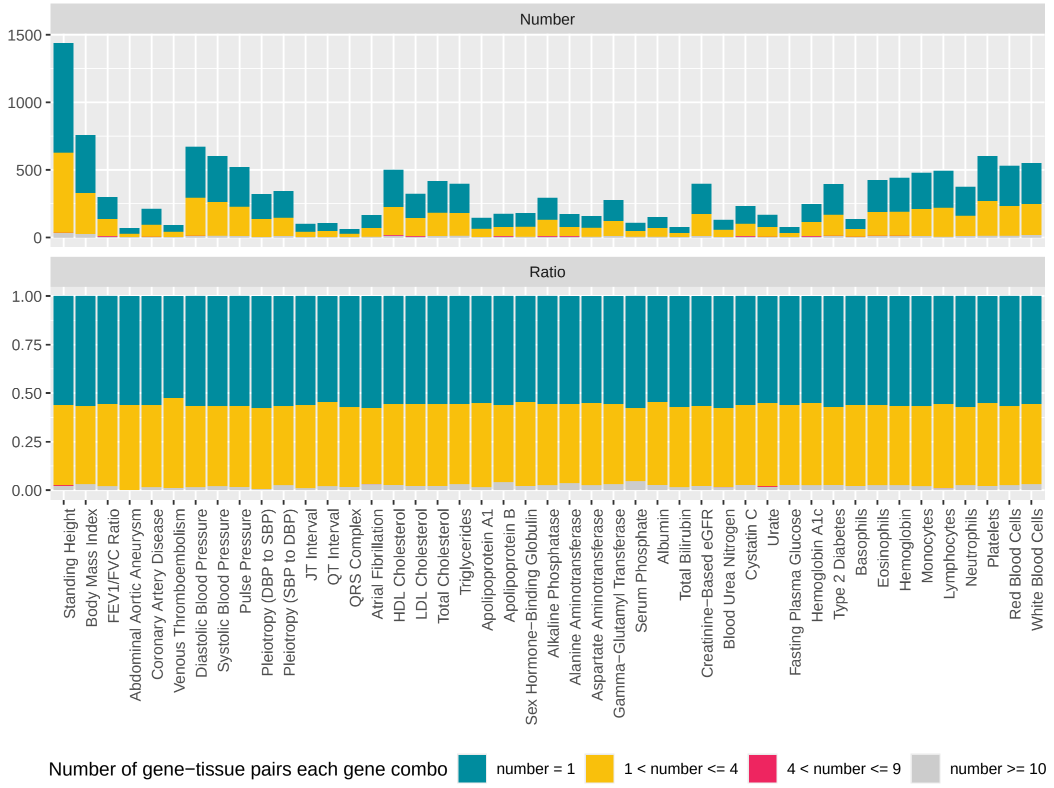

Figure S18: The figure above shows the total number of components in each credible set identified by **TGFM**. For each locus, we calculated the components involved in each credible set. Thus, the total count for each case (number = 1, $1 < \text{number} \leq 4$, $4 < \text{number} \leq 9$, $\text{number} \geq 10$) in the figure corresponds to the number of loci multiplied by the number of cases per locus. The figure below displays the proportion of these four cases averaged across each locus.

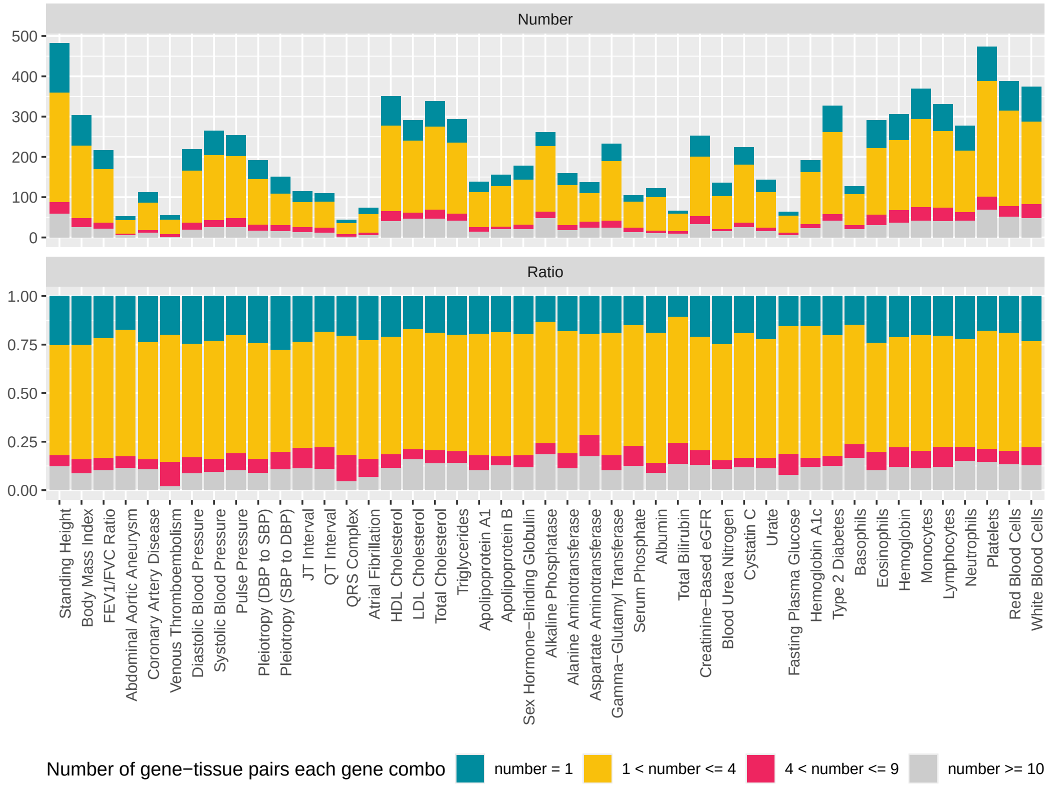

Figure S19: The figure above shows the total number of components in each credible set identified by **TGVIS**. For each locus, we calculated the components involved in each credible set. Thus, the total count for each case (number $= 1$, $1 <$ number $\leq 4$, $4 <$ number $\leq 9$, number $\geq 10$) in the figure corresponds to the number of loci multiplied by the number of cases per locus. The figure below displays the proportion of these four cases averaged across each locus.

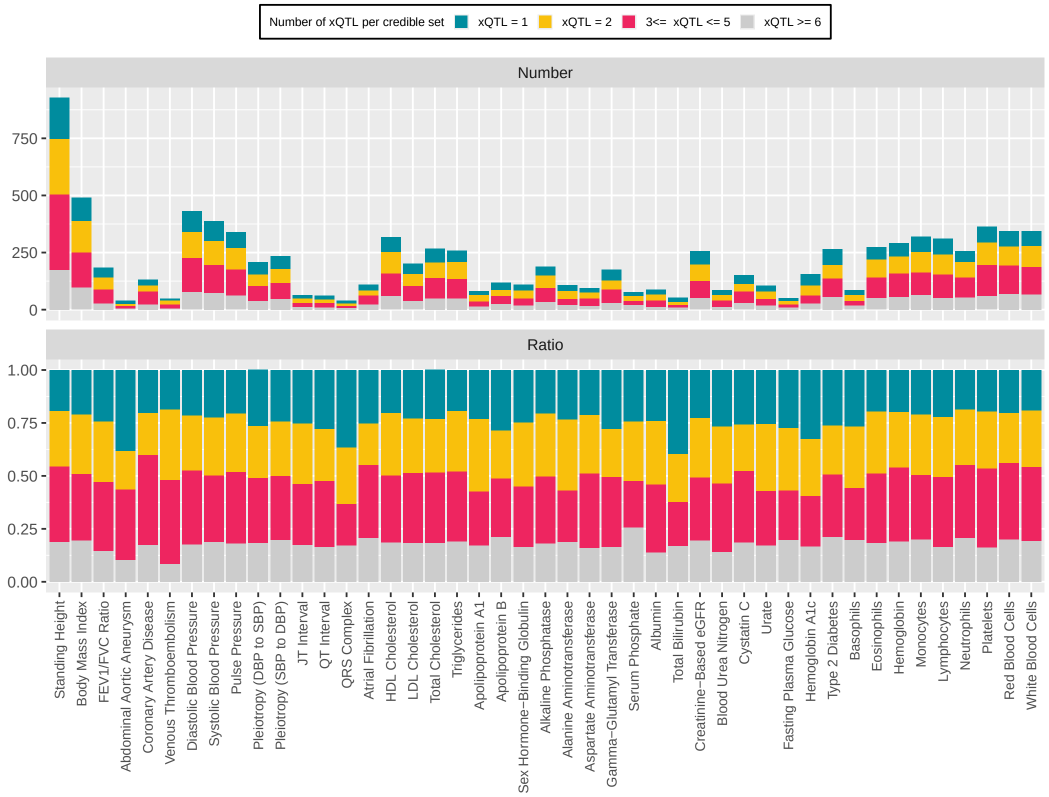

Figure S20: The figure above shows the total number of xQTL in each credible sets identified by **TGFM**. For each locus, we calculated the averaged number of xQTL of gene-tissue pairs in each credible set. Thus, the total count for each case (xQTL $= 1$, xQTL $= 2$, $3 \leq$ xQTL $\leq 5$, xQTL $\geq 6$) in the figure corresponds to the number of loci multiplied by the number of cases per locus. It is possible that different gene-tissue pairs within the same credible set have a different number of xQTLs. This often occurs when the shared xQTLs among these gene-tissue pairs have larger effects, while the non-overlapping xQTLs have smaller effects, leading them to be grouped into the same credible set. In such cases, we calculate the average number of xQTLs for the gene-tissue pairs within each credible set. The figure below displays the proportion of these four cases averaged across each locus. It's important to note that our approach differs from TGFM at this step by applying an additional threshold ('pip.min = 0.25'). This threshold requires not only that the coverage of the credible set is 0.95, but also that the minimum individual PIP within the credible set meets the 'pip.min' threshold. Since we have already removed highly correlated variants using the C+T approach, we believe that a credible set should not include too many variants in the xQTL selection Therefore, we apply this additional filter to remove some noise xQTL.

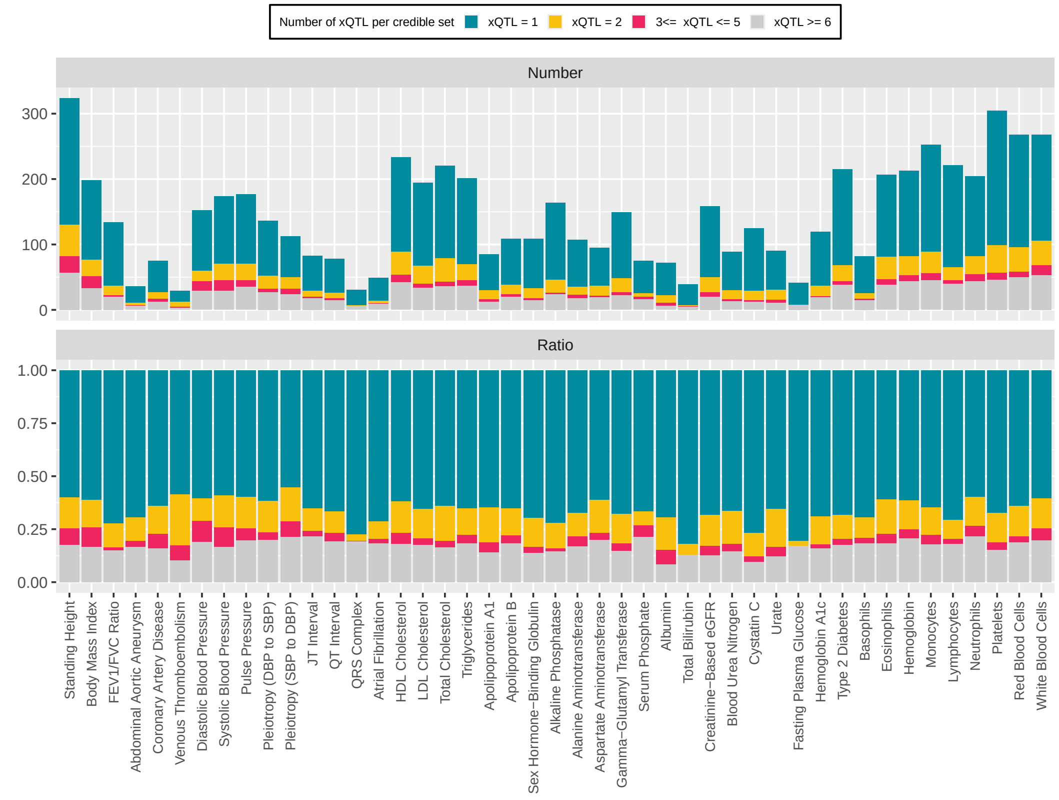

Figure S21: The figure above shows the total number of xQTL in each credible sets identified by **TGVIS**. For each locus, we calculated the averaged number of xQTL of gene-tissue pairs in each credible set. Thus, the total count for each case (xQTL $= 1$, xQTL $= 2$, $3 \leq$ xQTL $\leq 5$, xQTL $\geq 6$) in the figure corresponds to the number of loci multiplied by the number of cases per locus. It is possible that different gene-tissue pairs within the same credible set have a different number of xQTLs. This often occurs when the shared xQTLs among these gene-tissue pairs have larger effects, while the non-overlapping xQTLs have smaller effects, leading them to be grouped into the same credible set. In such cases, we calculate the average number of xQTLs for the gene-tissue pairs within each credible set. The figure below displays the proportion of these four cases averaged across each locus. It's important to note that our approach differs from TGFM at this step by applying an additional threshold ('pip.min = 0.25'). This threshold requires not only that the coverage of the credible set is 0.95, but also that the minimum individual PIP within the credible set meets the 'pip.min' threshold. Since we have already removed highly correlated variants using the C+T approach, we believe that a credible set should not include too many variants in the xQTL selection Therefore, we apply this additional filter to remove some noise xQTL.

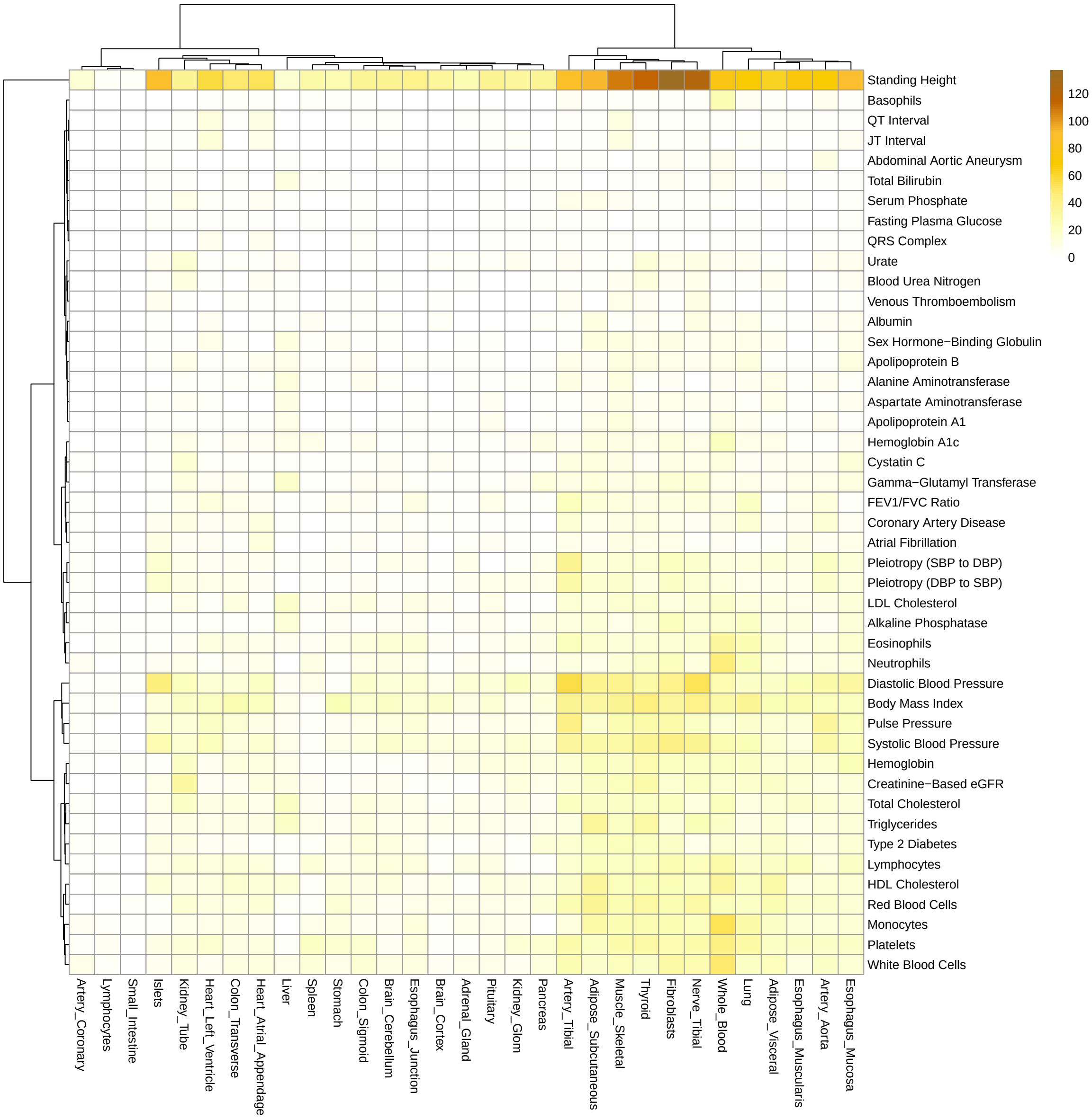

Figure S22: Heatmaps display the major tissues associated with each trait, identified using TGVIS. The major gene-tissue pairs are cataloged based on stringent criteria (PIP > 0.5). Hierarchical clustering is applied to arrange the heatmaps, utilizing the ‘average‘ method and ‘Manhattan‘ distance.

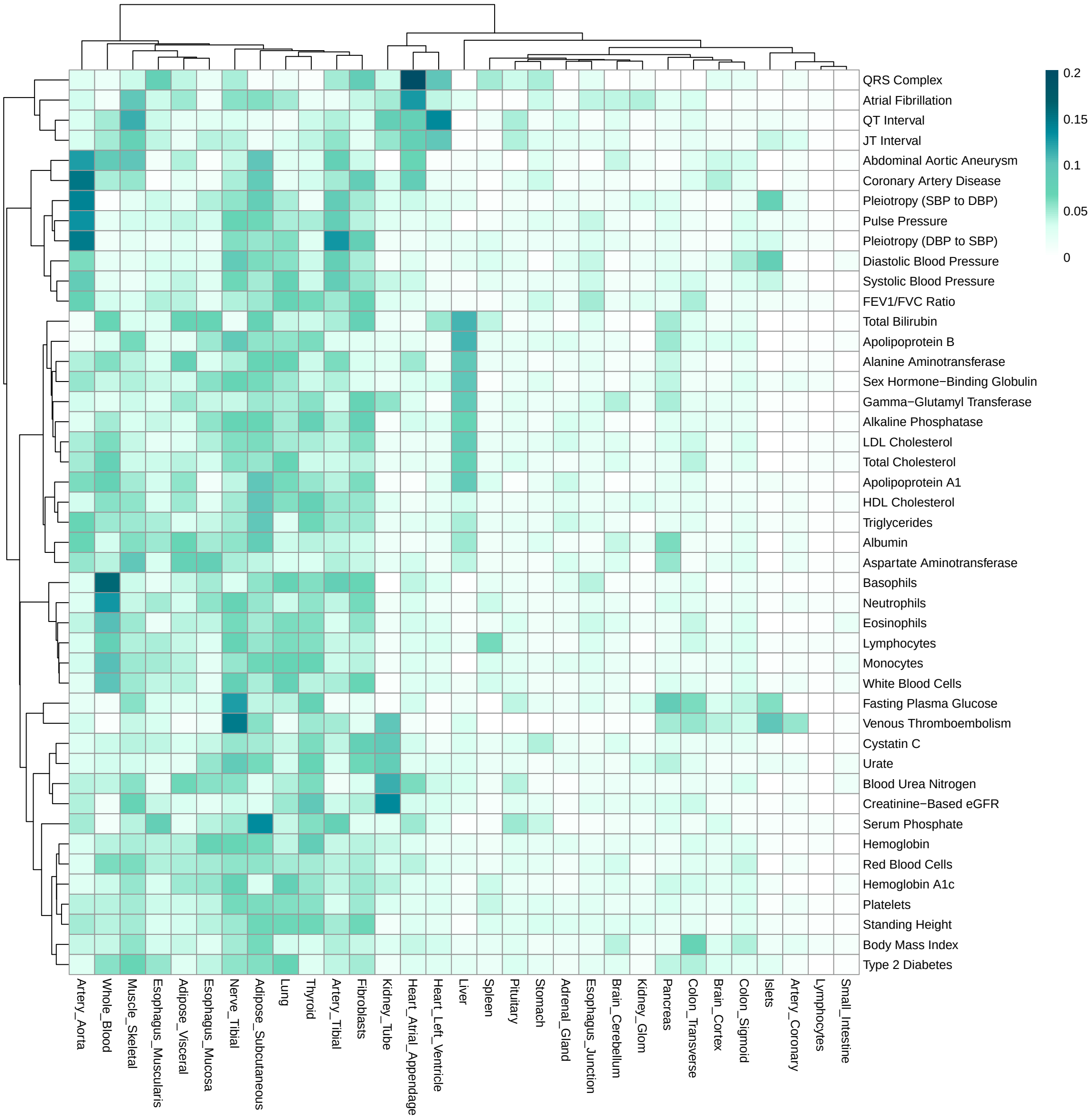

Figure S23: Heatmaps display the major tissues associated with each trait, identified using TGFM The major gene-tissue pairs are cataloged based on stringent criteria (CS-Pratt > 0.15). Hierarchical clustering is applied to arrange the heatmaps, utilizing the ‘average‘ method and ‘Manhattan‘ distance.

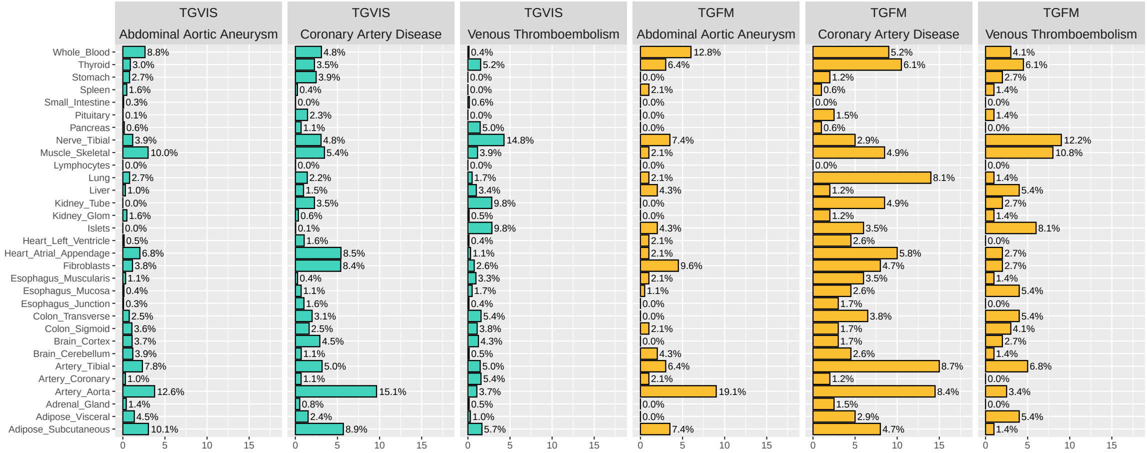

Figure S24: Major tissues of cardiovascular diseases identified by TGVIS and TGFM.

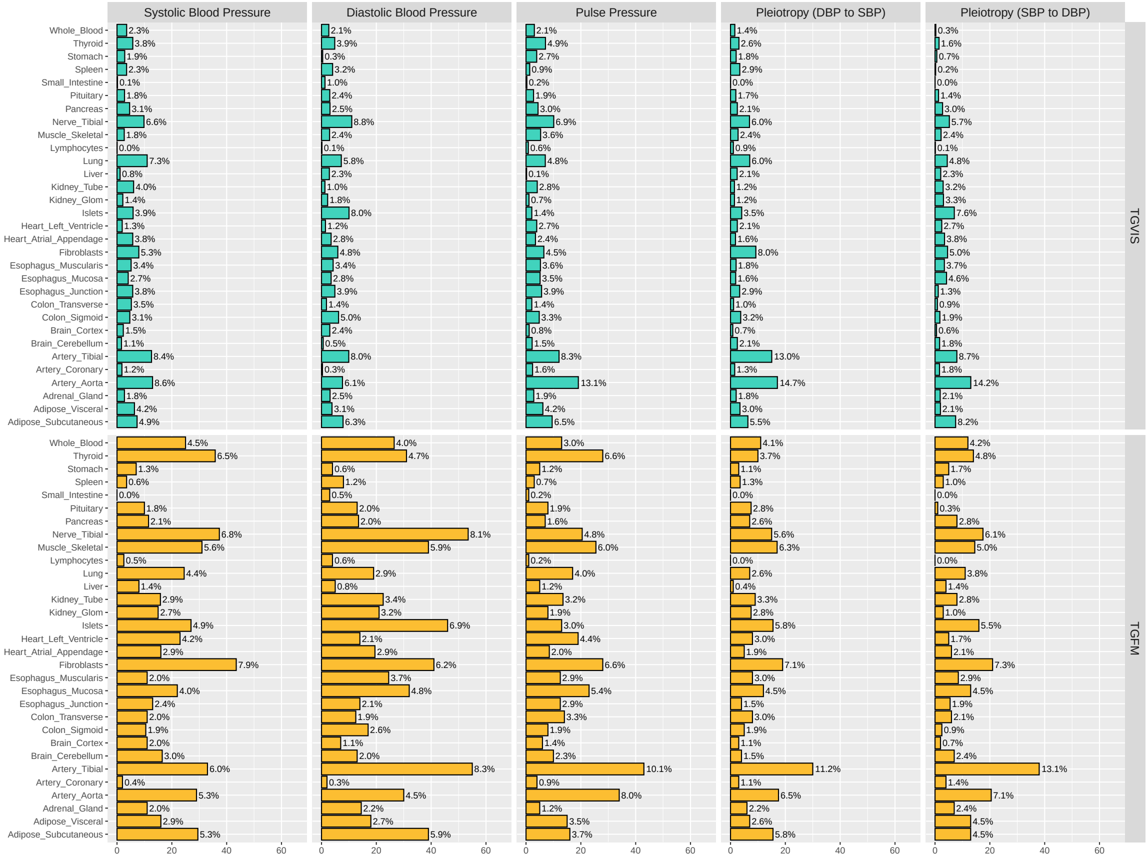

Figure S25: Major tissues of blood pressure traits identified by TGVIS and TGFM.

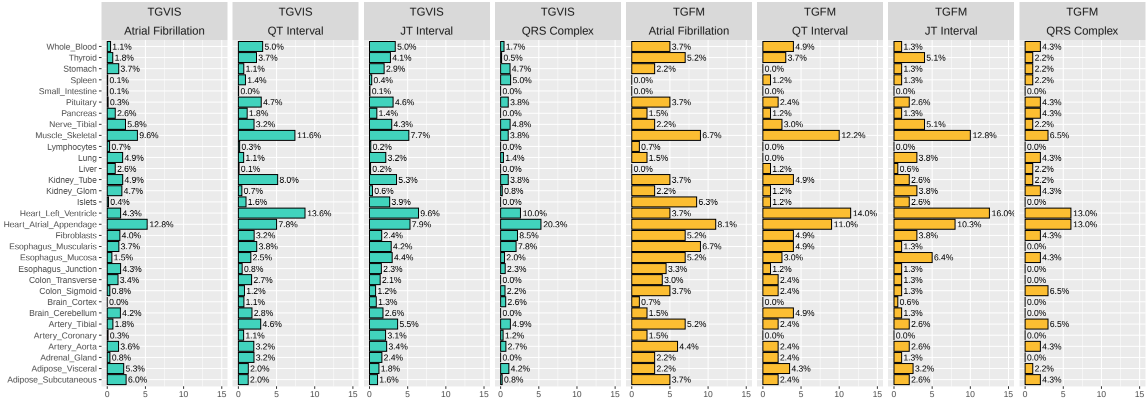

Figure S26: Major tissues of heart-related traits and disease identified by TGVIS and TGFM.

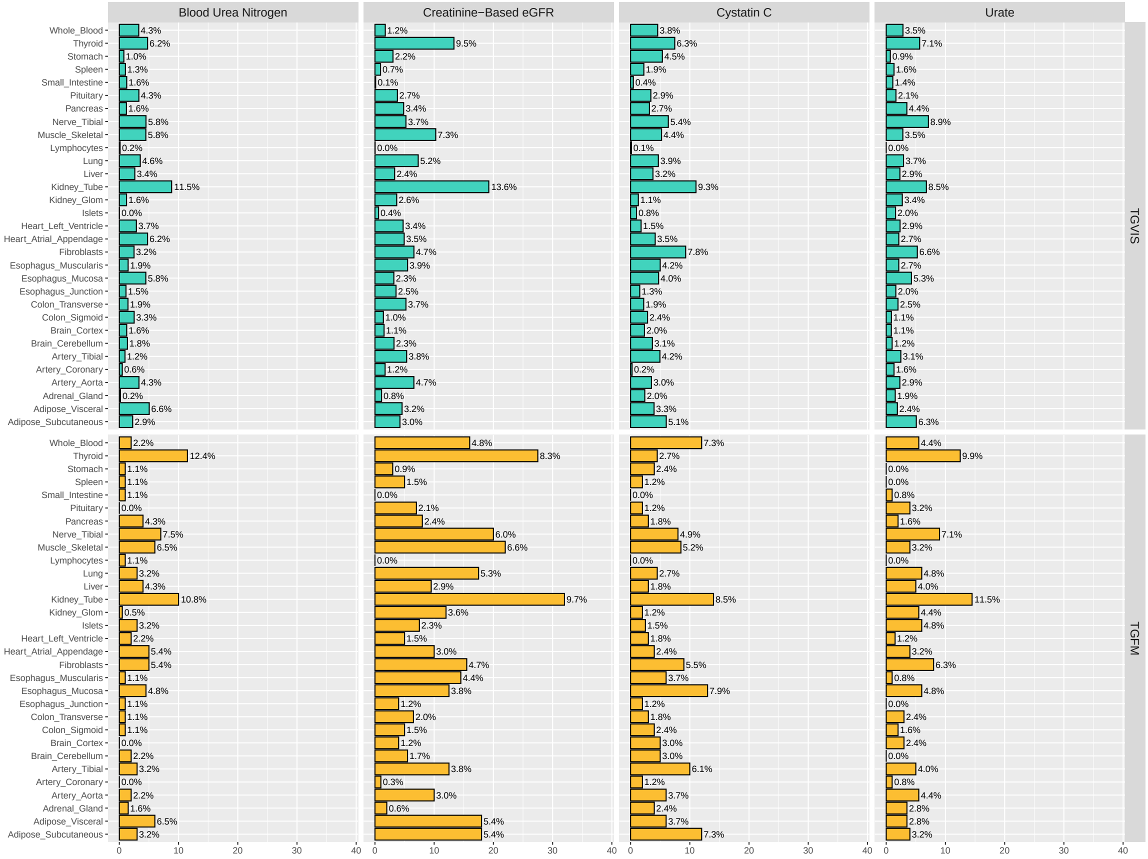

Figure S27: Major tissues of kidney-related traits identified by TGVIS and TGFM.

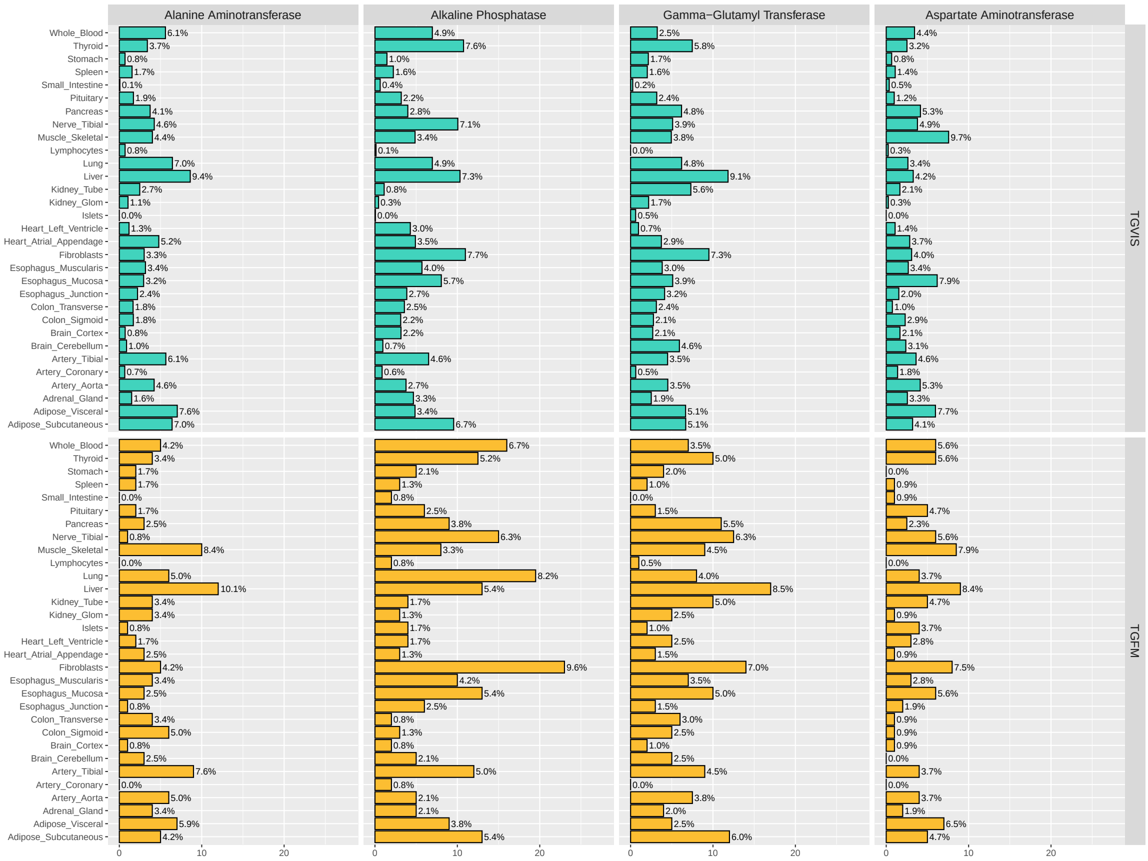

Figure S28: Major tissues of liver-related identified by TGVIS and TGFM.

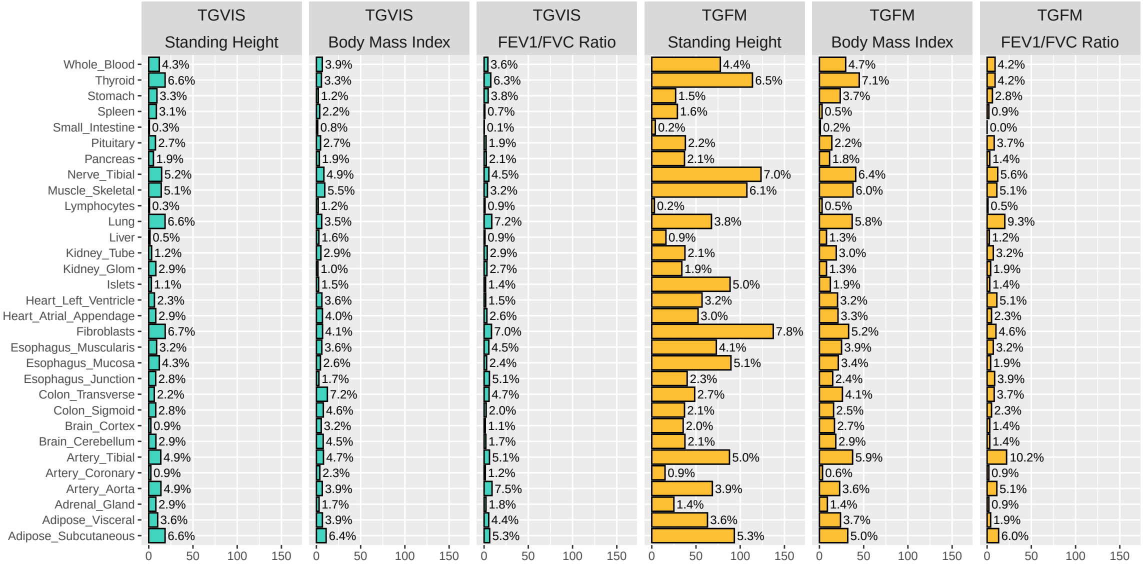

Figure S29: Major tissues of standing height, body mass index, and FEV1/FVC ratio identified by TGVIS and TGFM.

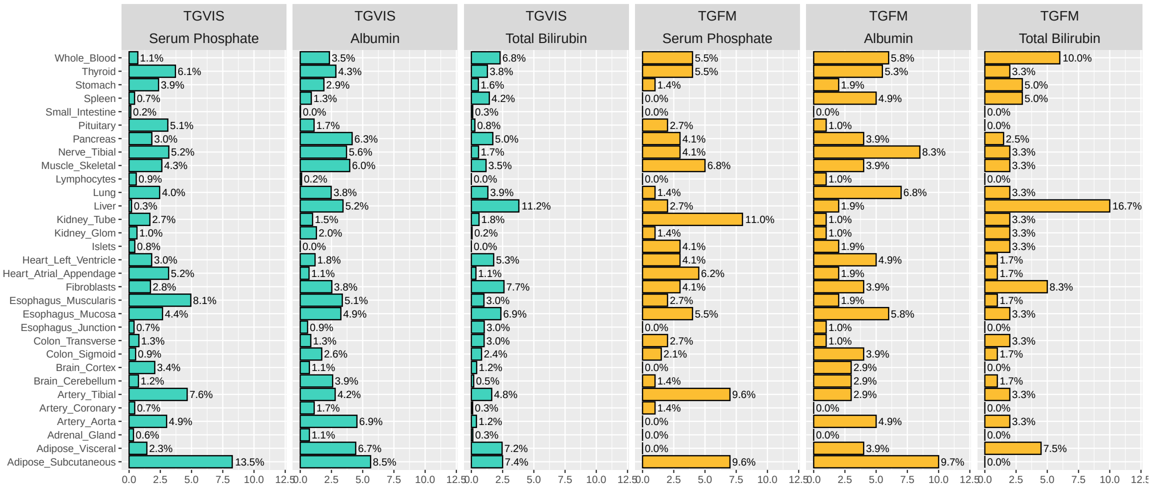

Figure S30: Major tissues of blood metabolic traits identified by TGVIS and TGFM.

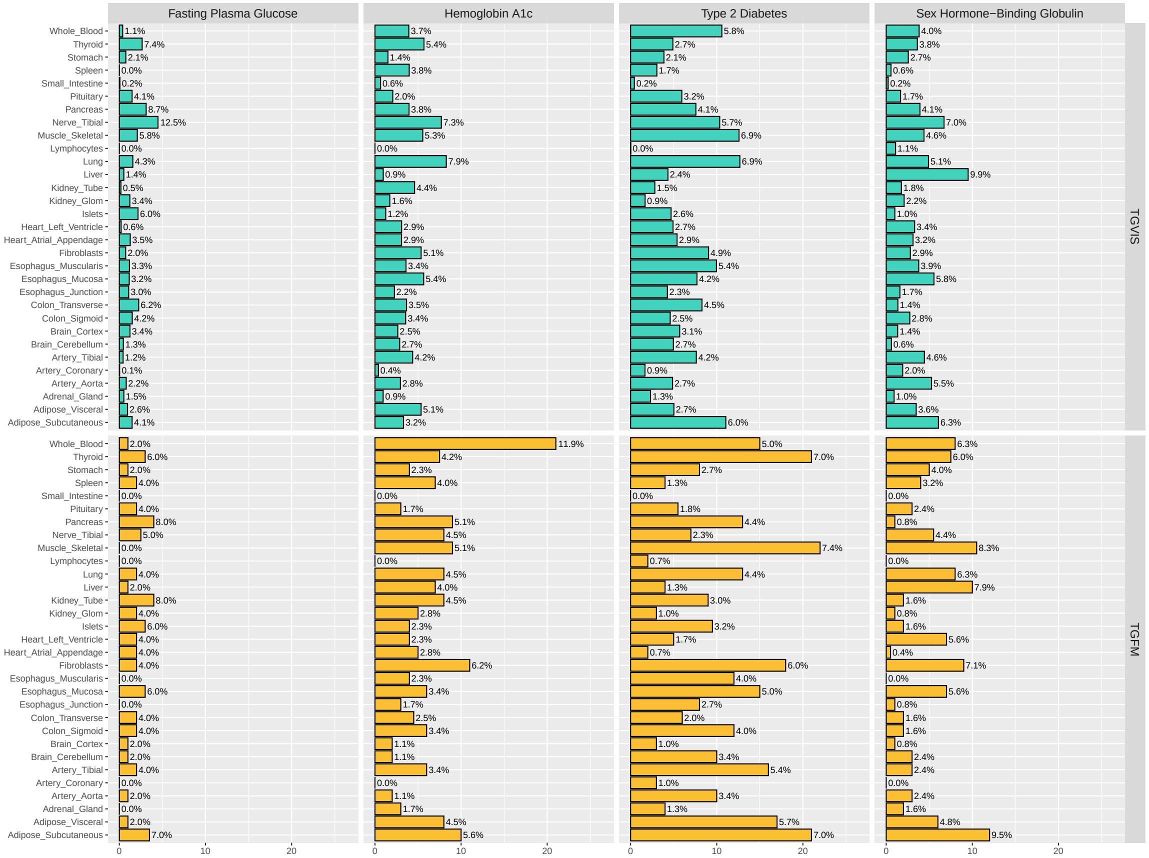

Figure S31: Major tissues of Type-2 diabetes and the related traits identified by TGVIS and TGFM.

### References

Bulik-Sullivan, B., Finucane, H. K., Anttila, V., Gusev, A., Day, F. R., Loh, P.-R., ReproGen Consortium, Psychiatric Genomics Consortium, Genetic Consortium for Anorexia Nervosa of the Wellcome Trust Case Control Consortium 3, Duncan, L., et al. (2015). An atlas of genetic correlations across human diseases and traits. Nature Genetics, **47**(11), pp.1236–1241.

Breheny, P. and Huang, J. (2011). Coordinate descent algorithms for nonconvex penalized regression, with applications to biological feature selection. The Annals of Applied Statistics, pp 232–253.

Lorincz-Comi, Noah. and Yang, Yihe. and Li, Gen. and Zhu, Xiaofeng. (2024). MRBEE: A bias-corrected multivariable Mendelian Randomization method. Human Genetics and Genomics Advances.

Yavorska, O.O. and Burgess, S. (2017). MendelianRandomization: an R package for performing Mendelian randomization analyses using summarized data. International Journal of Epidemiology, pp.1734-1739.

Yi, G. Y. (2016). Statistical analysis with measurement error or misclassification. Springer.

Zhao, Siming, Crouse, Wesley, Qian, Sheng, Luo, Kaixuan, Stephens, Matthew, and He, Xin (2024). Adjusting for genetic confounders in transcriptome-wide association studies improves discovery of risk genes of complex traits. Nature Genetics, 56(2), 336–347. Nature Publishing Group US New York.

Strober, Benjamin J., Zhang, Martin Jinye, Amariuta, Tiffany, Rossen, Jordan, and Price, Alkes L. (2023). Fine-mapping causal tissues and genes at disease-associated loci. medRxiv. Cold Spring Harbor Laboratory Preprints.

Chen, Jiahua, and Chen, Zehua (2012). Extended BIC for small-n-large-P sparse GLM. Statistica Sinica, 555–574. JSTOR.

Fan, Jianqing, Liao, Yuan, and Mincheva, Martina (2013). Large covariance estimation by thresholding principal orthogonal complements. Journal of the Royal Statistical Society Series B: Statistical Methodology, 75(4), 603–680. Oxford University Press.

Cavicchioli, Maddalena, Forni, Mario, Lippi, Marco, and Zaffaroni, Paolo (2016). Eigenvalue Ratio Estimators for the Number of Common Factors. CEPR Discussion Paper No. DP11440.

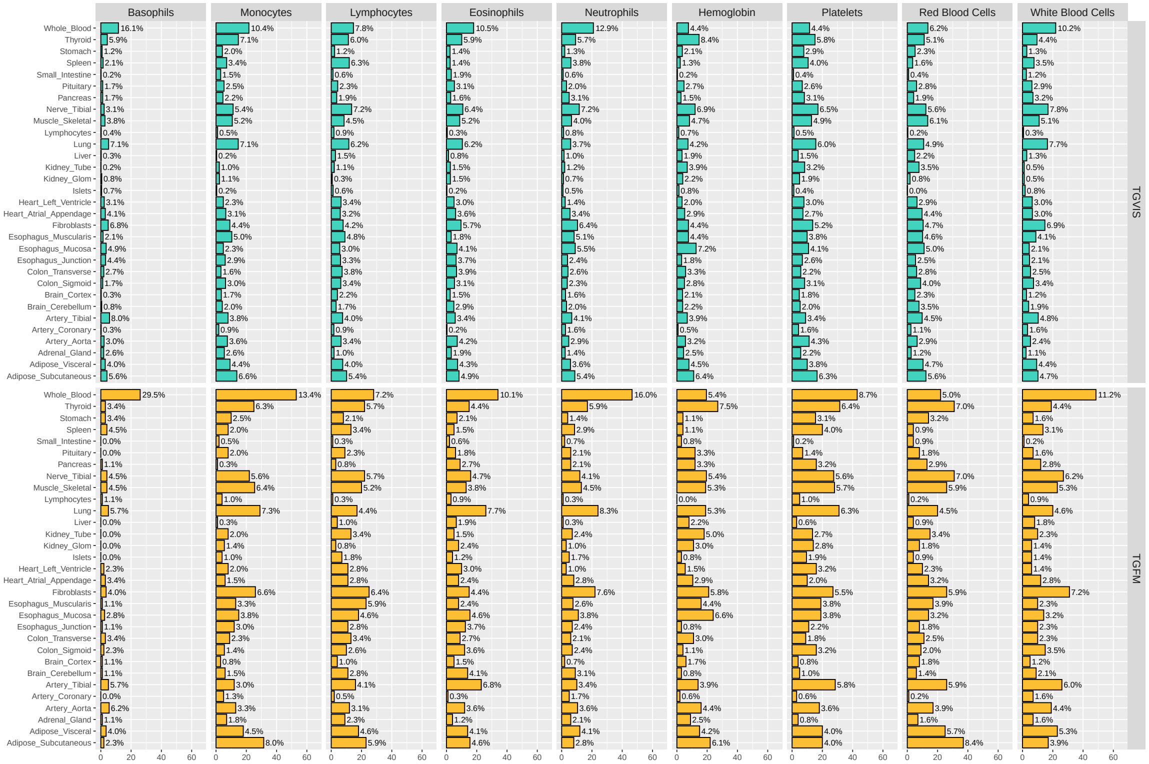

Figure S32: Major tissues of blood cell traits identified by TGVIS and TGFM.

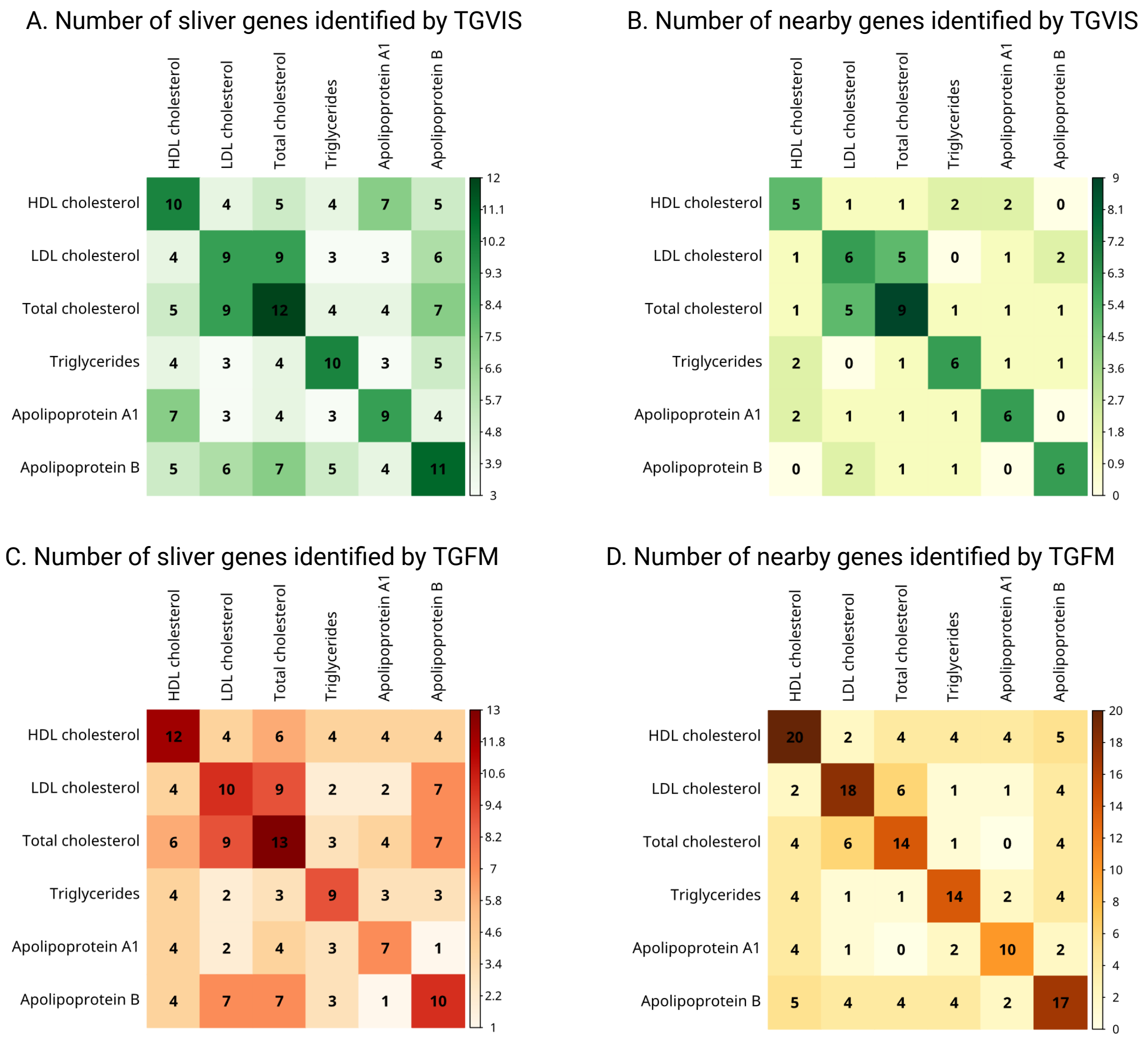

Figure S33: Overlap of trait-specific silver genes and bystander genes across different lipid traits, highlighting increased overlap among traits with high genetic correlation. We present the results for the silver standard of lipid genes. We examined the genes associated with HDL-C, LDL-C, TC, TG, APOA1, and APOB within the silver and bystander categories, and validated the pairwise overlap of these trait-specific silver genes and bystander genes across different lipid traits. The results indicate that there is some overlap between trait-specific silver genes and bystander genes across different traits, especially among traits with high genetic correlation (e.g., LDL-C and APOB).

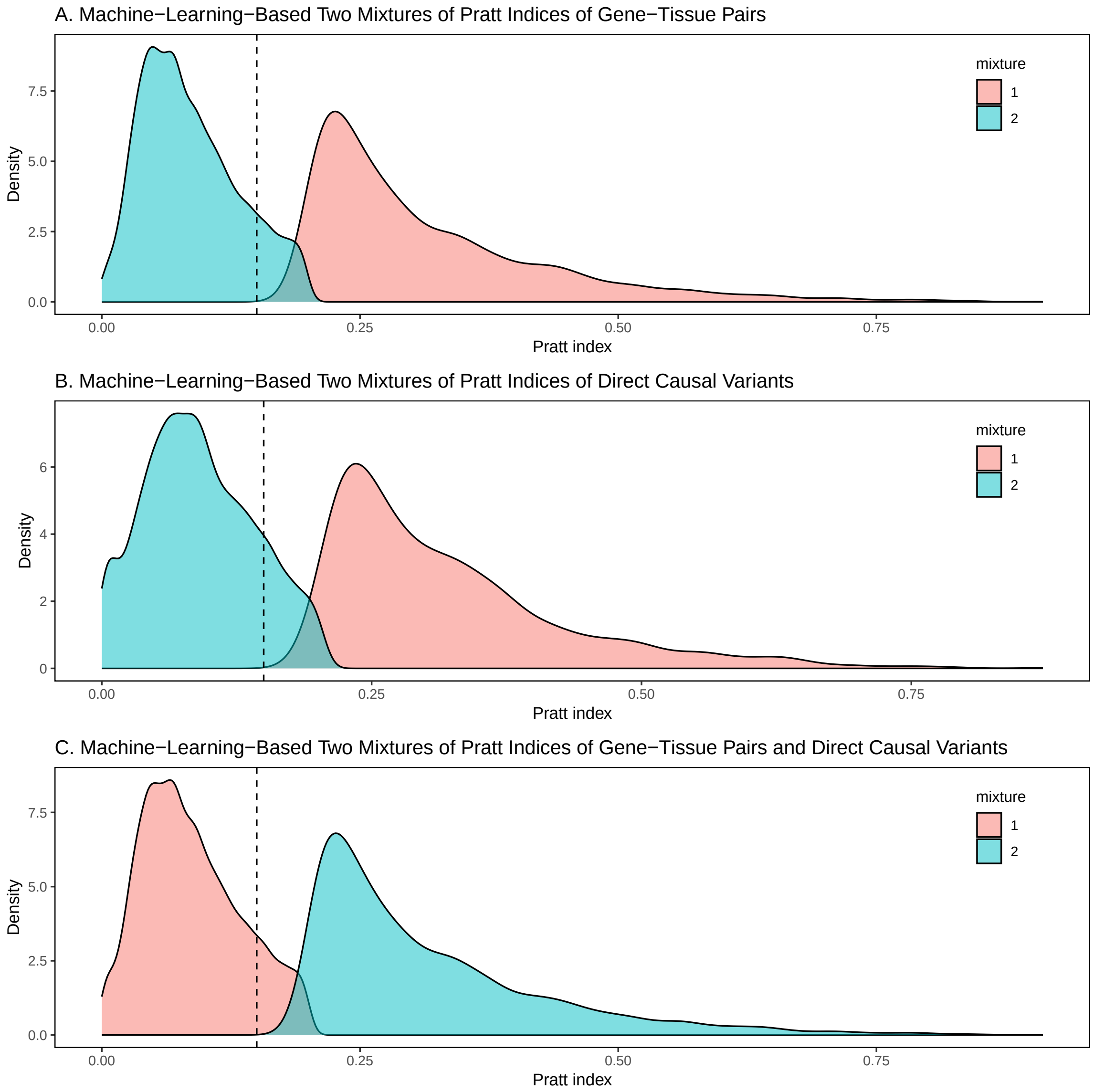

Figure S34: Mixture distributions of CS-Pratt indices for A gene-tissue pairs, B direct causal variants, and C both combined, identified by TGVIS within the 95

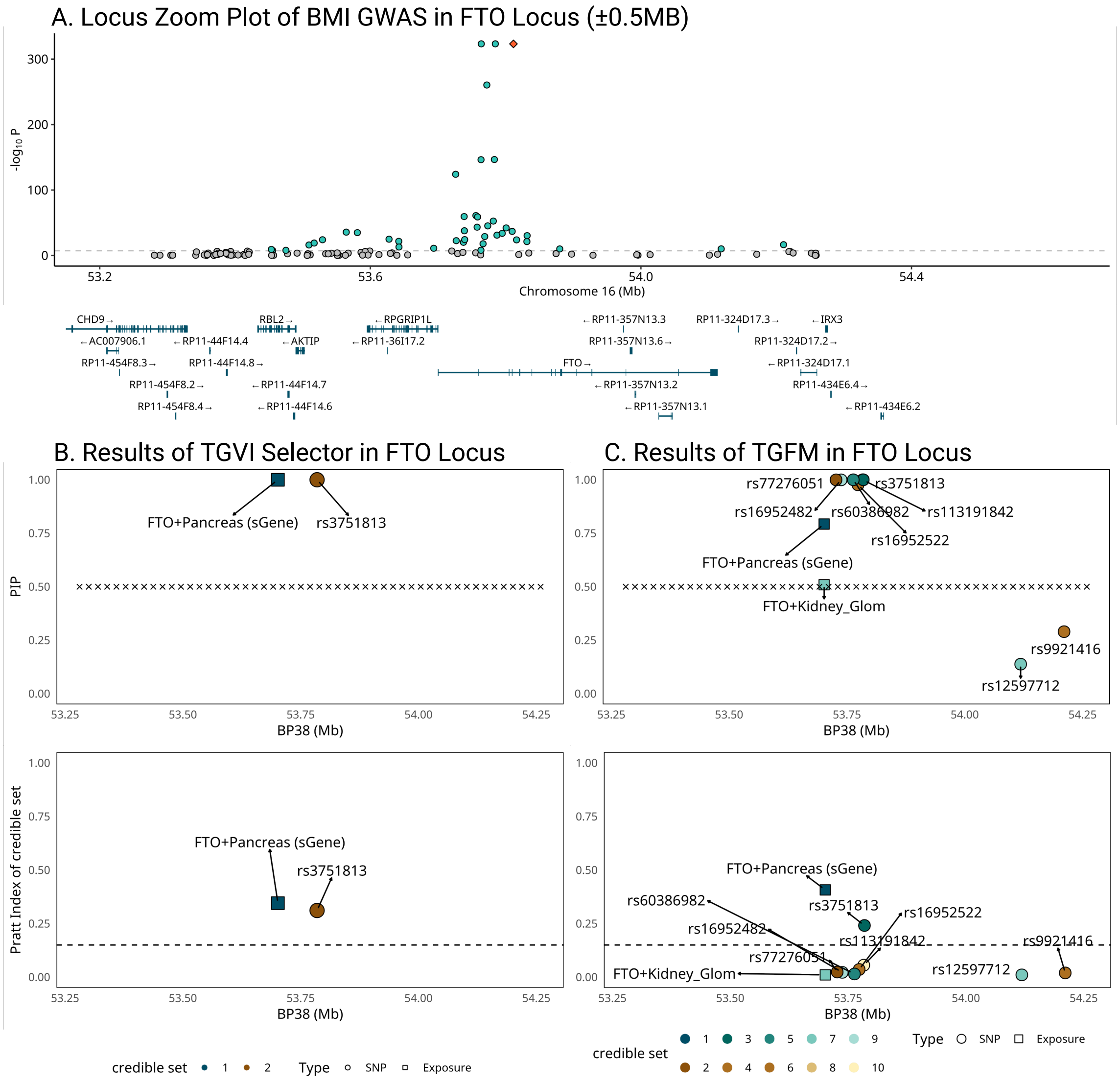

Figure S35: LocusZoom plots comparing the results of TGVIS and TGFM methods. This figure presents LocusZoom plots for various loci associated with different traits, illustrating the outcomes from both TGVIS and TGFM methods. We show the results of TGVIS and TGFM for BMI in the FTO locus, when the window size is ±500 KB.

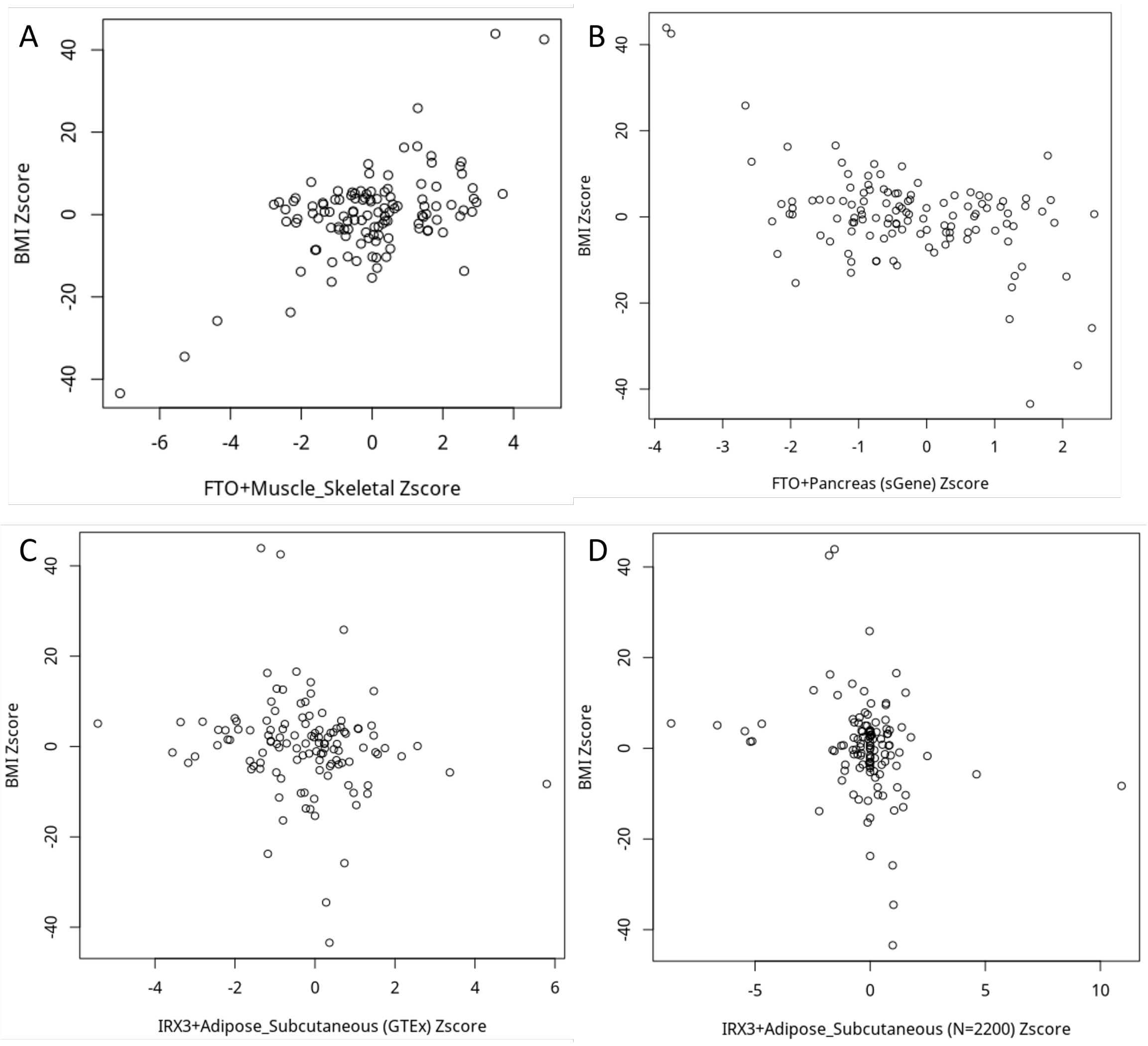

Figure S36: Scatter plots of Z-scores of eQTL effects versus Z-scores of GWAS effects for gene-tissue pairs used in the final TGVIS and TGFM analyses. Panels A–D correspond to FTO in skeletal muscle, FTO in pancreas (sGene), IRX3 in subcutaneous adipose tissue, and IRX3 in subcutaneous adipose tissue using eQTL summary data from https://www.biorxiv.org/content/10.1101/2023.10.26.563798v1, respectively. Only eQTLs that were included in the final analyses are shown, after filtering using the C+T method to remove highly correlated variants (r2>0.5) and low-power eQTLs (P>1E-5). Note that due to very low power, IRX5 was filtered out by S-PrediXcan and was not included in the final TGVIS and TGFM stages.

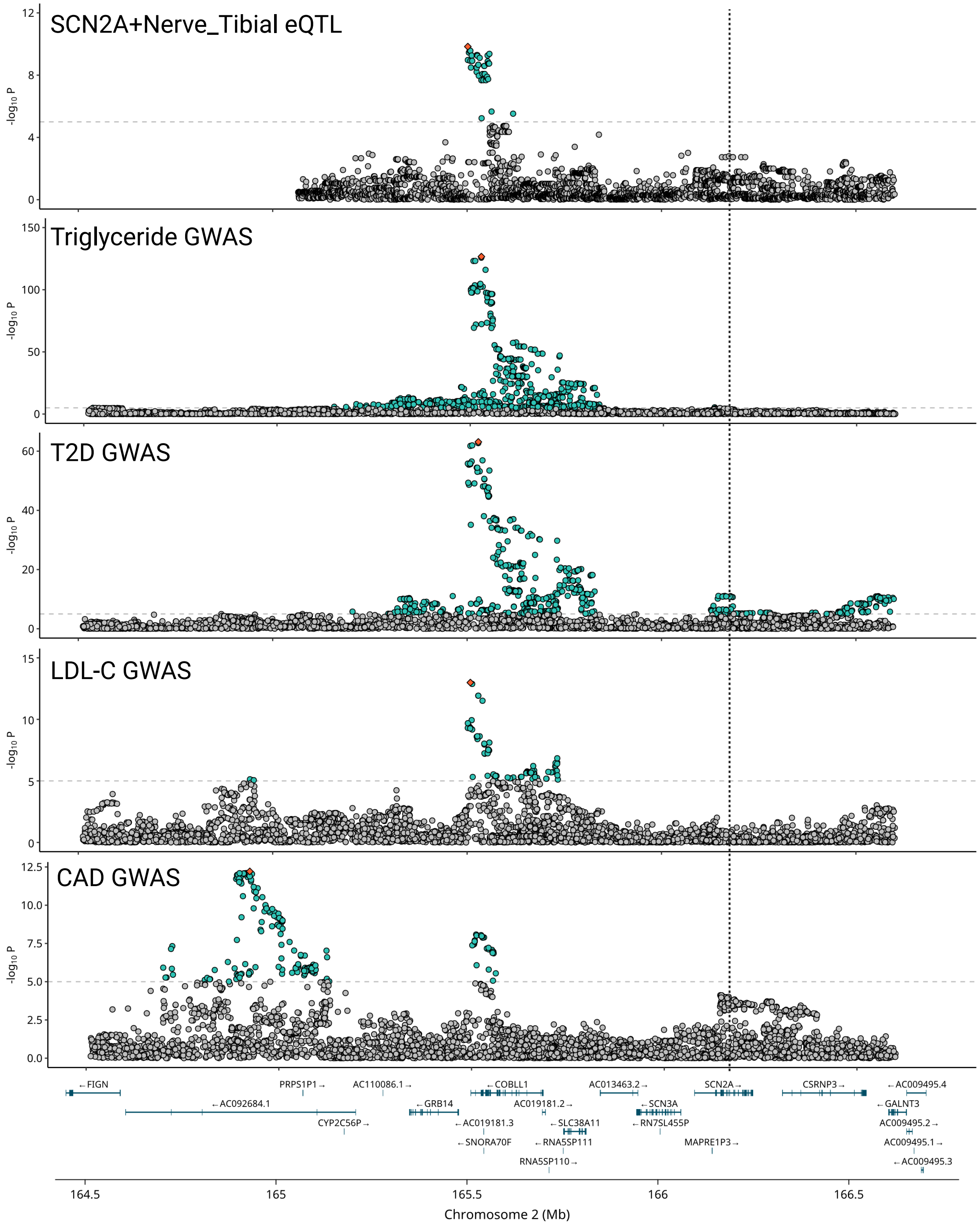

Figure S37: SCN2A-Nerve_Tibial is an example, identified by TGVIS as a causal gene-tissue pair for 18 traits, whereas TGFM identified it for only 6 traits. We presented the locus zoom plot of SCN2A-Nerve_Tibial with TG, T2D, LDL-C, and CAD in this region, where TGFM did not recognize SCN2A-Nerve_Tibial's causality for T2D and CAD. We found that the genetic variation patterns of these traits and this gene-tissue pair are highly similar in this region.

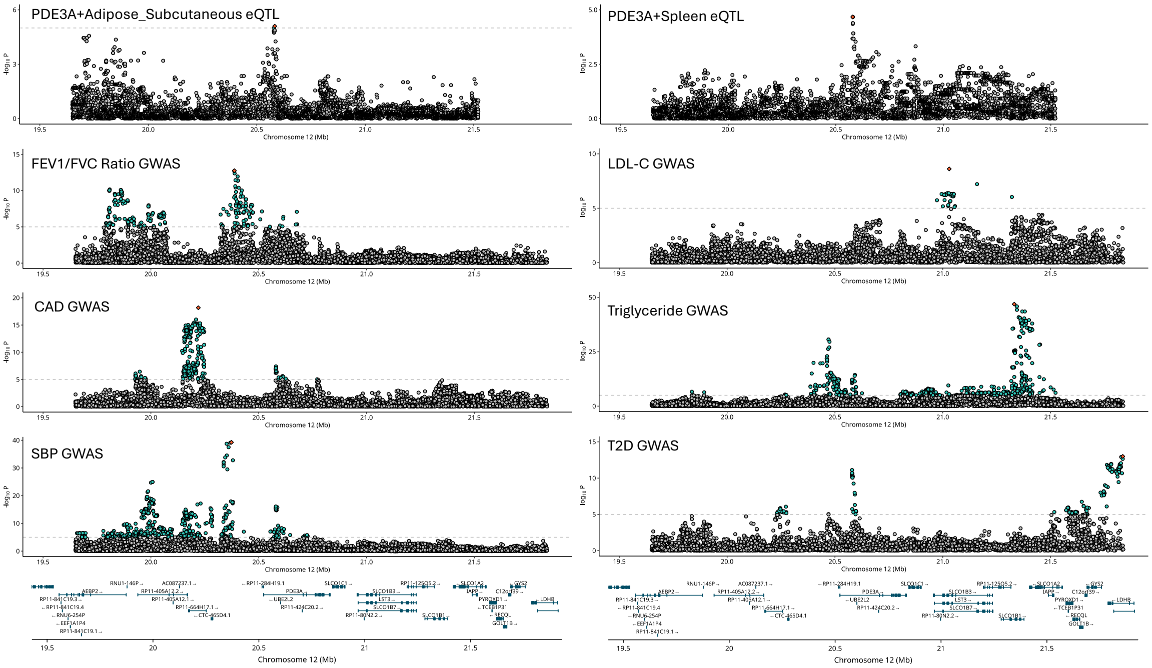

Figure S38: PDE3A-Adipose_Subcutaneous is another example, identified by TGFM as a causal pair for 12 traits, but not by TGVIS. We presented the locus zoom plot of PDE3A-Adipose_Subcutaneous and PDE3A-Spleen for six traits, including the FEV1FVC ratio, in the PDE3A region. It can be observed that the local genetic variation patterns of these two eGenes do not match those of any of the displayed traits. This suggests that the identification of this gene-tissue pair may be influenced by biases due to infinitesimal effects.
